# The metabolomic signature of weight loss in the Diabetes Remission Clinical Trial (DiRECT)

**DOI:** 10.1101/2022.07.15.22277671

**Authors:** Laura J. Corbin, David A. Hughes, Caroline J. Bull, Emma E. Vincent, Madeleine L. Smith, Alex McConnachie, Claudia-Martina Messow, Paul Welsh, Roy Taylor, Michael E. J. Lean, Naveed Sattar, Nicholas J. Timpson

**Affiliations:** MRC Integrative Epidemiology Unit at University of Bristol, Bristol, BS8 2BN, UK; Population Health Sciences, Bristol Medical School, University of Bristol, Bristol, BS8 2BN, UK; School of Translational Health Sciences, Dorothy Hodgkin Building, University of Bristol, Bristol, BS1 3NY, UK; Robertson Centre for Biostatistics, Institute of Health and Wellbeing, University of Glasgow, Glasgow G12 8QQ, UK; Institute of Cardiovascular and Medical Science, University of Glasgow G12 8TA, UK; Newcastle Magnetic Resonance Centre, Translational and Clinical Research Institute, Newcastle University, Newcastle upon Tyne NE4 5PL, UK; Human Nutrition, School of Medicine, Dentistry and Nursing, College of Medical, Veterinary & Life Sciences, University of Glasgow, Glasgow, G31 2ER, UK

## Abstract

Use of high-throughput metabolomics technologies in a variety of study designs has demonstrated a strong and consistent metabolomic signature of overweight and type 2 diabetes. However, the extent to which these metabolomic patterns can be recovered with weight loss and diabetes remission has not been investigated. We aimed to characterise the metabolomic consequences of a weight loss intervention in diabetes, within an existing randomised controlled trial – the Diabetes Remission Clinical Trial (DiRECT) – to provide insight into how weight loss-induced metabolic changes could lead to improved health. Decreases in branched chain amino acids, sugars and LDL triglycerides, and increases in sphingolipids, plasmalogens and metabolites related to fatty acid metabolism were associated with the intervention. The change in metabolomic pattern with mean 8.8kg weight loss thus reverses many features associated with the development of type 2 diabetes. Furthermore, metabolomic profiling also appears to capture variation in response to treatment seen across patients.

## Introduction

For conditions like type 2 diabetes where there is clear relationship between risk factors, intermediate metabolic phenotypes and disease, attention has turned to metabolomics as a potentially useful tool in elucidating the precise biological mechanisms underpinning disease pathology (1). Metabolomics can be defined as the comprehensive measurement of all metabolites and low-molecular-weight molecules in a biological specimen (2), typically via mass spectrometry (MS) or nuclear magnetic resonance (NMR) spectroscopy. The application of these approaches within a variety of study designs has the potential to disentangle directionality of associations between metabolites and risk. Metabolomic studies to date have demonstrated a strong and consistent metabolomic signature of type 2 diabetes (3). The majority of studies have focused on blood-based samples (plasma/serum) with urine metabolites also having been evaluated (4). The population-based KORA (Cooperative Health Research in the Region of Augsburg) study reported an expected shift in glucose and many related carbohydrate metabolites, an increase in the branched chain amino acids (BCAA) and their derivatives, some perturbation of lipid metabolism and mild indicators of ketosis in those with type 2 diabetes as compared to unaffected controls (5). Since then, evidence has accumulated to confirm most, if not all, of these preliminary findings in case/control studies (3).

Increasingly, focus has shifted to the study of incident cases and the evaluation of metabolomics as a tool for risk prediction (6). Perhaps unsurprisingly given the strong overlap in the metabolomic signature of type 2 diabetes and its precursors (obesity and insulin resistance) (7, 8), many of the metabolomic perturbations observed in patients diagnosed with disease also appear to have a role in predicting disease development (9, 10). In particular, the relevance of branched chain and other (e.g. aromatic) amino acids to insulin resistance in diabetes and obesity more generally, has come under increasing scrutiny as researchers seek to determine if this association is causal (11, 12). However, the question of whether the changes observed reflect a systemic response to high glucose and other disease-driven perturbations, or whether the metabolic changes are causative and themselves lead to the development of disease remains unanswered. Therefore, whilst there is evidence to suggest metabolomic profiling may help identify individuals at highest risk of developing type 2 diabetes in the future (13–16), whether these same metabolites are biomarkers of pathways relevant to prevention and treatment is unclear.

A complementary study design that has the potential to enhance the literature is the evaluation of the metabolomic response to disease remission following intervention. Weight loss is the intervention strategy most commonly evaluated but a recent review highlighted a number of issues that limit the reliability and robustness of results published to date (17). As such, there remains a requirement to characterize the molecular underpinnings of the events surrounding existing interventions targeting diabetes remission. We aimed to conduct a comprehensive evaluation of the metabolic consequences of weight loss in patients with type 2 diabetes within a clinical setting. Samples from a seminal randomised controlled trial (RCT) involving an intensive weight management programme were used to establish a causal framework for the analysis.

## Results

### Study characteristics

DiRECT is a two-year open-label, cluster-randomised controlled trial and is registered with the ISRCTN registry, number 03267836. The trial was carried out to assess whether effective weight management, delivered in a primary care setting, could produce sustained remission of type 2 diabetes. The trial was conducted over a two-year period with principal data and sample collection points scheduled at baseline, 12 months, and two years. In this study, we analysed samples from the baseline and 12-month time points using both an untargeted MS approach (Metabolon, Inc.) and ^1^H-NMR spectroscopy (Nightingale Health). We analysed all available data according to group allocation and as such, the results from this analysis represent the intervention effect in a clinical setting (i.e., accounting for non-compliance). After quality control (QC) filtering, there were 258 individuals with NMR data (115 intervention and 143 control) and 261 individuals with MS data (117 intervention and 144 control) available for statistical analysis. Previous work has shown the DiRECT cohort can be considered representative of the general population of people with short duration type 2 diabetes (18) and that baseline characteristics were similar between the control and intervention groups (Table 1 in (19)). Similarly, here, baseline characteristics were similar between the intervention and control groups in the subset of participants with metabolomics data available (**Table 1**).

**Table 1.** Baseline characteristics (N=261) Summary statistics calculated based on the MS sample (after quality control) (N=261).

|  | Intervention group (n=117) |  | Control group (n=144) |  |
| --- | --- | --- | --- | --- |
|  | Mean/n | St. Dev./% | Mean/n | St. Dev./% |
| Sex |  |  |  |  |
| Female | 49 | 42% | 55 | 38% |
| Male | 68 | 58% | 89 | 62% |
| Age (years) | 53.7 | 7.1 | 56.2 | 6.9 |
| Body mass index (kg/m<sup>2</sup>) | 34.8 | 4.5 | 34.3 | 4.3 |
| Weight (kg) | 100.3 | 16.8 | 99.0 | 16.0 |
| Fasting glucose (mmol/L) | 9.3 | 3.2 | 8.8 | 2.6 |
| Total cholesterol (mmol/L) | 4.3 | 1.1 | 4.3 | 1.1 |
| HDL cholesterol (mmol/L) | 1.1 | 0.3 | 1.2 | 0.3 |
| Triglycerides (mmol/L) | 2.0 | 1.5 | 1.9 | 0.9 |

An overview of the statistical analysis is shown in **Figure 1**. In all analyses, the control group is considered the reference group with effect estimates therefore representing the difference seen in the intervention group relative to the control group. In our evaluation of the effect of the intervention on metabolite levels, outcomes (metabolite levels at 12-months) were compared between groups using two modelling approaches. First, linear regression models were applied to rank normal transformed (RNT) metabolite levels to give estimates (betas) of the adjusted mean difference in metabolite levels in normalized standard deviation (SD) units. Secondly, logistic regression models were applied to metabolite data after transformation to a presence/absence (PA) phenotype to give estimates (betas) of an average change in the log odds of metabolite presence. Based on model performance (**Supplementary Figure S1**), the linear regression model formed the primary result for all metabolite features with less than 40%missing (unquantified) data at the 12-month timepoint whilst results from the logistic regression model were considered the primary result for all features with greater than or equal to 40%missing (unquantified) data.

**Figure 1.**
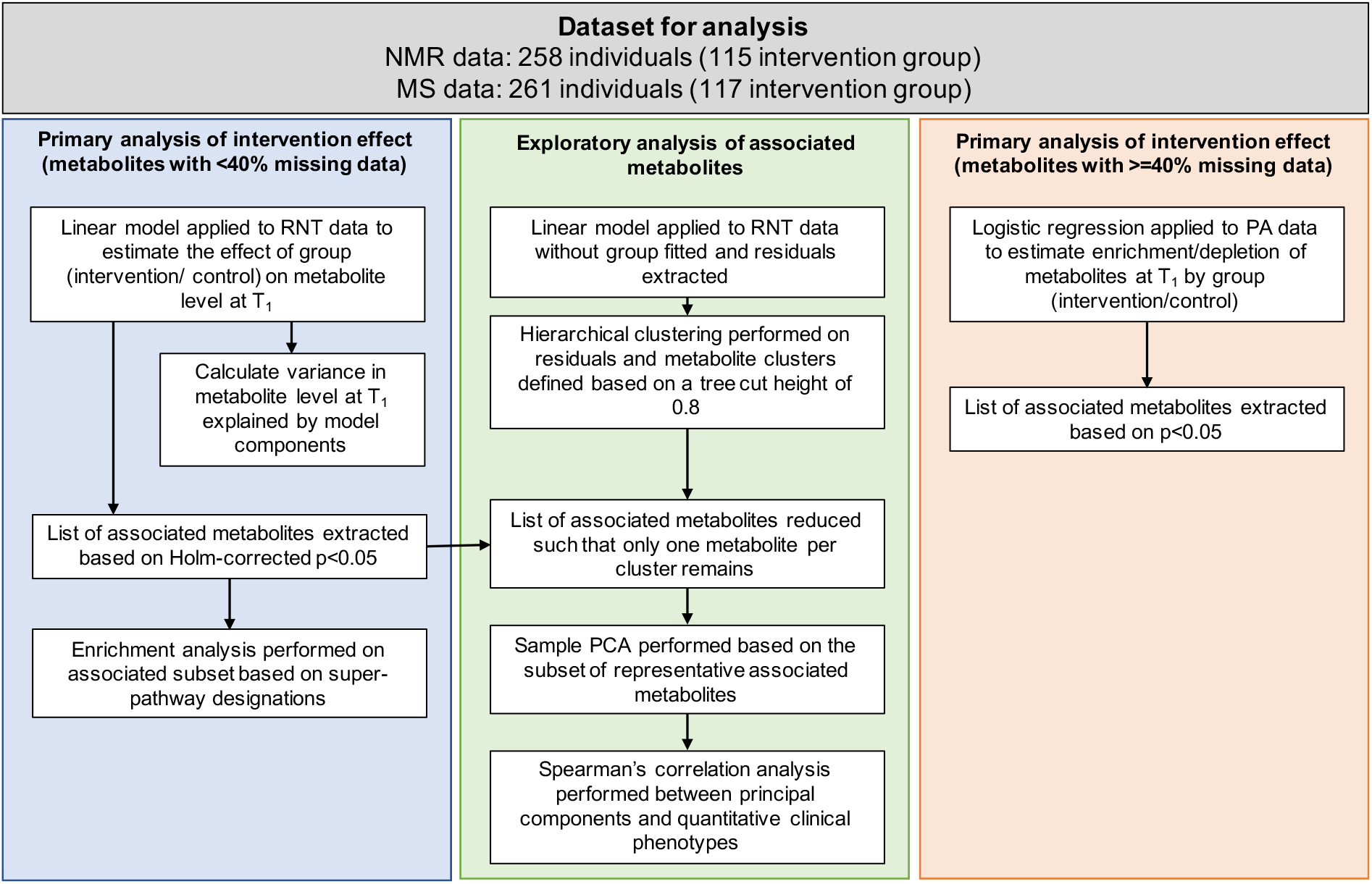
Statistical analysis overview. MS = mass spectrometry; NMR = nuclear magnetic resonance; RNT = data transformed using a rank based inverse normal transformation; PA = data transformed to a presence/absence phenotype with missing values replaced with 0 and non-missing values with 1; T_1_ = 12-month time point (post-intervention).

### Effect of intervention on metabolites – linear regression model

Results from the linear model formed the primary result for all 147 NMR metabolites and 78 NMR derived measures; in this analysis, the minimum (median) sample size was 199 (258). Of the NMR metabolites tested at one year, 59 (26%) were altered by the intervention (Holm-corrected *p*<0.05) (including 27 derived measures) with 41 (69%) showing an increase in response to treatment (**Figure 2** and **Supplementary Table S2**). Results from the linear model formed the primary result for 1064 (85%) of the MS metabolites; in this analysis, the minimum (median) sample size was 93 (260). Of the metabolites tested, 127 (12%) were associated with the intervention (Holm-corrected *p*<0.05) with 72 (57%) showing an increase in response to treatment (**Figure 2** and **Supplementary Table S3)**. The strongest association was seen for a metabolite identified as erythronate (beta: -0.82 (95%confidence interval (CI): -0.99, -0.65), Holm corrected *p*=2.84 × 10^−15^) although the identity of this metabolite has not yet been confirmed by Metabolon based on a standard. An estimated 16%of the variance in erythronate levels at 12-months was explained by group allocation. In most cases, post-hoc checks gave little evidence for between group (control/intervention) differences in metabolite levels at baseline; 1 out of 186 associated metabolites had p<0.05/186 (Wilcoxon rank sum test) (**Supplementary Figures S2A (MS) and S2B (NMR))**.

**Figure 2.**
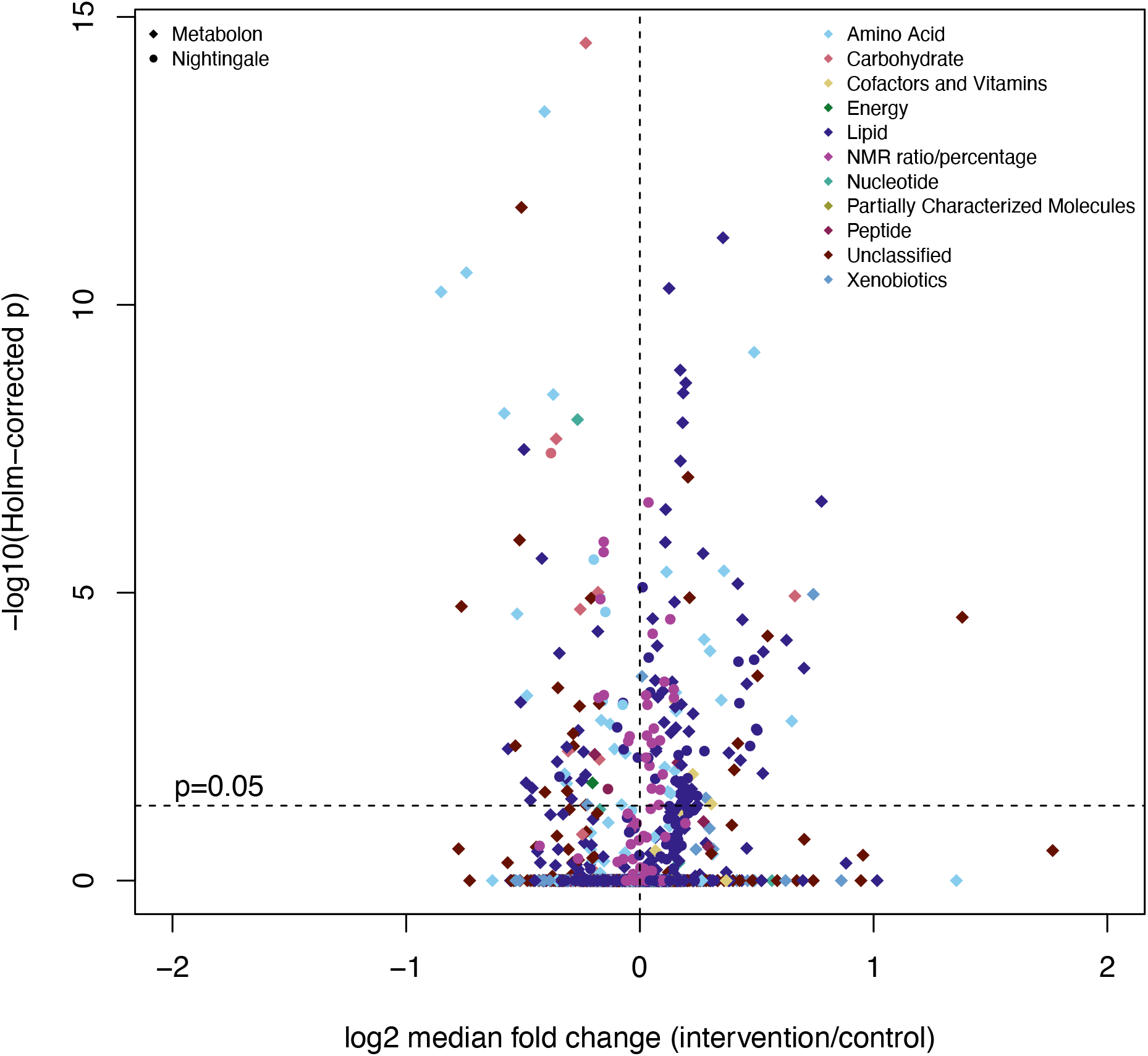
Association of metabolites run in linear model with intervention status. Holm-corrected p-values for allocation effect extracted from linear regression model. Log_2_ fold change calculated using raw (untransformed and unadjusted) metabolite data at T_1_ (12 months). Horizontal dashed line indicates Holm-corrected *p*=0.05.

In the intervention group, we observed a decrease in phosphatidylethanolamines (PEs), branched chain amino acids (BCAA) and related metabolites (i.e., those allocated to the same super-and sub-pathways), sugars (for example, monosaccharides) and in ratio measures that capture the relative abundance of triglycerides (TG) to total lipids within specific lipid fractions (for example, TG to total lipids ratio in small, medium and large low-density lipoproteins (LDL)). In contrast, increases were seen in a subset of lipids including sphingolipids, plasmalogens and metabolites assigned to the ‘fatty acid metabolism (acyl choline)’ sub-pathway and for amino acids from the sub-pathways ‘Glycine, Serine and Threonine Metabolism’ and ‘Urea cycle; Arginine and Proline Metabolism’. There was also evidence (from NMR) for an increase in the intervention group of the proportion of cholesterol and cholesterol esters relative to total lipids in a variety of lipid fractions and an increase in the ratio of polyunsaturated fatty acids to total fatty acids.

### Pathway enrichment analyses

Enrichment analyses were conducted to understand the properties of the metabolite features found to be associated with intervention in the linear models. Analyses were conducted jointly across the NMR and MS datasets and gave evidence for enrichment in the associated metabolites for NMR derived measures (2.4-fold, *p*=5.08 × 10^−06^) and for the carbohydrate super pathway (2.4-fold, *p*=0.011) (**Figure 3**). This indicates metabolites allocated to these groups were overrepresented in the list of associated metabolites.

**Figure 3.**
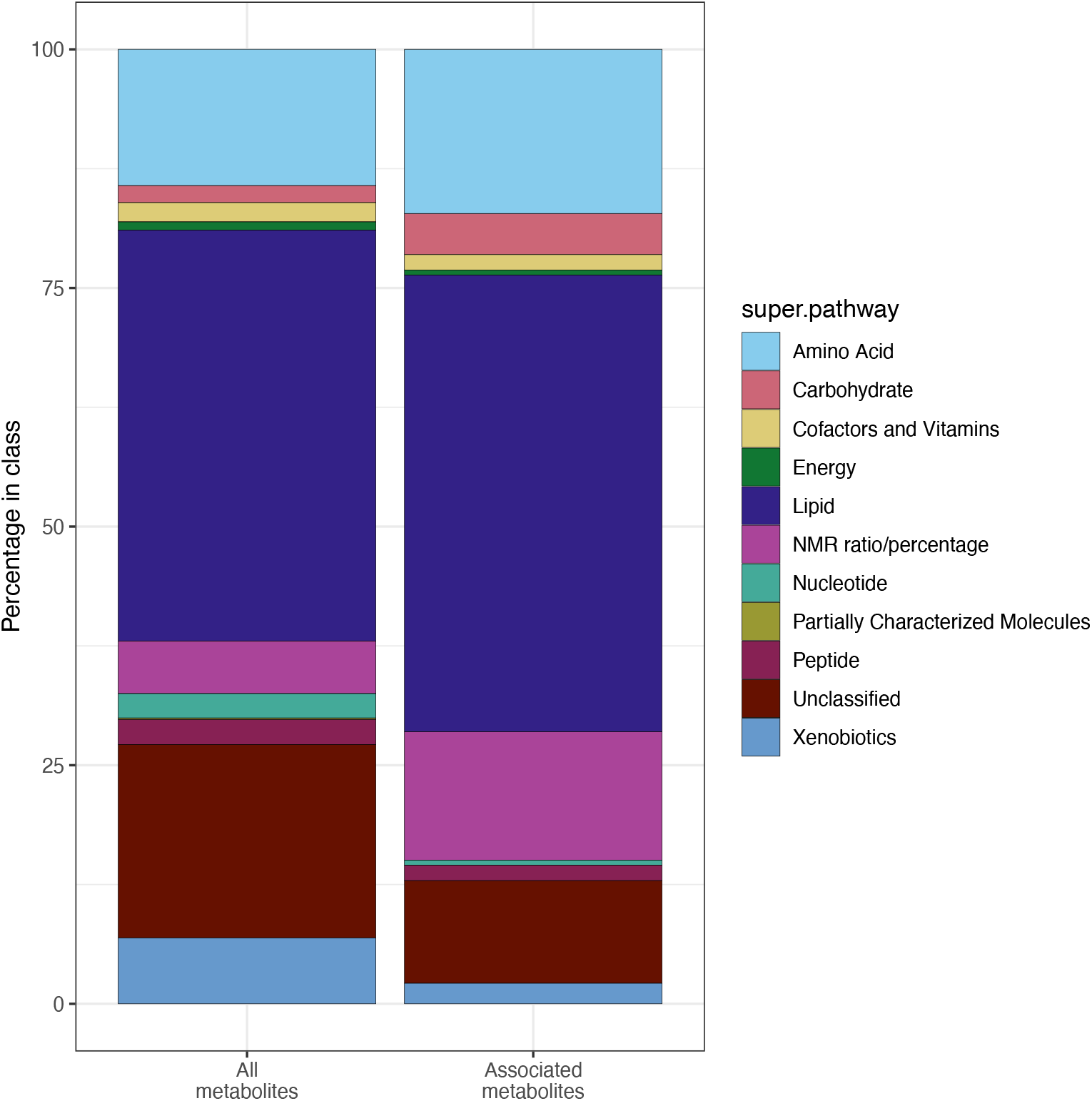
Distribution of metabolites across super pathways. ‘All metabolites’ = all metabolites with <40% missing data (N=1289); ‘Associated metabolites’ = all associated metabolites from linear model (Holm corrected p<0.05) (N=186). Hypergeometric-based enrichment analyses (conducted jointly across the NMR and MS datasets) gave evidence for enrichment in the associated metabolites for NMR derived measures (2.4-fold, *p*=5.08 × 10^−06^) and for the carbohydrate super pathways (2.4-fold, *p*=0.011).

### Exploratory analysis of associated metabolites and clinical phenotypes

An investigation was conducted into the relationship between the metabolites found to be associated with intervention in the linear models and change in a subset of clinical phenotypes selected based on their relevance to the long-term health of patients with type 2 diabetes. First, the set of features associated with the intervention in the linear model was reduced to a subset of approximately independent, representative features. This was done using a hierarchical clustering procedure that allocated the 1,289 metabolites with <40%missing data (225 NMR and 1064 MS) to 238 metabolite clusters with a minimum intra-cluster correlation of 0.2 (**Supplementary Table S4**). The median cluster size was four (range: 1 to 66). Once the list of 186 associated metabolites from linear model analysis was restricted to one representative metabolite per cluster, there were 61 associated representative metabolites - 11 from the NMR platform (including two derived measures) and 50 from the MS platform, of which 10 were classified as ‘unknown’ by Metabolon (i.e., they were not named/annotated) (see **Table 2** for linear regression results for 50 annotated metabolites). Between 20%and 100%of metabolites within clusters containing an associated representative metabolite were nominally associated with intervention (unadjusted p<0.05) with the mean proportion being 72%(**Table 2**). Of the 33 associated representative metabolites from clusters containing more than five metabolites and therefore tested for enrichment, 24 had the same super pathway designation as that for which its cluster showed enrichment (**Table 2**).

**Table 2.**
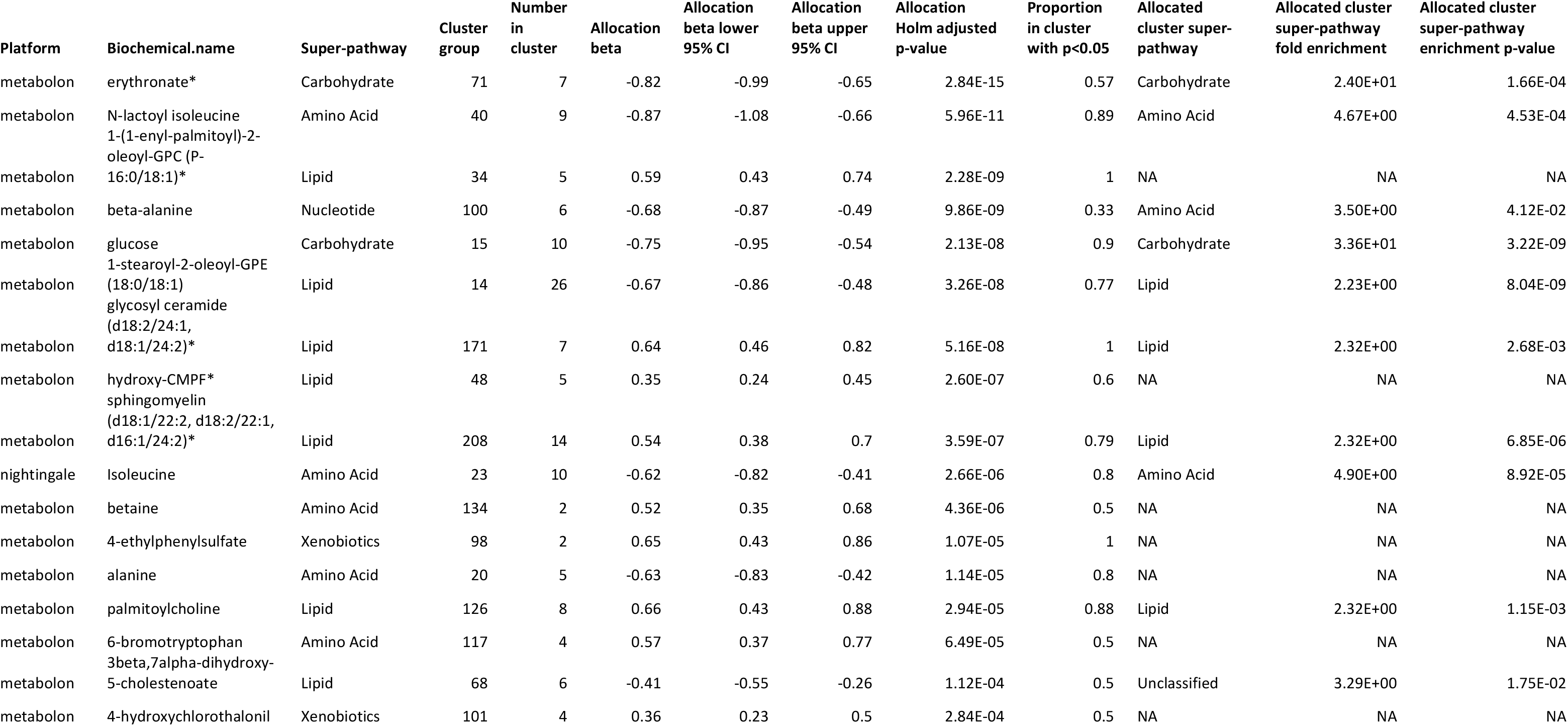
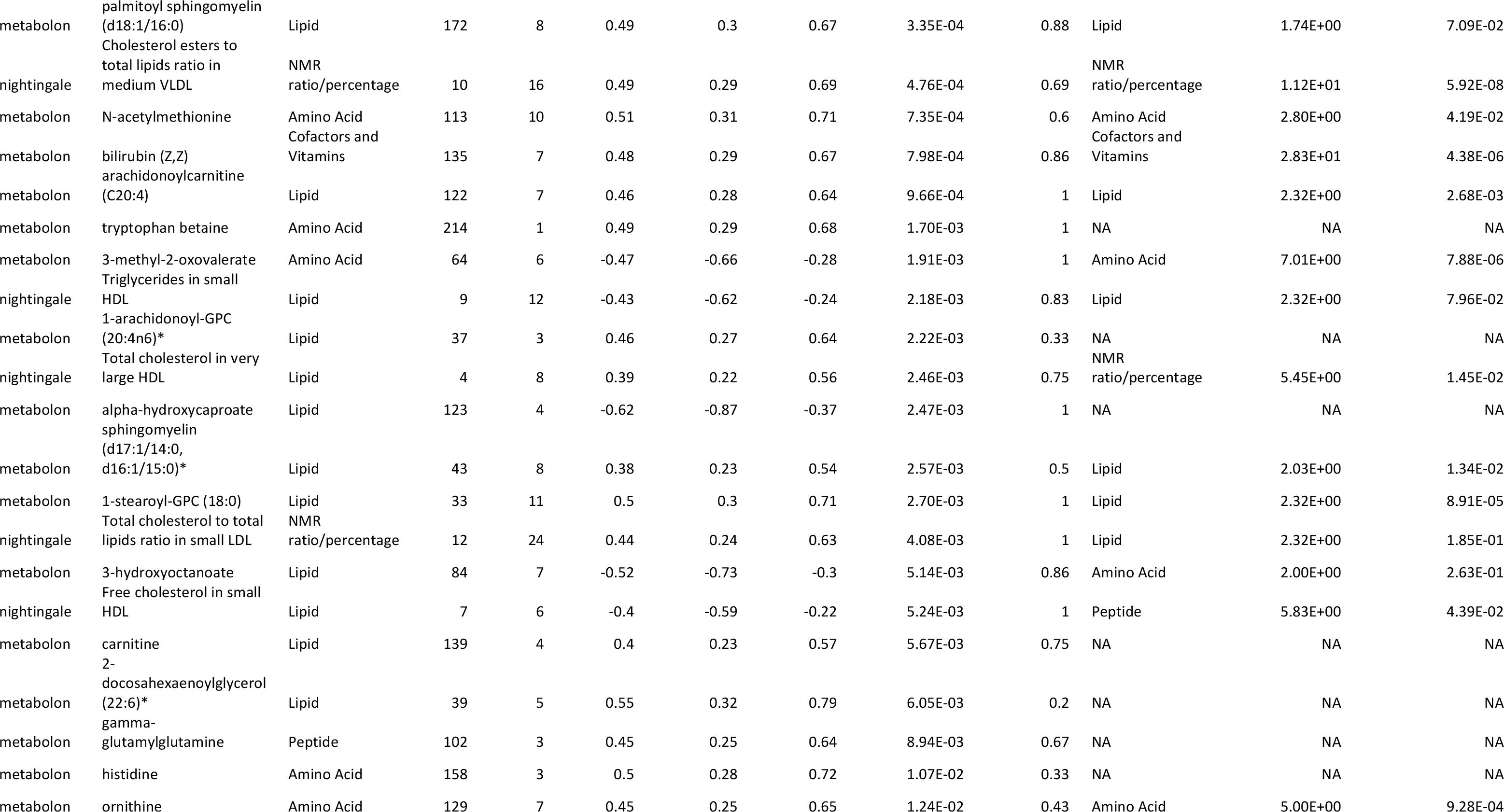
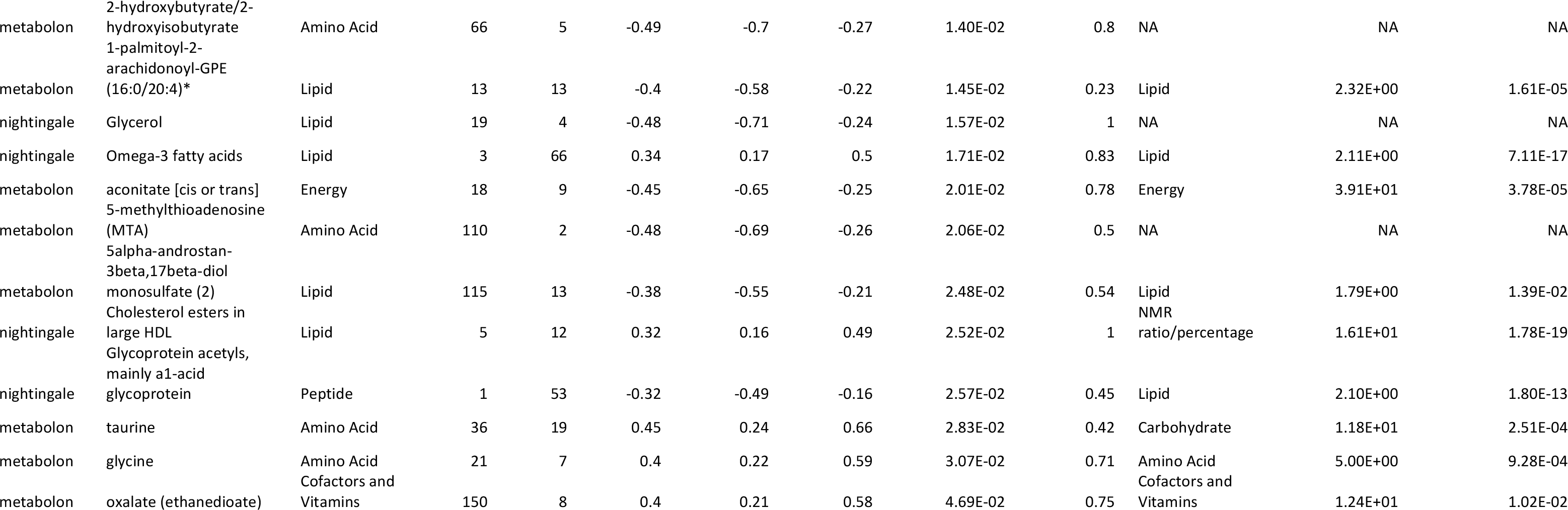
Associated representative metabolites from linear model (restricted to named/annotated features only) Effect estimates shown for the fixed effect ‘allocation’ where control group is considered the reference group with effect estimates therefore representing the difference seen in the intervention group relative to the control group (in normalized standard deviation units). CI = confidence interval; * = indicates a compound that has not been confirmed based on a standard.

Metabolite data at 12 months for the subset of 61 representative metabolites were used as input to a sample based probabilistic principal component analysis (PCA) (20) (see **Supplementary Figure S3** for the PCA scree plot). This analysis shows separation of participants on PC1 (which explained 21%of the variance) according to both their allocation to intervention or control arms of the trial and their remission status at 12 months (**Figure 4A & B**). This pattern can also be seen in a heatmap-based representation of these data whereby most participants who were allocated to the intervention and achieved type 2 diabetes remission were clustered together (**Supplementary Figure S4**). The relationship between the derived (top) PCs and selected clinical phenotypes was assessed by Pearson’s correlation (r) and presented as a biplot. Of the clinical phenotypes considered as indicators of general metabolic health, the strongest correlations with PC1 were observed for change in weight (r=-0.71, 95%CI: -0.76, -0.64), p=4.1 × 10^−41^), change in HbA1c (r=-0.52, 95%CI: - 0.60, -0.44, p=2.2×10^−19^) and change in EQ-5D visual analogue scale (EQ-5D VAS) (r=0.32, 95%CI: 0.20, 0.42, p=1.8 × 10^−07^) (**Figure 4A, Supplementary Table S5A**). The strongest correlations with PC2 (which explained 7%of the variance) were observed for change in total cholesterol (r=-0.31, 95%CI: -0.41, -0.19, p=4.8 × 10^−07^) and change in creatinine (r=-0.13, 95%CI: -0.25, -0.01, p=0.03) (**Figure 4A, Supplementary Table S5A**). Of the clinical phenotypes considered relevant for non-alcoholic fatty liver disease (NAFLD), a moderate negative correlation was observed between PC1 and all change phenotypes, with the strongest correlation seen for change in liver fat percentage (r=-0.62, 95%CI: -0.75, -0.44, p=7.2 × 10^−08^) (**Figure 4B, Supplementary Table S5B**). The contribution of the representative associated metabolites to the PCs is relatively consistent with their pattern of association with the intervention, such that the metabolites most strongly associated with the intervention, for example, glucose and isoleucine, have relatively large loadings for PC1 (**Figure 4C**). Two-way boxplots of raw (untransformed and unadjusted) metabolite data for the 10 metabolites with the greatest loadings show the distribution of metabolite change by weight change tertiles and remission status at 12 months (**Figure 5** and **Supplementary Figure S5**). For most features, there is evidence for mean differences both across weight change categories and between those of differing remission status within the same weight change tertile.

**Figure 4.**
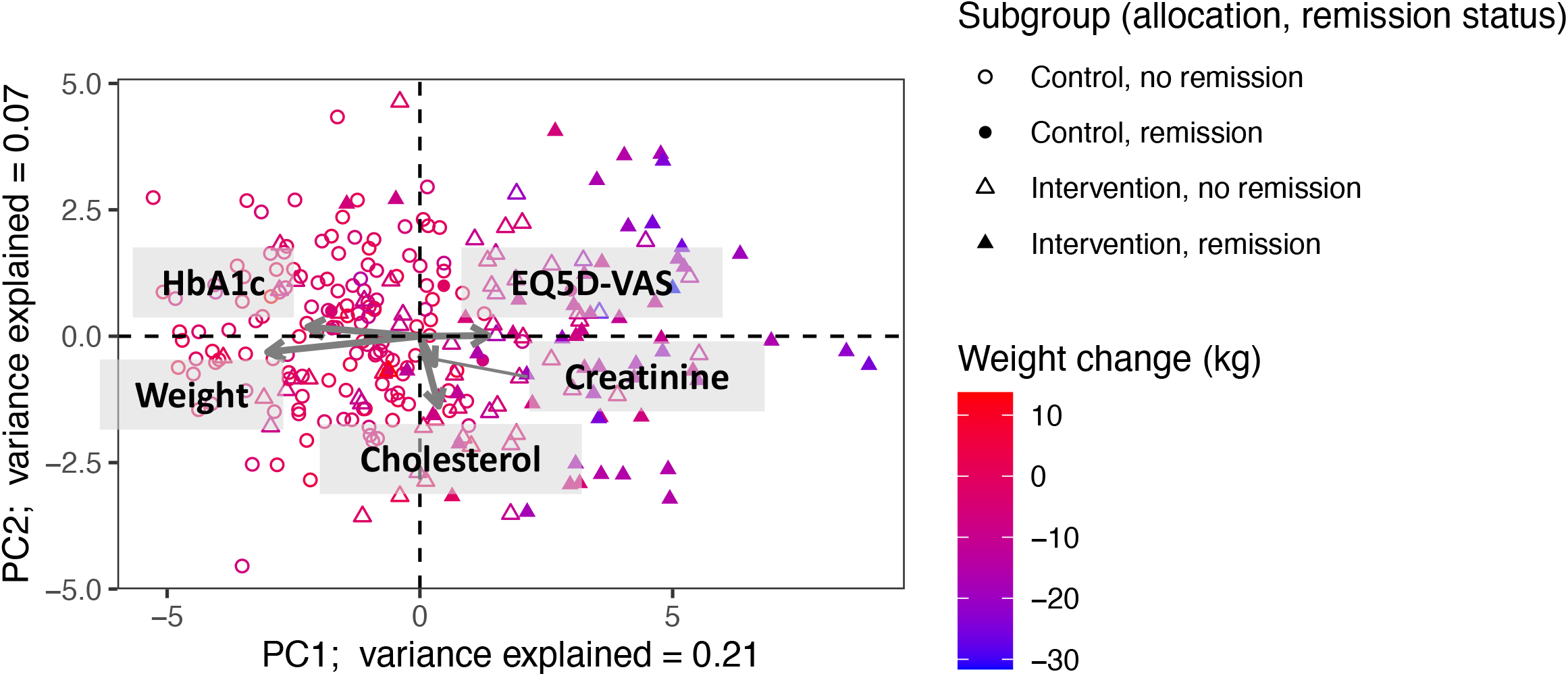
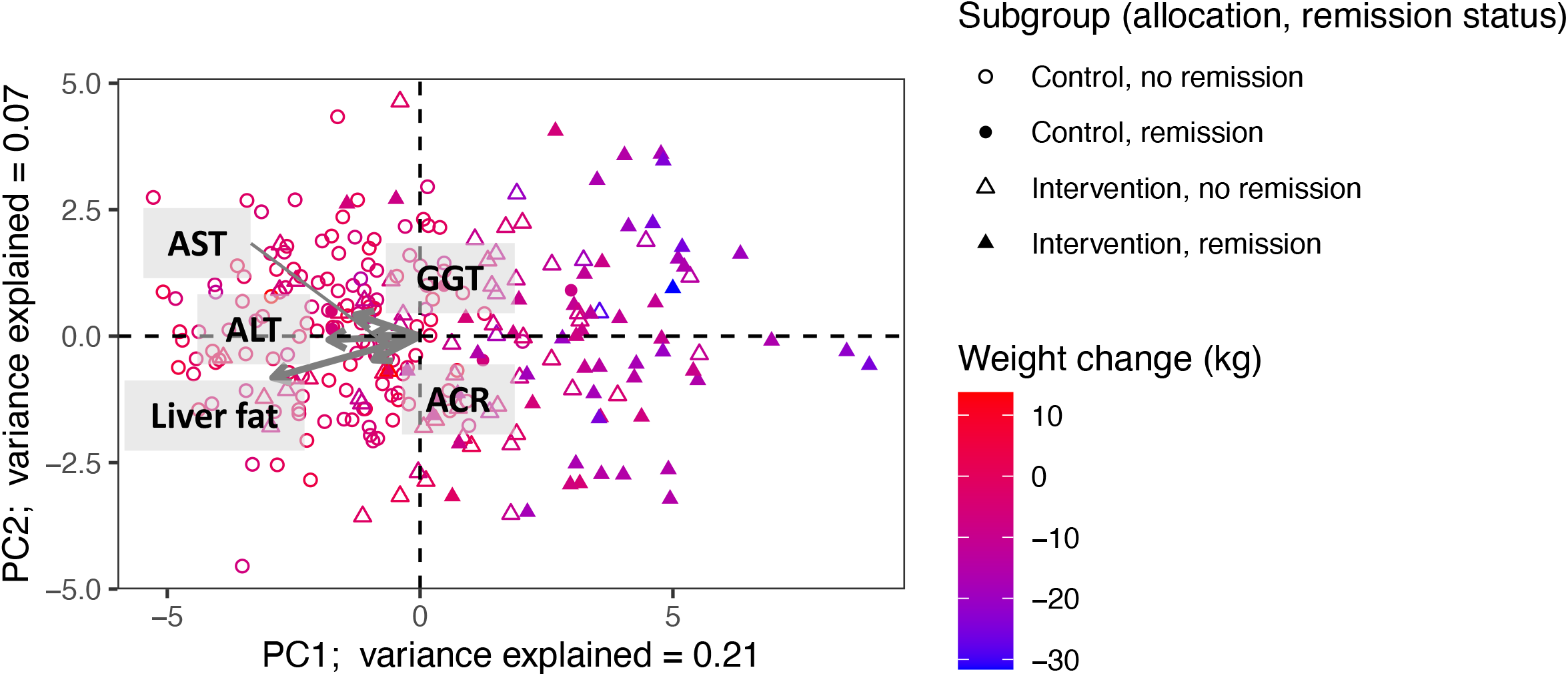
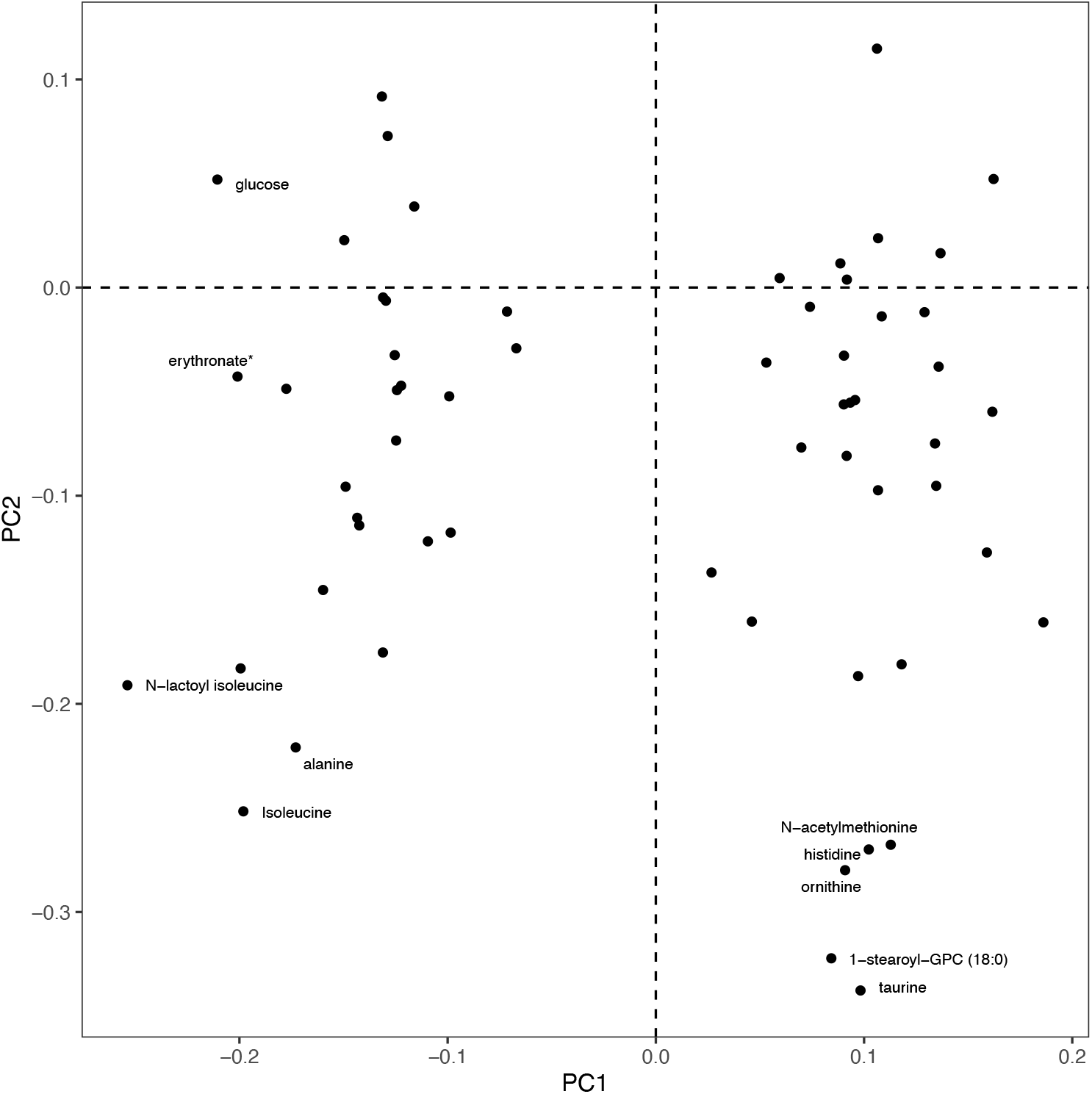
Principal component analysis (PCA) on samples using data for 61 associated representative metabolites after adjustment for covariates. Further description of plot generation in **Supplementary Methods**. Change refers to the difference in measurements from the baseline timepoint to the 12-month timepoint. Weight change expressed in kilograms. (4A) Biplot to show relationship of PC1 and PC2 with change in a set of indicators of general metabolic health found to be correlated with PC1 and/or PC2 (p<0.05): Cholesterol = total cholesterol (mmol/l); Creatinine (umol/l); EQ5D-VAS = quality of life as assessed by EQ-5D visual analogue scale; HbA1c = glycated haemoglobin (mmol/mol). (4B) Biplot to show relationship of PC1 and PC2 with change in a set of parameters relevant for non-alcoholic fatty liver disease (NAFLD) found to be correlated with PC1 and/or PC2 (p<0.05): AST = aspartate aminotransferase (units/l), ALT = alanine aminotransferase (units/l), ACR = albumin-to-creatinine ratio (mg/mmol), GGT = gamma-glutamyl transpeptidase (units/L); Liver fat = liver fat percentage. (4C) PCA loadings plot to show the contribution of selected metabolites to the PCs. Labels added to identify variables with loadings > 0.2 or <-0.2 for one or both PCs.

**Figure 5.**
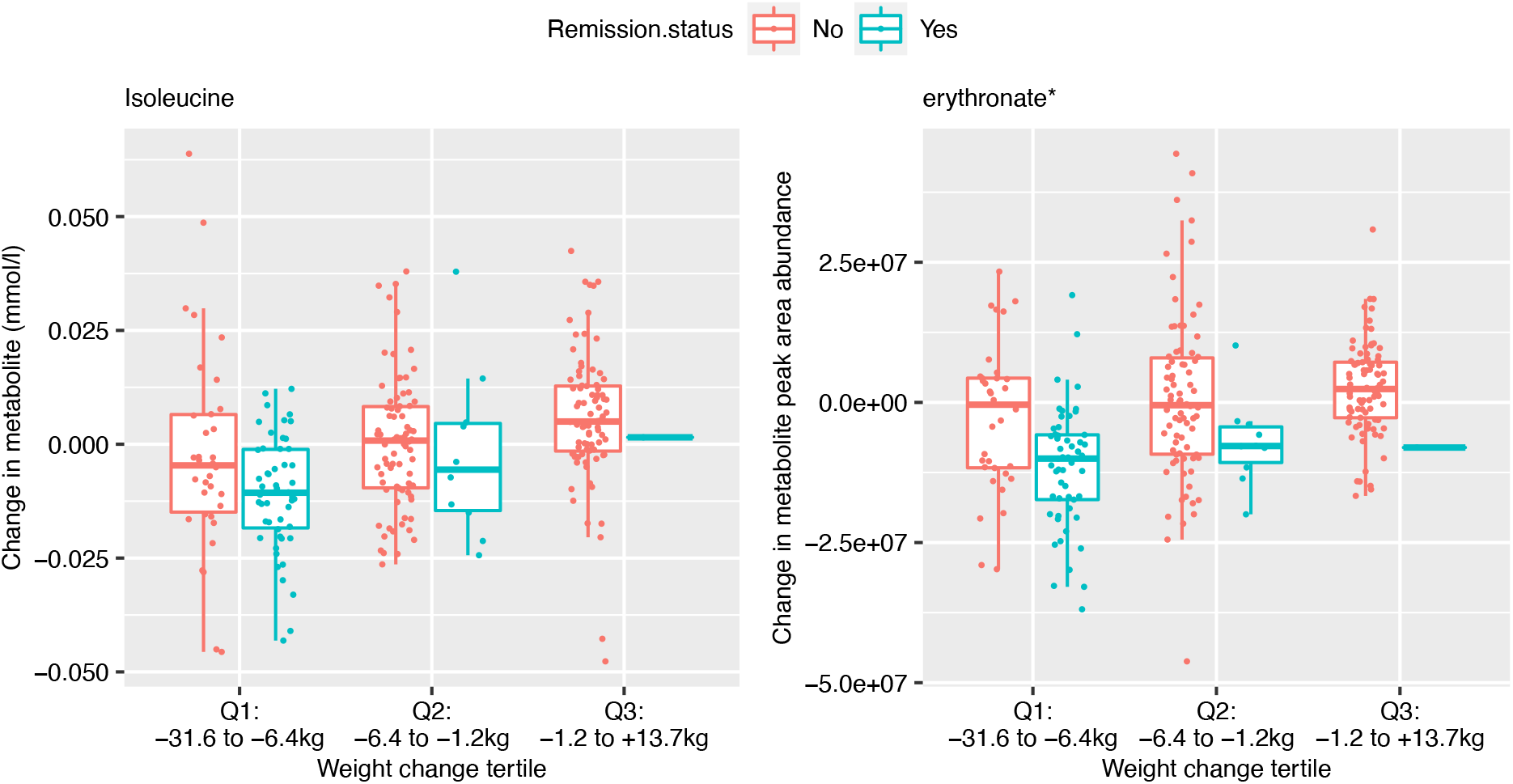
Distribution of metabolite change by weight change tertiles and remission status at 12 months. Metabolite change (the difference between the value at 12 months and that at baseline) was calculated using raw (untransformed and unadjusted) metabolite data. Data shown for isoleucine (left, source: NMR) and erythronate (right, source: MS). Similar plots for the remaining eight metabolites labelled on **Figure 4c** can be found in **Supplementary Figure S5**.

### Effect of intervention on metabolites – logistic regression model

Results from the logistic model (based on the PA dataset and able to detect and describe the presence of xenobiotics) formed the primary result for 190 of the MS metabolites, the majority of which were classified as xenobiotics or unidentified molecules (**Supplementary Table S6**). Of these, 19 were associated with the intervention (*p* < 0.05) with 11 showing depletion in the intervention group; 12 out of the 19 were named/annotated metabolites (**Table 3**). Metformin showed the strongest association and was present in 26%(31/117) of 12-month samples from those in the intervention group compared to 78%(112/144) in the control group. Concordance between metformin use as indicated by General Practitioner (GP) records and the presence/absence of metformin in blood samples was high both at baseline and 12 months with at most 15 discrepant cases out of N=261 (i.e., cases where metformin was detected in samples from patients whose records indicated no prescription and vice versa).

**Table 3.**
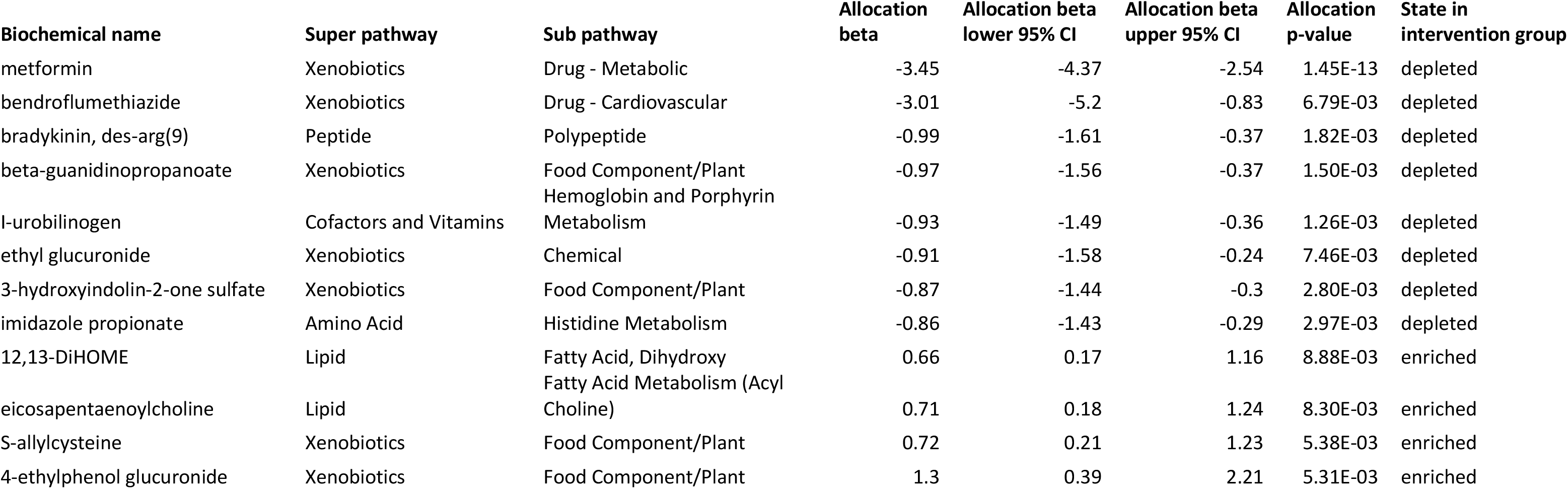
Associated metabolites from logistic model (restricted to named/annotated features only) Effect estimates shown for the fixed effect ‘allocation’ where control group is considered the reference group with effect estimates therefore representing the average change in log odds in the intervention group relative to the control group. CI = confidence interval.

## Discussion

In this study, we explored the metabolic consequences of a calorie restricted dietary-based intervention aimed at achieving and sustaining type 2 diabetes remission in patients attending primary care practices. We observed a broad metabolomic signature associated with the intervention with differences observed in metabolites from every one of the pre-designated biochemical classes (or super pathways). Under a conservative correction for multiple testing, 26%of NMR-derived metabolites and 12%of Metabolon-derived metabolites were altered by the intervention, suggesting that metabolite derangements are modifiable. We report a lipid pattern change with reduction in TG rich lipoproteins across the lipoprotein cascade, but enrichment of (lyso)plasmalogens and reversal of amino acid changes predictive of diabetes, as well as a reduction in a range of sugars, including but not limited to glucose. These changes suggest that the previously described perturbations of metabolite profile in people with type 2 diabetes are largely reversible with intentional weight loss. This also suggests that weight change is upstream of many downstream metabolite alterations.

### Characteristic changes in metabolite profile

Many of the strongest signals observed in these data mirror those described in existing literature describing the metabolic footprint of type 2 diabetes (i.e., associate with opposite directions of effect). For example, we see decreased concentrations of isoleucine, leucine and valine following dietary intervention where plasma concentrations of these BCAAs are frequently elevated in type 2 diabetes (5) and have been shown to be predictive of future type 2 diabetes both in prospective studies of disease (21) and based on genetic liability to disease (10). By using an untargeted metabolome-wide approach we were able to further characterize the plasticity of this highly relevant network. We observed concurrent decreases in a number of gamma-glutamyl BCAA dipeptides allocated to the same cluster as the BCAAs themselves; gamma-glutamyl AAs are produced when the enzyme, gamma-glutamyl transpeptidase (GGT), present mainly in the liver, catalyzes the transfer of the gamma-glutamyl moiety of glutathione to an amino acid (22). In addition, alanine, not a BCAA but a similarly hydrophobic AA involved in BCAA metabolism (23), was decreased in the intervention group. Given genetic evidence for a causal influence of elevated BMI on increased levels of BCAA (24) one might infer from our results that the weight loss caused a subsequent drop in the levels of BCAA. However, levels of circulating BCAA are influenced by dietary intake (25) and therefore, the decreases we see could also be the result of sustained changes to diet within the intervention group. Furthermore, it is possible that some of the improvements to metabolic health following the intervention (including weight loss itself) could have been driven by reductions in BCAA intake (26), with evidence emerging that specific reductions in dietary isoleucine in particular may contribute to improved metabolic health (27). Future work using additional samples collected in a subset of participants immediately after the total diet replacement phase (12 weeks) and during the weight maintenance phase of the trial (from 6 months) may help to differentiate metabolic responses to dieting from those that result from weight loss itself.

Similarly to BCAAs, the reduction we see in the levels of several simple sugars, including the monosaccharides fructose, glucose and mannose, are opposite to the elevations seen in these metabolites in the presence of obesity. Whilst structurally similar, the predominant dietary sources, metabolic pathways and biological effects of these simple sugars are quite different though interdependent (28). Mannose specifically has been associated not only with insulin resistance but also with increased risk of several chronic diseases including type 2 diabetes and cardiovascular disease (29). Whilst results of a recent mannose supplementation study suggest associations between mannose and adiposity are unlikely to be causal (30), experiments involving a mouse model of diet-induced obesity suggest mannose supplementation early in life can prevent weight gain via alterations to gut microbiota (31). Regardless, mannose is a useful biomarker of insulin resistance and its complications and therefore the reduction we see following the Counterweight Plus intervention supports the notion of downstream benefits to health.

Patients in the intervention arm saw increases in several lipids previously associated with a favourable metabolic profile. Specifically, increases were seen in concentrations of several (lyso)plasmalogens, a special class of phospholipids characterized by the presence of a vinyl–ether bond at the *sn*-1 position. The potential for this class of phospholipids to act as endogenous antioxidants has been recognised (32) with plasmalogens thought to be preferentially oxidized thus protecting other lipid species from oxidation (33). In a cross-sectional study of participants with overweight and obesity, plasmalogen levels were found to be inversely correlated with body fat percentage but seemingly not related to body mass index (BMI) or waist-to-hip ratio (WHR) (34). The lack of association in these more commonly used indicators of adiposity may be related to their sub-optimal performance as proxies for adiposity in this relatively small sample of individuals all with BMI>25 kg/m^2^ (n=65). Alternatively, this may point towards a more complex interplay between metabolic health and plasmalogens, an argument supported by results of a recent study comparing the effects on the serum lipidome of different types of bariatric surgery. In this study of 37 patients with morbid obesity who underwent either sleeve gastrectomy (n=25) or biliopancreatic diversion (n=12), plasmalogen levels were only seen to increase in the sleeve gastrectomy treated patients. However, both the small overall sample size and the imbalance in the size of the study arms (and therefore corresponding power) limits the robustness of these findings.

### Detecting associations with changes in exogenous factors

Whilst changes to the metabolism can be expected in response to the intervention-induced weight loss experienced by many of those in the intervention group, we also expect the intervention to have resulted in the adoption of new dietary patterns and a change in medication regimes. In support of this, the logistic regression analysis here revealed between group differences in the frequency of detection of both potential dietary biomarkers and medications. For example, S-allylcysteine, a biomarker for garlic consumption (35, 36) was enriched in the intervention group whilst ethyl glucuronide, a validated urine biomarker for alcohol consumption (37, 38) was depleted. The logistic regression also suggests an apparent depletion of imidazole proprionate. Levels of this microbially-produced metabolite have been shown to be elevated in subjects with incident type 2 diabetes (39, 40). More recently, pre-treatment of mice with imidazole proprionate has been shown to have an inhibitory effect on metformin action (41). This provides evidence for sustained dietary changes at the end of the weight loss maintenance phase of the intervention. Meanwhile the reduction we see in the presence of metformin in the intervention group at 12 months provides a useful positive control as well as offering an opportunity to verify medication usage and as such address the potential bias of compliance in future clinical trials.

### Metabolite profile variation and clinically relevant biomarkers

As previously reported, the adjusted mean difference in bodyweight at 12 months in the intervention group as compared to the control group was -8.8 kg (19) and many of the changes we see in metabolite levels likely reflect this difference. The exploratory PCA analysis points towards weight change over the course of the study as a key factor relating to the metabolic signature of the intervention with a correlation between weight change and PC1 (which explained 21%of the variance) of -0.71. The changes we see in levels of glucose and BCAA are characteristic of those seen with weight change in other settings (24, 42, 43). The shift we see in the amount and distribution of TG and cholesterol as captured by the NMR metabolomics platform points towards alterations in the metabolism of TG-rich lipoproteins. In general, we see a decrease in the TG to total lipids ratio across LDLs and VLDLs in patients in the intervention group with what appears to be a corresponding increase in the total cholesterol and/or cholesterol ester (CE) to total lipids ratio in a similar subset of lipoproteins as would be expected given the previously characterised decrease in hepatic production of VLDL TG (44). These effects are in keeping with the proposed mechanism by which excess TG in the circulation triggers the transfer of TGs from the core of TG-rich lipoproteins to LDL in exchange for CEs by the CE transfer protein (CETP) (45). The pattern of association we see in response to the intervention is approximately opposite that associated with type 2 diabetes development in a prospective analysis using the same NMR platform as used here (21).

The moderate correlations we see between PC1 and liver fat percentage and clinical indicators of liver health (e.g., aspartate aminotransferase (AST), alanine aminotransferase (ALT)) in the exploratory PCA analysis, together with the remission substructure, speaks to the ability of this metabolome wide approach to detect biological signatures of the effect of weight loss on liver health previously evidenced in DiRECT (44). Studies have shown the effect of low energy diets on the mobilization of liver fat to be almost instantaneous with dramatic decreases in liver fat seen within seven days as a result of the negative calorie balance and need to mobilize energy stores (46). By conducting an in-depth analysis of metabolites in the presence of sustained improvements to liver health as here, we can further investigate proposed biological systems, such as the twin cycle hypothesis (47), including in the context of variable patient response. We see an increase in the intervention group of several metabolites allocated to the sub-pathway ‘fatty acid metabolism (acyl choline)’. The liver represents an important location for choline metabolism and low dietary intake of choline has been associated with fatty liver and liver damage (48). The effect here is concordant with that seen by Al-Sulaiti *et al*. (49) in a comparison between equally obese insulin sensitive and insulin resistant individuals, that showed phospholipid metabolites including choline, glycerophosphoethanolamine and glycerophosphorylcholine (GCP) were present in lower concentrations in those with an insulin resistant phenotype. While choline is not normally an essential nutrient for healthy adults, deficiency can develop with severe illness and folate and methionine deficiencies, or in premature infants. In our analysis, whilst the evidence for association between intervention and choline was not strong, the estimated direction of effect was consistent (beta: 0.28, 95%CI: 0.06, 0.50, unadjusted *p*=0.01) and GCP was in the list of associated metabolites (beta: 0.65, 95 %CI: 0.45, 0.86), Holm corrected *p*=2.09 × 10^−6^). The hierarchical clustering saw choline placed in a cluster alongside taurine, one of the representative associated metabolites that contributed to the PCA. Together with the appearance in the list of associated metabolites of several other molecules allocated to the ‘glycine, serine and threonine metabolism’ sub-pathway, these results suggest that this constellation of metabolites may be relevant to recovery of liver and pancreatic cell function.

### Strengths and limitations

A major strength of this study is the use of samples and clinical data collected from a relatively large (as compared to existing literature) cluster randomized trial with a well-matched control arm. That we were able to measure metabolites both at baseline and 12 months added to the robustness of the analysis whilst the use of two complementary metabolomics platforms increased the overall coverage of the metabolome. However, metabolomic data were not available for all 298 patients enrolled in the trial. In particular, 12-month samples were not available from patients who dropped out of the trial prematurely, the majority of whom were from the intervention arm of the trial. This differential missingness has the potential to bias effect estimates, however, given the small number of patients who dropped out and the fact that primary analyses were restricted to patients with both baseline and 12-month measures, we believe the impact of this on our results is minimal. Whilst our study design enabled us to conduct a comprehensive evaluation of the metabolomic impact of the Counterweight Plus intervention overall, it is challenging to attribute those changes to specific elements of the intervention – for example, to weight loss or changes to dietary patterns. Also, further work is needed to dissect the temporal order of effects, for example, the extent to which the metabolite changes we observe occur before or after improvements in clinical markers such as insulin sensitivity and GGT. Finally, whilst the hierarchical clustering approach used here enabled us to look beyond single-metabolite associations, more sophisticated network approaches could provide further insights into biological pathways relevant to type 2 diabetes and related metabolic disorders.

## Conclusion

In this study, we have effectively characterized the impact of Counterweight-Plus, a weight management programme designed to be delivered in primary care, at the level of the metabolome. The potential for this intervention to deliver meaningful changes in the health of patients with type 2 diabetes has already been demonstrated and to this we add evidence for widespread improvements in a range of lipid and sugar moieties and reductions in BCAAs in a pattern directly opposite to documented perturbances in type 2 diabetes. These results suggest either excess weight and/or its upstream dietary drivers, are largely responsible for such perturbances. Whether these data contain important information relating to the interindividual differences in efficacy of the intervention, both in terms of weight loss and type 2 diabetes remission, will be the focus of future work. Through analyses such as these, molecular phenotyping can be used alongside high quality trials data to better understand the physiological impact of proposed interventions.

## Methods

### Study design and participants

DiRECT is a two-year open-label, cluster-randomised controlled trial and is registered with the ISRCTN registry, number 03267836. Ethics approval was granted by West 3 Ethics Committee in January, 2014, with approvals by the National Health Service (NHS) health board areas in Scotland and clinical commissioning groups in Tyneside. The trial was carried out to assess whether effective weight management, delivered in a primary care setting, could produce sustained remission of type 2 diabetes. The protocol, including details of recruitment methods, study conduct, and planned analyses, has been published elsewhere (50) as have the baseline characteristics of the groups (18). In brief, the trial was conducted at 49 primary care practices in Scotland and the Tyneside region of England between 25th July 2014 and 5th August 2016. Practices were randomly assigned (1:1) to provide either a weight management programme (intervention) or best-practice care by guidelines (control), with stratification for study site (Tyneside or Scotland) and practice list size (>5700 or ≤5700). Participants, carers, and research assistants who collected outcome data were aware of group allocation. Individuals aged 20–65 years who had been diagnosed with type 2 diabetes within the past 6 years, had a body-mass index of 27–45 kg/m^2^, and were not receiving insulin were recruited. The intervention (Counterweight-Plus) comprised withdrawal of antidiabetic and antihypertensive drugs, total diet replacement (TDR) (825– 853 kcal/day formula diet for 3–5 months), stepped food reintroduction (2–8 weeks), and structured support for long-term weight loss maintenance. All participants provided written informed consent.

The trial was conducted over a two-year period with principal data collection points scheduled at baseline, 12 months, and two years. Bloods were collected and a range of clinically relevant outcomes measured, including for example, liver function tests, serum total cholesterol, HDL cholesterol and triglycerides. In addition, serum samples were stored for future research. In this study, we analysed samples from the baseline and 12-month time points using both an untargeted mass spectrometry (MS) approach (Metabolon, Inc.) and ^1^H-nuclear magnetic resonance (^1^H-NMR) spectroscopy (Nightingale Health). For all other data used in our analyses, we used the same version of the trial database as used for the main trial analysis at 12 months, as described and analysed in Lean *et al*. (2017) (19). These data comprised an intention-to-treat population of 149 participants per group (total n=298) with baseline characteristics similar between groups (as published previously, Table 1 in Lean *et al*. (19)). At 12 months, mean bodyweight had fallen by 10·0 kg in the intervention group and by 1·0 kg in the control group (adjusted difference: –8·8 kg, 95%confidence interval –10·3 to –7·3; p<0·0001) and almost half of the 149 participants in the trial arm (68 or 46%) had achieved type 2 diabetes remission (as defined in the trial protocol) (19).

### Sample collection

Patients were asked to fast overnight before the blood draw. Blood for the serum samples was drawn into an SST Serum separator tube then kept at 4°C until being processed. Within five hours of blood draw samples were centrifuged at 2000g for 15 minutes at 4°C then the serum was separated and stored as five 0.5 mL aliquots at -80°C. Samples collected in Scotland were transported as whole blood at 4°C to the Central Glasgow laboratory (BHF GCRC, University of Glasgow) and processed there, whilst samples collected in the Newcastle area were processed locally and stored as serum at -80°C before transfer to Glasgow. In total, 574 serum samples collected from 302 unique individuals during the trial were sent for metabolomic analysis. Aliquots run on the two different metabolomics platforms were extracted from the same sample source. Samples were sent first to Metabolon, Inc. (Durham, North Carolina, USA) where they were thawed for the first time for aliquoting. Remaining sample material was then re-frozen and sent to the MRC IEU Metabolomics Facility, University of Bristol, for NMR analyses (after one further thaw). All analysts were blind to intervention/control status.

### Metabolite data acquisition

#### ^1^H-NMR metabolomics

A serum ^1^H-NMR metabolomics platform (Nightingale Health Ltd, Helsinki, Finland) was used to quantify circulating metabolites with an emphasis on lipid or lipoprotein lipid measures (51). Details of the experimentation have been described elsewhere (51, 52). This high-throughput metabolomics platform provides simultaneous quantification of routine lipids, lipid concentrations of 14 lipoprotein subclasses and major subfractions, and further abundant fatty acids, amino acids, ketone bodies, and gluconeogenesis-related metabolites in absolute concentration units. The measured variables include 148 primary measures quantified in absolute concentrations as well as 79 additional derived measures such as ratios and percentages, primarily related to fatty acids and lipoprotein composition. Herein, this dataset is referred to as ‘NMR data’ and the 79 additional measures specifically referred to as ‘derived measures.’

#### Mass spectroscopy data

An untargeted metabolomics analysis of over 1,000 metabolites was performed at Metabolon, Inc. (Durham, North Carolina, USA) using established protocols. The Metabolon analysis consisted of four independent ultra-high-performance liquid chromatography-tandem mass spectrometry (UPLC-MS/MS) runs. All methods utilized a Waters ACQUITY ultra-performance liquid chromatography (UPLC) and a Thermo Scientific Q-Exactive high resolution/accurate mass spectrometer interfaced with a heated electrospray ionization (HESI-II) source and Orbitrap mass analyzer operated at 35,000 mass resolution. Raw data were extracted, peak-identified and quality control (QC) processed using Metabolon’s hardware and software. Compounds were identified by comparison to library entries of purified standards or recurrent unknown entities with more than 3300 commercially available purified standard compounds. Further details can be found in the **Supplementary Methods** and in published work (53, 54). Baseline and 12-month samples from the same individuals were analysed in the same batch to avoid the confounding of technical variation (batch effects) with time point. The resulting dataset comprised a total of 1,276 metabolite features comprising 959 compounds of known identity (named biochemicals with the majority matched to purified standards) and 317 compounds of unknown structural identity (unnamed biochemicals) as of February 2018 when data were generated (subsequent library updates are described in **Supplementary Methods**). Herein, this dataset is referred to as ‘MS data’

### Metabolite data preparation

Both metabolite measurement approaches were conducted with stringent quality control based on standard workflows implemented by the commercial laboratories. Additional checks were carried out locally to evaluate overall sample quality and to ensure the appropriateness of data for the intended analyses. A pre-release version of the R package *metaboprep* (55) was used; full details of the procedures implemented are in **Supplementary Methods** and the HTML format data summaries produced included as **Supplementary Documents 2 (MS data) & 3 (NMR data)**. In brief, samples were excluded based on high levels of missing (unquantified) metabolite data (>20%) and if they were deemed to be outliers following a probabilistic principal component analysis (PCA) and, in the case of MS data, if samples were deemed to be outliers based on the total peak area (TPA) statistic. Metabolites were excluded only based on high levels of missingness across samples (>20%missing (unquantified) data in NMR data and <5 observations in MS data). Following the application of these filters to the data, the NMR data comprised 567 samples and 225 metabolic features (147 primary measures and 78 derived measures) and the MS data comprised 571 samples and 1254 metabolites (**Supplementary Table S1**). The metabolite data were then merged with that contained within the trial database (N=298 individuals, 149 intervention and 149 control). Data were restricted to include only those individuals present in the trial database and with both a baseline (T_0_) and 12-month follow-up (T_1_) sample present in the filtered metabolite data. This left 258 individuals in the NMR data (115 intervention and 143 control) and 261 individuals in the MS data (117 intervention and 144 control) available for statistical analysis.

Due to the characteristics of metabolite data derived using both NMR and MS approaches, some pre-processing was performed. Two processed datasets were derived: (1) RNT dataset: metabolite data were transformed (across individuals within timepoint) using a rank based inverse normal transformation (where tied ranks were split by assigning a random order); (2) PA dataset – metabolite data were transformed to a presence/absence phenotype with missing values replaced with 0 and non-missing values with 1.

### Statistical analysis

An overview of the statistical analysis is shown in **Figure 1**. We analysed all available data according to group allocation and as such, the result from this analysis represents the intervention effect in a clinical setting (i.e., accounting for non-compliance). In all analyses, the control group is considered the reference group with effect estimates therefore representing the difference seen in the intervention group relative to the control group.

#### Linear regression model

In our evaluation of the effect of the intervention on metabolite levels, outcomes (metabolite levels at T_1_) were compared between groups with linear regression models applied to the RNT dataset. Where metabolite_T0_ or metabolite_T1_ was missing (unquantified) for an individual, that individual was excluded from the analysis of that specific metabolite and therefore, the analysed sample size varied across metabolites. Models were adjusted for the minimisation variables (study centre and practice list size) along with the baseline measurement of the outcome (metabolite at T_0_), age and sex, all fitted as fixed effects:

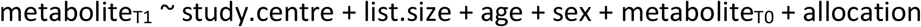

Where metabolite_T1_ is metabolite level at 12 months, study.centre is a binary variable (Tyneside or Scotland), list.size is a binary variable indicating practice list size (<= 5700 or >5700), age is patient age at baseline (years), sex is a binary variable (male/female), metabolite_T0_ is metabolite level at baseline and allocation is a binary variable indicating the individual’s treatment group (control or intervention). Model betas represent the estimated change in metabolite level expressed as normalized SD units per unit change in dependent variable (in the case of allocation this is the mean difference between groups). This linear modelling approach reflects that used to analyse secondary outcomes in the trial itself (19).

Results from this analysis were considered the primary result for all metabolite features with less than 40%missing (unquantified) data at the 12-month timepoint. This missingness threshold was selected based on the increase in the standard error of the treatment group effect estimated from the linear model when the number of observations fell below this level (see **Supplementary Figure S1**). Finally, the Holm (56) method was used to adjust p-values for multiple testing and an adjusted p-value of <0.05 considered as evidence for association. The variance in the metabolite levels (metabolite_T1_) explained by each of the fixed effects was also derived from the model.

#### Pathway enrichment analyses

Hypergeometric-based enrichment analyses were conducted to evaluate the enrichment of classes in the subset of associated features derived from the linear model as compared to all features that were tested by the same model. Metabolite super pathway designation provided by Metabolon was used for the enrichment analysis, with NMR-derived metabolites allocated to super pathways following the approach of Wahl *et al*. (2015) (42).

#### Exploratory analysis of associated metabolites and clinical phenotypes

An investigation was conducted into the relationship between the metabolites found to be associated with the intervention and change (calculated as the difference between the value at 12 months and that at baseline) in a subset of clinical phenotypes selected based on their relevance to the long-term health of patients with type 2 diabetes. First, a hierarchical clustering approach was implemented to reduce the large number of intervention-associated metabolites to a smaller set of representative features that captured the main signals of association (for full details see **Supplementary Methods**). Metabolite data at 12 months for the subset of representative metabolites (adjusted for the same covariates as fitted in the linear model used for the primary analysis) were then used as input to a sample based probabilistic PCA (20). The relationship between the derived (top) PCs and a selection of clinical phenotypes collected as part of the main trial protocol was assessed by Pearson’s correlation (r) and presented as a biplot. The clinical phenotypes included: (1) phenotypes selected for their relevance to the trial’s primary outcomes: allocation (control/intervention) and type 2 diabetes remission status at 12 months where remission is defined as glycated haemoglobin (HbA1_c_) less than 6.5%(<48 mmol/mol) after at least 2 months off all antidiabetic medications (57); (2) a set of indicators of general metabolic health: weight (kg), glycated haemoglobin (HbA1c) (mmol/mol), diastolic and systolic blood pressure, total cholesterol (mmol/l), creatinine (umol/l), c-reactive protein (CRP) (mg/l) and quality of life as assessed by EQ-5D visual analogue scale (VAS) and utility index values; and (3) parameters relevant for non-alcoholic fatty liver disease (NAFLD), a common complication of type 2 diabetes: aspartate aminotransferase (AST) (units/l), alanine aminotransferase (ALT) (units/l), albumin-to-creatinine ratio (ACR) (mg/mmol), gamma-glutamyl transpeptidase (GGT) (units/L) and liver fat percentage.

#### Logistic regression model

A logistic model was applied to the PA datasets to compare the presence of each metabolite by allocation as follows:

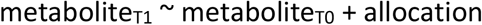

Where metabolite_T1_ is metabolite presence at 12 months (1 = present / 0 = absent) and metabolite_T0_ is metabolite presence at baseline (1 = present / 0 = absent) and allocation is as defined above. In this exploratory model, no covariates were fitted to maximise the power of the test. Model betas for ‘allocation’ represent the coefficient for intervention group, that is the log odds ratio between intervention group and control group. Results from this analysis were considered the primary result for all features with greater than or equal to 40%missing (unquantified) data, with a p-value of *p*<0.05 considered as evidence for association. In this case, no adjustment was made for multiple testing.

Metabolite data preparation, data pre-processing, statistical analyses and figure generation were performed in R Studio v.1.0.143 (58) using R v.4.0.2 (59).

## Data Availability

The terms of participant consent in DiRECT do not allow making the study data freely available in its raw form. The data used for analysis will be placed on a research data repository (https://researchdata.gla.ac.uk/) with access given to researchers subject to appropriate data sharing agreements. The R code for this study have been made publicly available on GitHub at: https://github.com/lauracorbin/metabolomics_of_direct.

## Data Availability

The terms of participant consent in DiRECT do not allow making the study data freely available in its raw form. The data used for analysis will be placed on a research data repository (https://researchdata.gla.ac.uk/) with access given to researchers subject to appropriate data sharing agreements.

## Code Availability

The R code for this study have been made publicly available on GitHub at: https://github.com/lauracorbin/metabolomics_of_direct.

## Acknowledgments

The DiRECT study was funded as a Strategic Research Initiative by Diabetes UK (award number 13/0004691). The formula diet was donated by Cambridge Weight Plan. Neither organisation had any input into the study design, data analysis or interpretation. We thank the National Health Service (NHS) Primary Care Research Network and North East Commissioning Support for their support and valuable input to recruitment. We thank Elaine Butler, Josephine Cooney, Sara-Jane Duffus, and Philip Stewart from the University of Glasgow for providing technical assistance; Helen Pilkington from the Newcastle upon Tyne Hospitals NHS Foundation Trust for providing research nurse support; and Sarah Weeden and Sarah Cadzow from the Robertson Centre for Biostatistics. We thank Wilma Leslie and research dietitians Louise McCombie, George Thom, Naomi Brosnahan and Alison Barnes for their contributions to the collection and transport of samples. We are enormously grateful to the GP practices, health-care professionals, and volunteers for their participation. Metabolomics analysis was funded by Wellcome Trust (award number 202802/Z/16/Z).

NJT is a Wellcome Trust Investigator (202802/Z/16/Z), is the PI of the Avon Longitudinal Study of Parents and Children (MRC & WT 217065/Z/19/Z), is supported by the University of Bristol NIHR Biomedical Research Centre (BRC-1215-2001), the MRC Integrative Epidemiology Unit (MC_UU_00011/1) and works within the CRUK Integrative Cancer Epidemiology Programme (C18281/A29019). D.A.H. & L.J.C. are supported by N.J.T.’s Wellcome Investigator Award (202802/Z/16/Z). E.E.V. is supported by a Diabetes UK RD Lawrence Fellowship (17/0005587), by Cancer Research UK (C18281/A29019) and by the World Cancer Research Fund (WCRF UK), as part of the World Cancer Research Fund International grant program (IIG_2019_2009). C.J.B is supported by E.E.V.’s World Cancer Research Fund grant (IIG_2019_2009). M.L.S is supported by the Wellcome Trust through a PhD studentship [218495/Z/19/Z]. RT is the CI of the study ‘Reversal of Type 2 diabetes Upon Normalisation of Energy intake in non-obese people’ (ReTUNE) funded by Diabetes UK (17/0005645).

This research was funded in whole, or in part, by the Wellcome Trust [202802/Z/16/Z]. For the purpose of Open Access, the author has applied a CC BY public copyright licence to any Author Accepted Manuscript version arising from this submission.

## Author Contributions

Conceptualization, R.T., N.S., and N.J.T.; Methodology, L.J.C., D.A.H., N.S. and N.J.T.; Formal Analysis, L.J.C., Resources, N.S., R.T., and M.E.J.L., Data Curation, A.M., and C-M.M., Writing – Original Draft, L.J.C., Writing – Review & Editing, L.J.C., D.A.H., C.J.B., E.E.V., M.L.S., P.W., R.T., N.S., and N.J.T.; Visualisation, L.J.C., and D.A.H.; Supervision, R.T., N.S., M.E.J.L., and N.J.T.; Funding Acquisition, R.T., N.S., M.E.J.L., and N.J.T.

## Declaration of Interests

RT has received lecture honoraria from Eli Lilly, Nestle Health and Janssen. All other authors declare no competing interests.

## Supplemental Information

Supplementary Document 1:

Supplementary Methods

Supplementary Table S1

Supplementary Figures S1, S3, S4, S5

Supplementary Document 2:

Quality control report for Metabolon (MS) data

Supplementary Document 3:

Quality control report for Nightingale (NMR) data

Supplementary Document 4:

Supplementary Figure S2A: Boxplots of raw (untransformed and unadjusted) MS metabolite data to show between group differences at baseline and at 12-months for 127 intervention-associated metabolites.

Supplementary Document 5:

Supplementary Figure S2B: Boxplots of raw (untransformed and unadjusted) NMR metabolite data to show between group differences at baseline and at 12-months for 59 intervention-associated features.

Supplementary Document 6:

Supplementary Figure S5: Distribution of metabolite change by weight change tertiles and remission status at 12 months. Metabolite change (the difference between the value at 12 months and that at baseline) was calculated using raw (untransformed and unadjusted) metabolite data. Data shown for the ten metabolites labelled on **Figure 4C** with all except isoleucine measured by MS.

Supplementary Document 7:

Excel file containing Supplementary Tables S2-S6

## Notes

### Clinical Trial

ISRCTN03267836

### Funding Statement

The DiRECT study was funded as a Strategic Research Initiative by Diabetes UK (award number 13/0004691). NJT is a Wellcome Trust Investigator (202802/Z/16/Z), is the PI of the Avon Longitudinal Study of Parents and Children (MRC & WT 217065/Z/19/Z) is supported by the University of Bristol NIHR Biomedical Research Centre (BRC-1215-2001), the MRC Integrative Epidemiology Unit (MC_UU_00011/1) and works within the CRUK Integrative Cancer Epidemiology Programme (C18281/A29019). D.A.H. & L.J.C. are supported by N.J.T.s Wellcome Investigator Award (202802/Z/16/Z). E.E.V. is supported by a Diabetes UK RD Lawrence Fellowship (17/0005587), by Cancer Research UK (C18281/A29019) and by the World Cancer Research Fund (WCRF UK), as part of the World Cancer Research Fund International grant program (IIG_2019_2009). C.J.B is supported by E.E.V.s World Cancer Research Fund grant (IIG_2019_2009). M.L.S is supported by the Wellcome Trust through a PhD studentship [218495/Z/19/Z]. RT is the CI of the study "Reversal of Type 2 diabetes Upon Normalisation of Energy intake in non-obese people" (ReTUNE) funded by Diabetes UK (17/0005645).

### Author Declarations

West of Scotland REC 3 of NHS Health Research Authority gave ethical approval for this work in January 2014, with approvals by the National Health Service (NHS) health board areas in Scotland and clinical commissioning groups in Tyneside.

## References

1. Jin Q, Ma RCW. Metabolomics in Diabetes and Diabetic Complications: Insights from Epidemiological Studies. Cells 2021;10.

2. Clish CB. Metabolomics: an emerging but powerful tool for precision medicine. Mol Case Stud 2015;1:a000588.

3. Arneth B, Arneth R, Shams M. Metabolomics of Type 1 and Type 2 Diabetes. Int J Mol Sci 2019;20:2467.

4. Urpi-Sarda M, Almanza-Aguilera E, Llorach R, et al. Non-targeted metabolomic biomarkers and metabotypes of type 2 diabetes: A cross-sectional study of PREDIMED trial participants. Diabetes Metab 2019;45:167–174.

5. Suhre K, Meisinger C, Döring A, et al. Metabolic Footprint of Diabetes: A Multiplatform Metabolomics Study in an Epidemiological Setting Breant B (ed.). PLoS One 2010;5:e13953.

6. Roberts LD, Koulman A, Griffin JL. Towards metabolic biomarkers of insulin resistance and type 2 diabetes: progress from the metabolome. Lancet Diabetes Endocrinol 2014;2:65–75.

7. Cirulli ET, Guo L, Leon Swisher C, et al. Profound Perturbation of the Metabolome in Obesity Is Associated with Health Risk. Cell Metab 2019;29:488–500 e2.

8. Abu Bakar MH, Sarmidi MR, Cheng K-K, et al. Metabolomics – the complementary field in systems biology: a review on obesity and type 2 diabetes. Mol Biosyst 2015;11:1742–1774.

9. Guasch-Ferré M, Hruby A, Toledo E, et al. Metabolomics in Prediabetes and Diabetes: A Systematic Review and Meta-analysis. 2016.

10. Bell JA, Bull CJ, Gunter MJ, et al. Early metabolic features of genetic liability to type 2 diabetes: Cohort study with repeated metabolomics across early life. Diabetes Care 2020;43:1537–1545.

11. Wang Q, Holmes M V., Smith GD, Ala-Korpela M. Genetic support for a causal role of insulin resistance on circulating branched-chain amino acids and inflammation. Diabetes Care 2017;40:1779–1786.

12. Lotta LA, Scott RA, Sharp SJ, et al. Genetic Predisposition to an Impaired Metabolism of the Branched-Chain Amino Acids and Risk of Type 2 Diabetes: A Mendelian Randomisation Analysis Minelli C (ed.). PLOS Med 2016;13:e1002179.

13. Rebholz CM, Yu B, Zheng Z, et al. Serum metabolomic profile of incident diabetes. Diabetologia 2018;61:1046–1054.

14. Yang SJ, Kwak S-Y, Jo G, Song T-J, Shin M-J. Serum metabolite profile associated with incident type 2 diabetes in Koreans: findings from the Korean Genome and Epidemiology Study. Sci Rep 2018;8:8207.

15. Liu J, Semiz S, van der Lee SJ, et al. Metabolomics based markers predict type 2 diabetes in a 14-year follow-up study. Metabolomics 2017;13:104.

16. Suvitaival T, Bondia-Pons I, Yetukuri L, et al. Lipidome as a predictive tool in progression to type 2 diabetes in Finnish men. Metabolism 2018;78:1–12.

17. Tulipani S, Griffin J, Palau-Rodriguez M, et al. Metabolomics-guided insights on bariatric surgery versus behavioral interventions for weight loss. Obesity 2016;24:2451–2466.

18. Taylor R, Leslie WS, Barnes AC, et al. Clinical and metabolic features of the randomised controlled Diabetes Remission Clinical Trial (DiRECT) cohort. Diabetologia 2018;61:589–598.

19. Lean ME, Leslie WS, Barnes AC, et al. Primary care-led weight management for remission of type 2 diabetes (DiRECT): an open-label, cluster-randomised trial. Lancet (London, England) 2018;391:541–551.

20. Stacklies W, Redestig H, Scholz M, Walther D, Selbig J. pcaMethods - A bioconductor package providing PCA methods for incomplete data. Bioinformatics 2007;23:1164–1167.

21. Ahola-Olli A V., Mustelin L, Kalimeri M, et al. Circulating metabolites and the risk of type 2 diabetes: a prospective study of 11,896 young adults from four Finnish cohorts. Diabetologia 2019;62:2298–2309.

22. Jump RLP, Polinkovsky A, Hurless K, et al. Metabolomics analysis identifies intestinal microbiota-derived biomarkers of colonization resistance in clindamycin-treated mice. PLoS One 2014;9.

23. Holeček M. Branched-chain amino acids in health and disease: Metabolism, alterations in blood plasma, and as supplements. Nutr Metab 2018;15:33.

24. Würtz P, Wang Q, Kangas AJ, et al. Metabolic Signatures of Adiposity in Young Adults: Mendelian Randomization Analysis and Effects of Weight Change. PLOS Med 2014;11:e1001765.

25. Shah SH, Crosslin DR, Haynes CS, et al. Branched-chain amino acid levels are associated with improvement in insulin resistance with weight loss. Diabetologia 2012;55:321–330.

26. Fontana L, Cummings NE, Arriola Apelo SI, et al. Decreased Consumption of Branched-Chain Amino Acids Improves Metabolic Health. Cell Rep 2016;16:520–530.

27. Yu D, Richardson NE, Green CL, et al. The adverse metabolic effects of branched-chain amino acids are mediated by isoleucine and valine. Cell Metab 2021;33:905-922.e6.

28. Alam YH, Kim R, Jang C. Metabolism and Health Impacts of Dietary Sugars. J Lipid Atheroscler 2022;11:20–38.

29. Mardinoglu A, Stančáková A, Lotta LA, et al. Plasma Mannose Levels Are Associated with Incident Type 2 Diabetes and Cardiovascular Disease. Cell Metab 2017;26:281–283.

30. Ferrannini E, Bokarewa M, Brembeck P, et al. Mannose is an insulin-regulated metabolite reflecting whole-body insulin sensitivity in man. Metabolism 2020;102.

31. Sharma V, Smolin J, Nayak J, et al. Mannose Alters Gut Microbiome, Prevents Diet-Induced Obesity, and Improves Host Metabolism. Cell Rep 2018;24:3087–3098.

32. Brites P, Waterham HR, Wanders RJA. Functions and biosynthesis of plasmalogens in health and disease. Biochim Biophys Acta - Mol Cell Biol Lipids 2004;1636:219–231.

33. Wallner S, Schmitz G. Plasmalogens the neglected regulatory and scavenging lipid species. Chem Phys Lipids 2011;164:573–589.

34. Mousa A, Naderpoor N, Mellett N, et al. Lipidomic profiling reveals early-stage metabolic dysfunction in overweight or obese humans. Biochim Biophys Acta - Mol Cell Biol Lipids 2019;1864:335–343.

35. Rosen RT, Hiserodt RD, Fukuda EK, et al. Determination of allicin, S-allylcysteine and volatile metabolites of garlic in breath, plasma or simulated gastric fluids. In: Journal of Nutrition.Vol 131. American Institute of Nutrition, 2001, pp 968S–971S.

36. Wang Y, Gapstur SM, Carter BD, et al. Untargeted metabolomics identifies novel potential biomarkers of habitual food intake in a cross-sectional study of postmenopausal women. J Nutr 2018;148:932–943.

37. Borucki K, Schreiner R, Dierkes J, et al. Detection of Recent Ethanol Intake With New Markers: Comparison of Fatty Acid Ethyl Esters in Serum and of Ethyl Glucuronide and the Ratio of 5-Hydroxytryptophol to 5-Hydroxyindole Acetic Acid in Urine. Alcohol Clin Exp Res 2005;29:781–787.

38. van de Luitgaarden IAT, Schrieks IC, Kieneker LM, et al. Urinary Ethyl Glucuronide as Measure of Alcohol Consumption and Risk of Cardiovascular Disease: A Population-Based Cohort Study. J Am Heart Assoc 2020;9:e014324.

39. Koh A, Molinaro A, Ståhlman M, et al. Microbially Produced Imidazole Propionate Impairs Insulin Signaling through mTORC1. Cell 2018;175:947-961.e17.

40. Molinaro A, Bel Lassen P, Henricsson M, et al. Imidazole propionate is increased in diabetes and associated with dietary patterns and altered microbial ecology. Nat Commun 2020;11:5881.

41. Koh A, Mannerås-Holm L, Yunn NO, et al. Microbial Imidazole Propionate Affects Responses to Metformin through p38γ-Dependent Inhibitory AMPK Phosphorylation. Cell Metab 2020;32:643-653.e4.

42. Wahl S, Vogt S, Stückler F, et al. Multi-omic signature of body weight change: results from a population-based cohort study. BMC Med 2015;13:48.

43. Perez-Cornago A, Brennan L, Ibero-Baraibar I, et al. Metabolomics identifies changes in fatty acid and amino acid profiles in serum of overweight older adults following a weight loss intervention. J Physiol Biochem 2014;70:593–602.

44. Taylor R, Al-Mrabeh A, Zhyzhneuskaya S, et al. Remission of Human Type 2 Diabetes Requires Decrease in Liver and Pancreas Fat Content but Is Dependent upon Capacity for β Cell Recovery. Cell Metab 2018;28:547-556.e3.

45. März W, Scharnagl H, Winkler K, et al. Low-density lipoprotein triglycerides associated with low-grade systemic inflammation, adhesion molecules, and angiographic coronary artery disease: The Ludwigshafen Risk and Cardiovascular Health Study. Circulation 2004;110:3068–3074.

46. EL L, KG H, BS A, MJ C, JC M, R T. Reversal of type 2 diabetes: normalisation of beta cell function in association with decreased pancreas and liver triacylglycerol. Diabetologia 2011;54:2506–2514.

47. Taylor R. Pathogenesis of type 2 diabetes: Tracing the reverse route from cure to cause. Diabetologia 2008;51:1781–1789.

48. Corbin KD, Zeisel SH. Choline metabolism provides novel insights into nonalcoholic fatty liver disease and its progression. Curr Opin Gastroenterol 2012;28:159–165.

49. Al-Sulaiti H, Diboun I, Agha M V., et al. Metabolic signature of obesity-associated insulin resistance and type 2 diabetes. J Transl Med 2019;17.

50. Leslie WS, Ford I, Sattar N, et al. The Diabetes Remission Clinical Trial (DiRECT): protocol for a cluster randomised trial. BMC Fam Pract 2016;17:20.

51. Soininen P, Kangas AJ, Würtz P, Suna T, Ala-Korpela M. Quantitative Serum Nuclear Magnetic Resonance Metabolomics in Cardiovascular Epidemiology and Genetics. Circ Cardiovasc Genet 2015;8:192–206.

52. Soininen P, Kangas AJ, Würtz P, et al. High-throughput serum NMR metabonomics for cost-effective holistic studies on systemic metabolism. Analyst 2009;134:1781.

53. DeHaven CD, Evans AM, Dai H, Lawton KA. Organization of GC/MS and LC/MS metabolomics data into chemical libraries. J Cheminform 2010;2:9.

54. Evans AM, DeHaven CD, Barrett T, Mitchell M, Milgram E. Integrated, Nontargeted Ultrahigh Performance Liquid Chromatography/Electrospray Ionization Tandem Mass Spectrometry Platform for the Identification and Relative Quantification of the Small-Molecule Complement of Biological Systems. Anal Chem 2009;81:6656–6667.

55. Hughes DA, Taylor K, McBride N, et al. metaboprep: an R package for preanalysis data description and processing. Bioinformatics 2022.

56. Holm S. A simple sequentially rejective multiple test procedure. Scand J Stat 1979;6:65–70.

57. Nagi D, Hambling C, Taylor R. Remission of type 2 diabetes: a position statement from the Association of British Clinical Diabetologists (ABCD) and the Primary Care Diabetes Society (PCDS). Br J Diabetes 2019;19:73–76.

58. RStudio Team. RStudio: Integrated Development Environment. 2016.

59. R Core Team. R: A Language and Environment for Statistical Computing. 2020.

