## Supplementary Document 1 for "The metabolomic signature of weight loss in the Diabetes Remission Clinical Trial (DiRECT)"

Corbin *et al.*

### Supplementary Methods

#### Extended metabolite data acquisition

##### Nuclear magnetic resonance spectroscopy

A serum $^{1}$H-NMR metabolomics platform (Nightingale Health Ltd, Helsinki, Finland) was used to quantify circulating metabolites with an emphasis on lipid or lipoprotein lipid measures (1). Data were received from Nightingale on 19$^{th}$ March 2019.

##### Mass spectroscopy data

The methodological details provided herein are as supplied by Metabolon, Inc. Data were issued by Metabolon on 16$^{th}$ February 2018.

*Sample preparation*

Samples were prepared using the automated MicroLab STAR® system from Hamilton Company. Several recovery standards were added prior to the first step in the extraction process for QC purposes. Proteins were precipitated with methanol under vigorous shaking for 2 min (Glen Mills GenoGrinder 2000) followed by centrifugation. The resulting extract was divided into five fractions: two for analysis by two separate reverse phase (RP)/UPLC-MS/MS methods with positive ion mode electrospray ionization (ESI), one for analysis by RP/UPLC-MS/MS with negative ion mode ESI, one for analysis by HILIC/UPLC-MS/MS with negative ion mode ESI, and one for backup. Samples were placed briefly on a TurboVap® (Zymark) to remove the organic solvent. The sample extracts were stored overnight under nitrogen before preparation for analysis.

*Quality assurance / Quality control*

Three types of controls were used when analyzing the experimental samples: a pooled matrix sample generated by taking a small volume of each experimental sample (or alternatively, use of a pool of well-characterized human plasma); extracted water samples; and a cocktail of QC standards. Instrument variability was determined by calculating the median relative standard deviation (RSD) for the standards that were added to each sample prior to injection into the mass spectrometers. Overall process variability was determined by calculating the median RSD for all endogenous metabolites (i.e., non-instrument standards) present in 100% of the pooled matrix samples. Experimental samples were randomized across the platform run with QC samples spaced evenly among the injections.

*Ultrahigh Performance Liquid Chromatography-Tandem Mass Spectroscopy (UPLC-MS/MS)*

All methods utilized a Waters ACQUITY ultra-performance liquid chromatography (UPLC) and a Thermo Scientific Q-Exactive high resolution/accurate mass spectrometer interfaced with a heated electrospray ionization (HESI-II) source and Orbitrap mass analyzer operated at 35,000 mass resolution. The sample extract was dried then reconstituted in solvents compatible to each of the four methods. Each reconstitution solvent contained a series of standards at fixed concentrations to ensure injection and chromatographic consistency. One aliquot was analyzed using acidic positive ion conditions, chromatographically optimized for more hydrophilic compounds. In this method, the extract was gradient eluted from a C18 column (Waters UPLC BEH C18-2.1x100 mm, 1.7 µm) using water and methanol, containing 0.05% perfluoropentanoic acid (PFPA) and 0.1% formic acid (FA). Another aliquot was also analyzed using acidic positive ion conditions, however it was chromatographically optimized for more hydrophobic compounds. In this method, the extract was gradient eluted from the same afore mentioned C18 column using methanol, acetonitrile, water, 0.05% PFPA and 0.01% FA and was operated at an overall higher organic content. Another aliquot was analyzed using basic negative ion optimized conditions using a separate dedicated C18 column. The basic extracts were gradient eluted from the column using methanol and water, however with 6.5mM Ammonium Bicarbonate at pH 8. The fourth aliquot was analyzed via negative ionization following elution from a HILIC column (Waters UPLC BEH Amide 2.1x150 mm, 1.7 µm) using a gradient consisting of water and acetonitrile with 10mM Ammonium Formate, pH 10.8. The MS analysis alternated between MS and data-dependent MSn scans using dynamic exclusion. The scan range varied slighted between methods but covered 70-1000 m/z.

###### *Data extraction and compound identification*

Raw data were extracted, peak-identified and QC processed using Metabolon's hardware and software. Compounds were identified by comparison to library entries of purified standards or recurrent unknown entities. More than 3300 commercially available purified standard compounds have been acquired and registered into LIMS for analysis on all platforms for determination of their analytical characteristics. Additional mass spectral entries have been created for structurally unnamed biochemicals, which have been identified by virtue of their recurrent nature (both chromatographic and mass spectral).
Metabolite Quantification and Data Normalization
Peaks were quantified using area-under-the-curve. A data normalization step was performed to correct variation resulting from instrument inter-day tuning differences.

###### *Metabolite library updates*

In February 2018, data for 959 known and 317 unnamed biochemicals were returned by Metabolon. In January 2022, Metabolon issued revised identifications for the following metabolites:

| **Incorrect identification** | **Correct identification** |
|---|---|
| 1-carboxyethylleucine | N-lactoyl leucine |
| 1-carboxyethylisoleucine | N-lactoyl isoleucine |
| 1-carboxyethylphenylalanine | N-lactoyl phenylalanine |
| 1-carboxyethyltyrosine | N-lactoyl tyrosine |
| 1-carboxyethylvaline | N-lactoyl valine |
| 1-carboxyethylhistidine | N-lactoyl histidine |

#### Extended metabolite data preparation

##### NMR data

Missingness was assessed by samples and by metabolite feature. By sample missingness rates were calculated based on the 148 primary measures (i.e. excluding derived measures[1]). Three samples with more than 20% of the primary measures unquantified were excluded. By feature missingness was calculated for all 227 measures after sample exclusions had been applied and any features with >20% missing values flagged for exclusion. The only primary feature to be excluded was 22:6, docosahexaenoic acid (DHA), concentrations of which were returned for only 32.2% of samples. This was likely due to interference with the quantification of this metabolite in the majority of the samples, probably originating from the sample tubes used for sample collection. Therefore, measures of DHA and the ratio of DHA to total fatty acids were excluded from all analyses. After these sample and feature exclusions, the median (minimum, maximum) rate of sample missingness in primary measures (i.e. excluding derived measures) was 0.00 (0.0, 1.4) % and the median rate of metabolite feature missingness (across all 225 measures) was 0.0 (0.0, 14.5) %.

Following sample and feature filtering based on missingness, a principal component analysis (PCA) was performed (using the 147 primary measures only) in order to identify potential sample outliers. First, a hierarchical clustering approach was used to reduce the redundancy in the data (after exclusion of the derived measures). A Spearman's correlation matrix was generated using the 'pairwise complete observations' of the base R cor() function applied to metabolite data for the 147 primary measures. From this a distance matrix was constructed (calculated as 1 minus the absolute correlations) and used to build a dendrogram using hclust() and the method 'complete' from the R 'stats' package. The resulting tree was cut at a height of 0.2 (corresponding to a maximum within cluster correlation of 0.8) giving 37 groups of correlated metabolites. A single representative metabolite was selected from each group for inclusion in the PCA based on minimum missingness.  This reduced dataset was then centred and scaled, and a probabilistic PCA (2) implemented; the probabilistic method was used to allow for missing values. Samples located more than five standard deviations (5 SD) from the mean of the first and/or second principal components were excluded. The PCA was then re-run to check for further outliers.  This procedure resulted in the exclusion of one sample. Finally, three samples were removed on the basis of quality control tags supplied as part of the data release supplied by Nightingale and indicative of poor sample quality, namely, high pyruvate, high lactate and low glutamine/high lactate ratio (as compared to the expected values based on benchmarking). After filtering, there were 567 samples and 225 metabolites (147 primary measures and 78 derived ratios) available for the next steps in the analysis.

##### MS data

Missingness was assessed by samples and by metabolite feature.  By sample missingness rates were calculated after excluding 203 features designated as xenobiotics[2] (leaving 1073

---

[1] In data from Nightingale Health, derived variables are metabolite traits that are a summary of two or more other metabolites (possibly already represented in the dataset). These variables can introduce bias in estimates of sample missingness (where a single metabolite is missing, any derived measures based on that metabolite will also be missing) and may not be appropriate to retain when identifying a set of representative metabolites for the data set.

[2] Xenobiotics are metabolites not produced by the body, such as drug compounds and consequently can have very high rates of missingness, while still being critically informative to a study. For this reason, we do not include xenobiotics when calculating by sample missingness but would advocate affording them special consideration in any downstream statistical analyses.

metabolites). There were no samples with more than 20% of metabolites unquantified and therefore no samples were excluded on the basis of high missingness. By feature missingness was then calculated for all 1,276 metabolites and 22 features with less than five observations across the entire set of samples excluded. After these sample and feature exclusions, the median (minimum, maximum) rate of sample missingness in primary measures was 15.3 (10.3, 25.0) % and the median rate of metabolite feature missingness (across all 225 measures) was 1.0 (0.0, 99.1) %. Note, the maximum by sample missingness in the filtered set is greater than the 20% threshold applied because data for xenobiotics was included in this summary.

Following sample and feature filtering based on missingness, total peak area (TPA) per sample was calculated as the sum of peak areas across all features. Samples with a TPA more than 5 SD from the mean (above or below) were identified for exclusion; no samples were excluded based on these criteria. Next, the hierarchical clustering and probabilistic PCA was performed as described above (in the NMR data section) but using only those features with <20% missing data as input to the hierarchical clustering step. No sample outliers were identified as a result of the PCA. Since the same sample material was used for both the NMR and MS-based analyses, any samples excluded from the NMR data on the basis of NMR quality control tags were also excluded from the MS dataset. After filtering, there were 571 samples and 1,254 metabolites for the next steps in the analysis.

The metabolite data preparation procedures described were performed in R Studio v.1.0.143 (3) using R v.4.0.2 (4).

#### Extended statistical analysis

##### Descriptive analysis

To enable robust statistical analyses that would not be unduly affected by distributional characteristics typical of metabolomics data such as non-normal distributions and small numbers of extreme values (typically at the high end of the distribution), the primary linear regression model-based analysis was performed on the RNT dataset. However, this can make the comparison of relative effect sizes across metabolites challenging. Therefore, we also calculated the median fold change difference on the raw data as the ratio between the two group medians (intervention group median/control group median); no adjustments were made for other variables. This fold change was transformed to a log2 scale before plotting.

For all metabolites shown to be associated with the intervention in the primary linear regression model-based analysis, we checked for a difference in metabolite levels at baseline. First, raw metabolite data were centred (by subtracting the mean) and scaled (by dividing by the standard deviation) within time point. Then, a two-sample Wilcoxon rank sum test was performed to compare metabolite levels in control and intervention groups at baseline and at 12-months. No adjustments were made for other variables.

##### Exploratory analysis of associated metabolites and clinical phenotypes

In this analysis, data from the two platforms (NMR and MS) were combined and restricted to features with <40% missing data at 12-months (i.e., those metabolites for which the linear model represented the primary analysis). A hierarchical clustering approach (by the same method as described above in '**Extended data preparation**') was applied to residuals extracted from a re-run of the linear model as described in the main manuscript (using the RNT metabolite data) but without allocation fitted. These residuals represent the metabolite levels at 12 months after adjusting for covariates. The resulting dendrogram was cut at a height of 0.80 (which corresponds to a maximum within cluster correlation of 0.2) to define a set of metabolite clusters. The list of associated features from the linear model analysis was then reduced such that a single feature per cluster was retained for the next step; the feature to be retained was selected on the basis of a principal variables analysis (PVA) such that the feature that captured the most variation within the cluster was kept as the representative metabolite (using 'PVA' function from the 'growthPheno' R package (5)). Model residuals for the subset of representative metabolites were then used as input to a sample based probabilistic PCA. The relationship between the derived (top) PCs and the aforementioned clinical phenotypes was assessed by Pearson's correlation (r) and presented as a biplot.

In addition, hypergeometric-based enrichment analyses were conducted in order to designate a super pathway to the clusters. To do so, each cluster containing more than five metabolites was tested for enrichment for particular classes as compared to all features tested in the linear model. The cluster was then designated a super pathway annotation according to the most enriched class.

#### Figure generation

*Heatmap (Figure S4)*: Heatmaps were generated using the R package 'heatmap3'(6, 7). The same set of residuals used for the PCA analysis described in the '**Extended statistical analysis'** section above were used as input. Dendrograms were built for samples and metabolites using the hclust() function from the R 'stats' package with the method 'ward.D2'(8, 9) specified.

*Biplots (Figure 4):* Plots presented in Figure 4 are based on the probabilistic PCA as described in the '**Exploratory analysis of associated metabolites and clinical phenotypes**' section above and in the main manuscript. Data points (samples) in Figures 4A and 4B are positioned based on scores for PC1 and PC2. Whilst the original loadings (for metabolites) are shown in Figure 4C, the biplot arrows in Figures 4A and 4B were redefined such that the length and direction of the arrows are scaled according to the correlation coefficients between the two PCs and the selected clinical phenotypes.

### Supplementary Tables

#### Table S1 Overview of metabolite data post-curation filtering

| | | NMR data | MS data |
|---|---|---|---|
| | **ORIGINAL DATASET** | **574 samples<br>227 metabolite features** | **547 samples<br>1276 metabolites** |
| **QC CRITERIA** | Sample missingness | Samples excluded if >20% features missing. Data from derived measures excluded.<br>**N=3 samples excluded** | Samples excluded if >20% features missing. Data from xenobiotics excluded.<br>**N=0 samples excluded** |
| | Feature missingness | Features excluded if >20% samples do not have a value.<br>**M=2 features excluded** | Features excluded if <5 observations.<br>**M=22 features excluded** |
| | Principal components analysis (PCA) to look for sample outliers | Samples excluded if >5SD from the mean on PC1 or PC2.<br>**N=1 sample excluded** | Samples excluded if >5SD from the mean on PC1 or PC2.<br>**N=0 sample excluded** |
| | QC fails from Nightingale indicative of poor sample quality | **N=3 samples excluded** | **N=3 samples excluded** |
| | **POST-FILTERING DATASET** | **567 samples<br>225 metabolite features** | **571 samples<br>1254 metabolites** |

#### Table S2 NMR data: Results of linear model
See Excel file.

#### Table S3 MS data: Results of linear model
See Excel file.

#### Table S4 Super pathway and cluster allocation
See Excel file.

#### Table S5 Relationship between the derived (top) PCs and the select clinical phenotypes as assessed by Pearson's correlation (r)
See Excel file.

#### Table S6 MS data: Results of logistic model
See Excel file.

#### Supplementary Figures

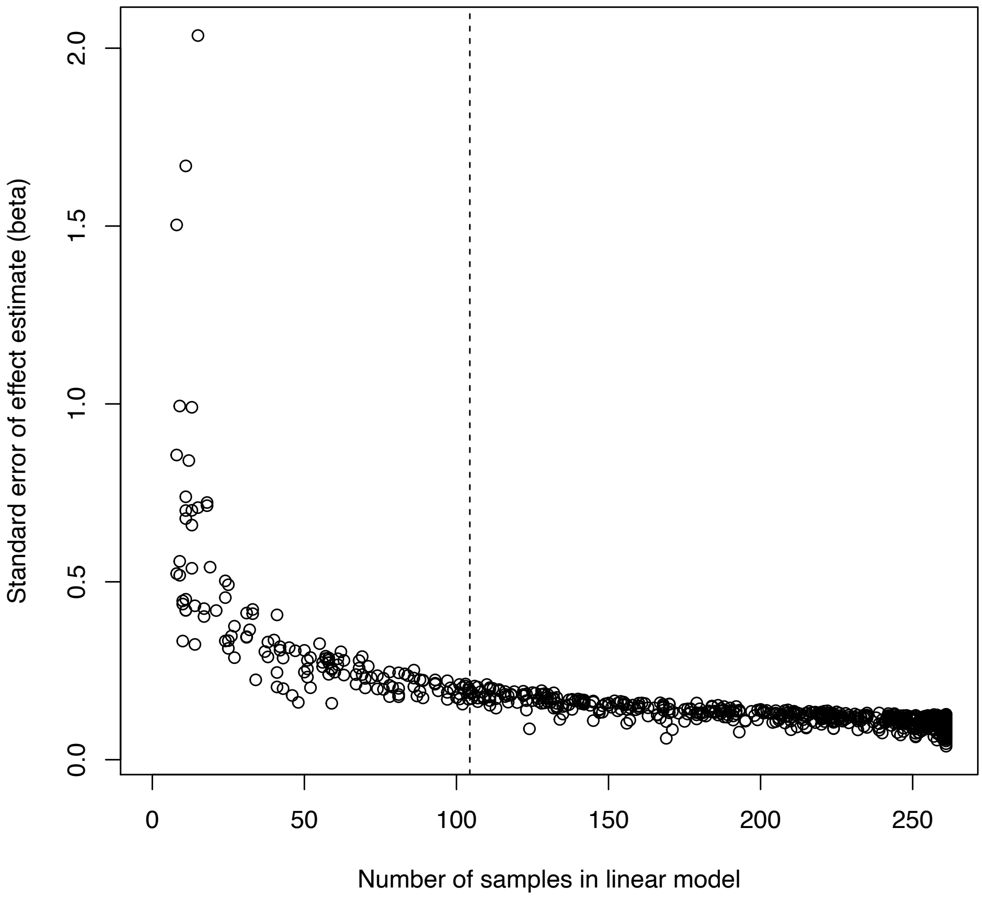

Figure S1. A demonstration of the poor performance of the linear model below the chosen threshold of missingness (<40%)

Figure S2A. Boxplots of raw (untransformed and unadjusted) MS metabolite data to show between group differences at baseline and at 12-months.

See separate supplementary PDF document. P-values from a Wilcoxon rank sum test.

Figure S2B. Boxplots of raw (untransformed and unadjusted) NMR metabolite data to show between group differences at baseline and at 12-months.

See separate supplementary PDF document. P-values from a Wilcoxon rank sum test.

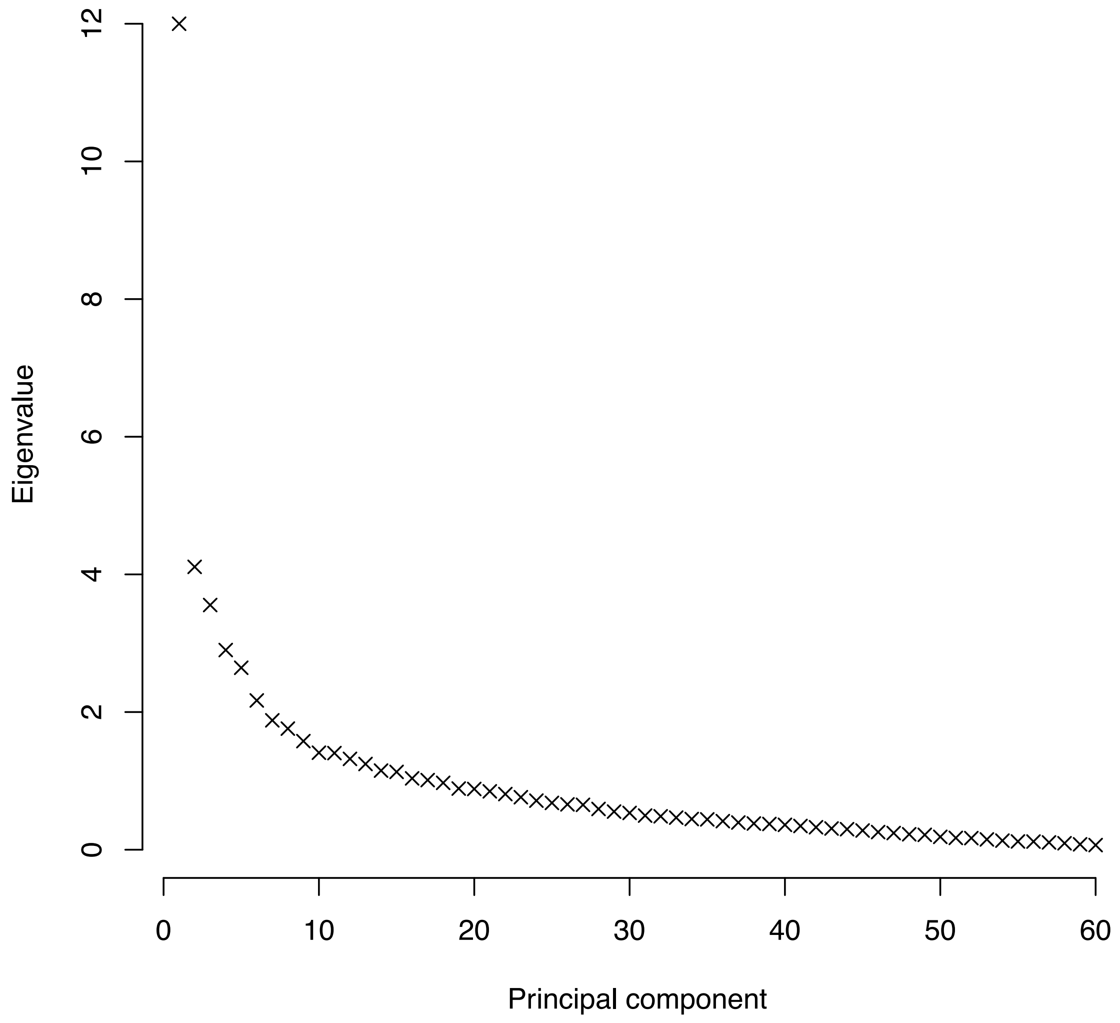

Figure S3. Scree from PCA performed on data for the 61 associated representative metabolites

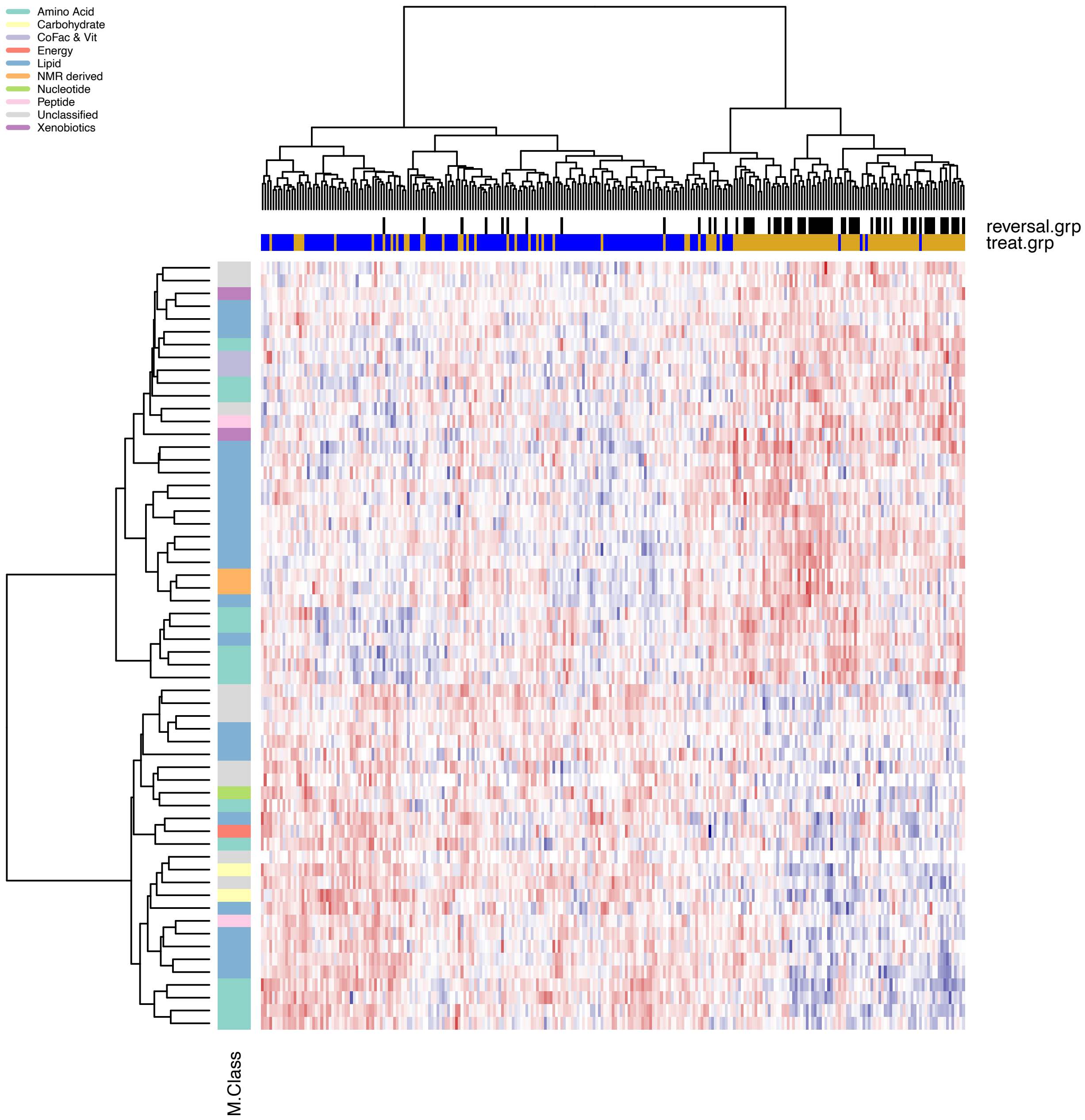

#### Figure S4 Heatmap based on PCA performed on data for the 61 associated representative metabolites

Further description of plot generation in **Supplementary Methods.**
*reversal.grp* = remission status at 12months: white = no remission; black = remission
*treat.grp* = group allocation: blue = control; orange = intervention.

#### Figure S5 Distribution of metabolite change by weight change tertiles and remission status at 12 months.

Metabolite change (the difference between the value at 12 months and that at baseline) was calculated using raw (untransformed and unadjusted) metabolite data. Data shown for the ten metabolites labelled on **Figure 4C** with all except isoleucine measured by MS.

See separate supplementary PDF document.

#### References

1. Soininen P, Kangas AJ, Würtz P, Suna T, Ala-Korpela M. Quantitative Serum Nuclear Magnetic Resonance Metabolomics in Cardiovascular Epidemiology and Genetics. *Circ Cardiovasc Genet* 2015;8:192–206.
2. Stacklies W, Redestig H, Scholz M, Walther D, Selbig J. pcaMethods - A bioconductor package providing PCA methods for incomplete data. *Bioinformatics* 2007;23:1164–1167.
3. RStudio Team. RStudio: Integrated Development Environment. 2016.
4. R Core Team. R: A Language and Environment for Statistical Computing. 2020.
5. Brien C. growthPheno: Plotting, Smoothing and Growth Trait Extraction for Longitudinal Data. 2019.
6. Zhao S, Guo Y, Sheng Q, Shyr Y. Heatmap3: an improved heatmap package with more powerful and convenient features. *BMC Bioinformatics* 2014;15:P16.
7. Zhao S, Yin L, Guo Y, Sheng Q, Shyr Y. heatmap3: An Improved Heatmap Package. 2020.
8. Ward JH. Hierarchical Grouping to Optimize an Objective Function. *J Am Stat Assoc* 1963;58:236–244.
9. Murtagh F, Legendre P. Ward's Hierarchical Agglomerative Clustering Method: Which Algorithms Implement Ward's Criterion? *J Classif* 2014;31:274–295.
