## Supplementary Document 2 for "The metabolomic signature of weight loss in the Diabetes Remission Clinical Trial (DiRECT)"

### Metabolon Metabolite Data QC Report

L.J.Corbin

31/03/2022

### Data overview

Number of samples in OrigScale file: 574
Number of features in OrigScale file: 1276

### Power exploration for case/control analysis

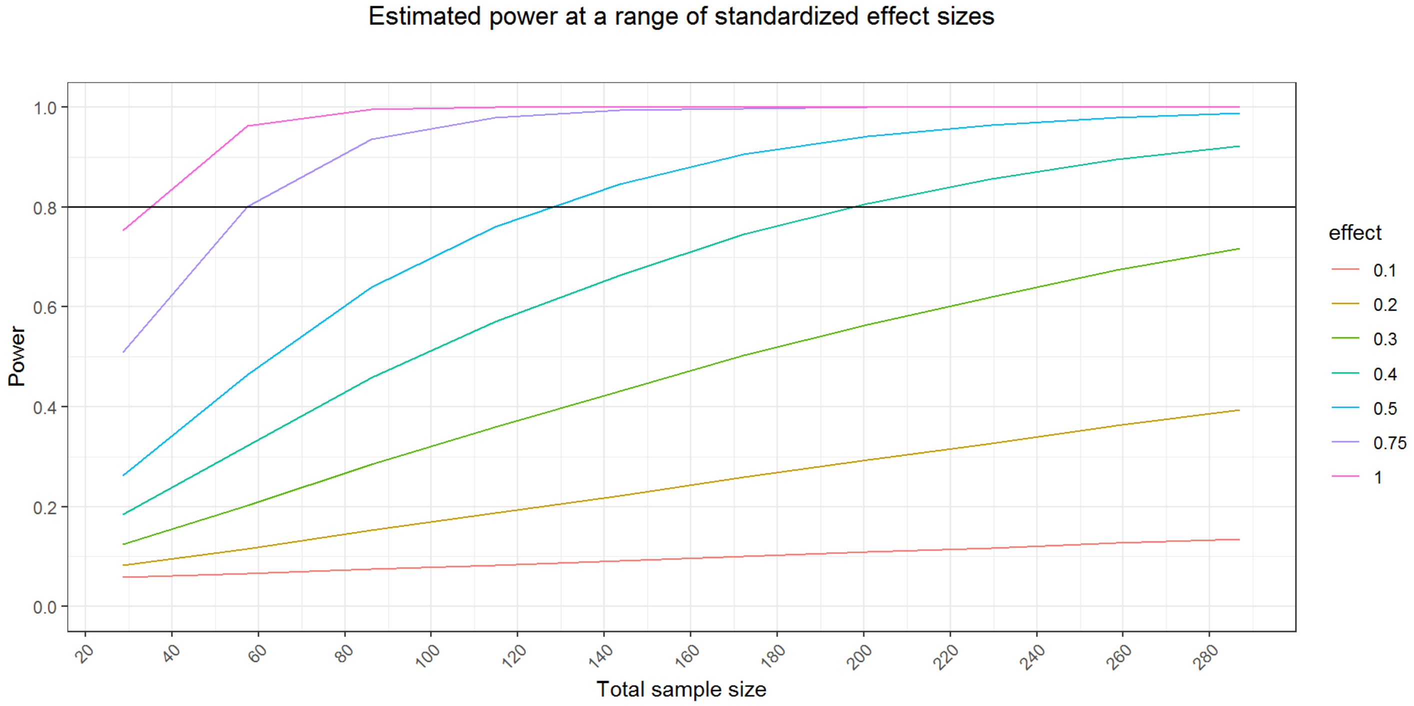


The minimum sample size required for achieving 80% power assuming a standardised effect size of 1, and an equal split between intervention/control groups, is: 34

### Power exploration for continuous outcome analysis

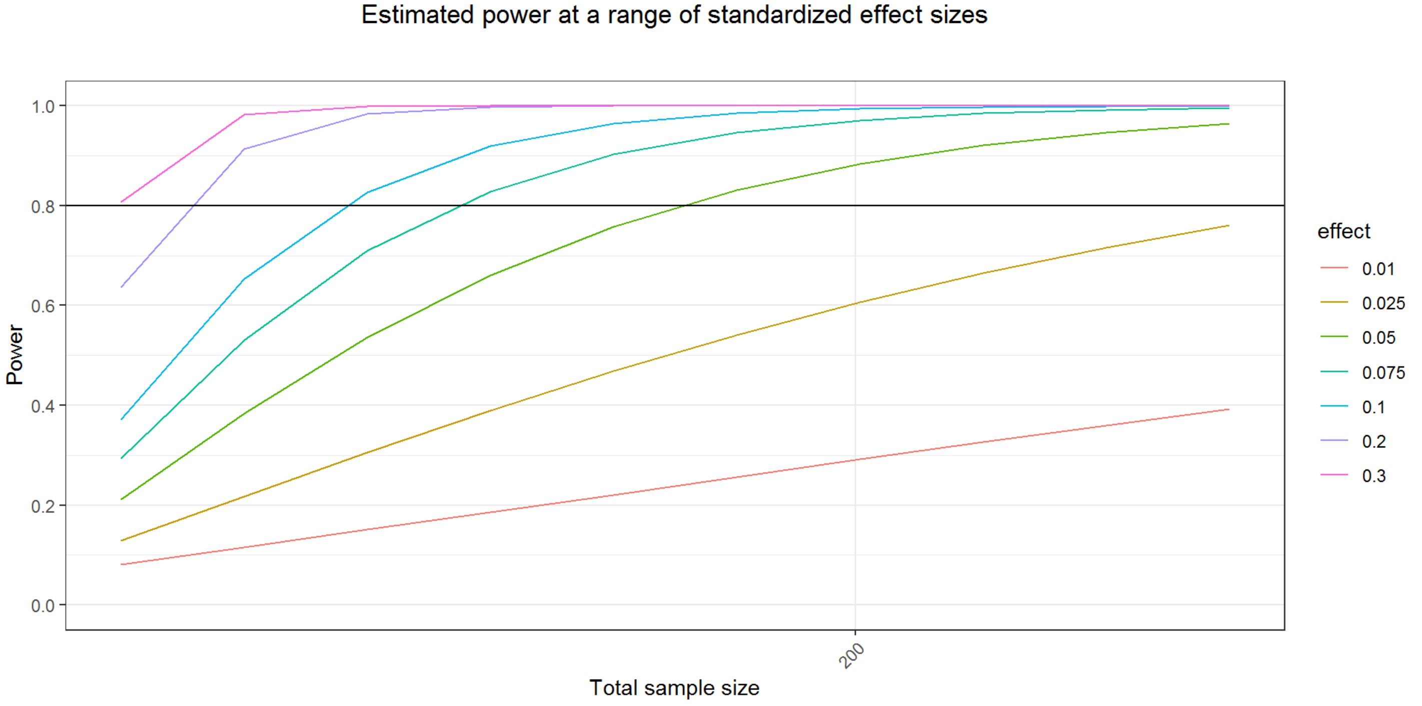


### Summarise missingness (pre filtering)

```
[1] "Summary of sample missingness:"
```

```
   Min. 1st Qu.  Median    Mean 3rd Qu.    Max. 
 0.1183  0.1530  0.1677  0.1706  0.1850  0.2625 
```

```
[1] "Summary of feature missingness:"
```

```
   Min. 1st Qu.  Median    Mean 3rd Qu.    Max. 
 0.0000  0.0000  0.0122  0.1706  0.2404  0.9983 
```

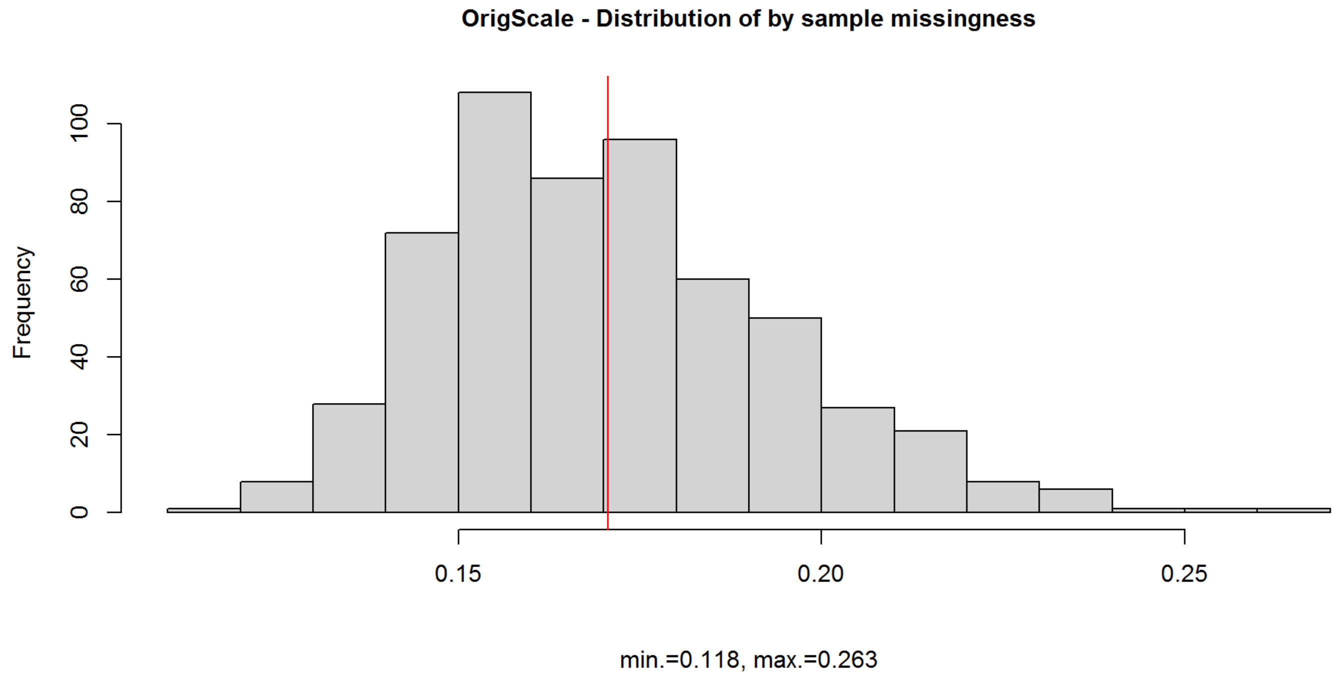


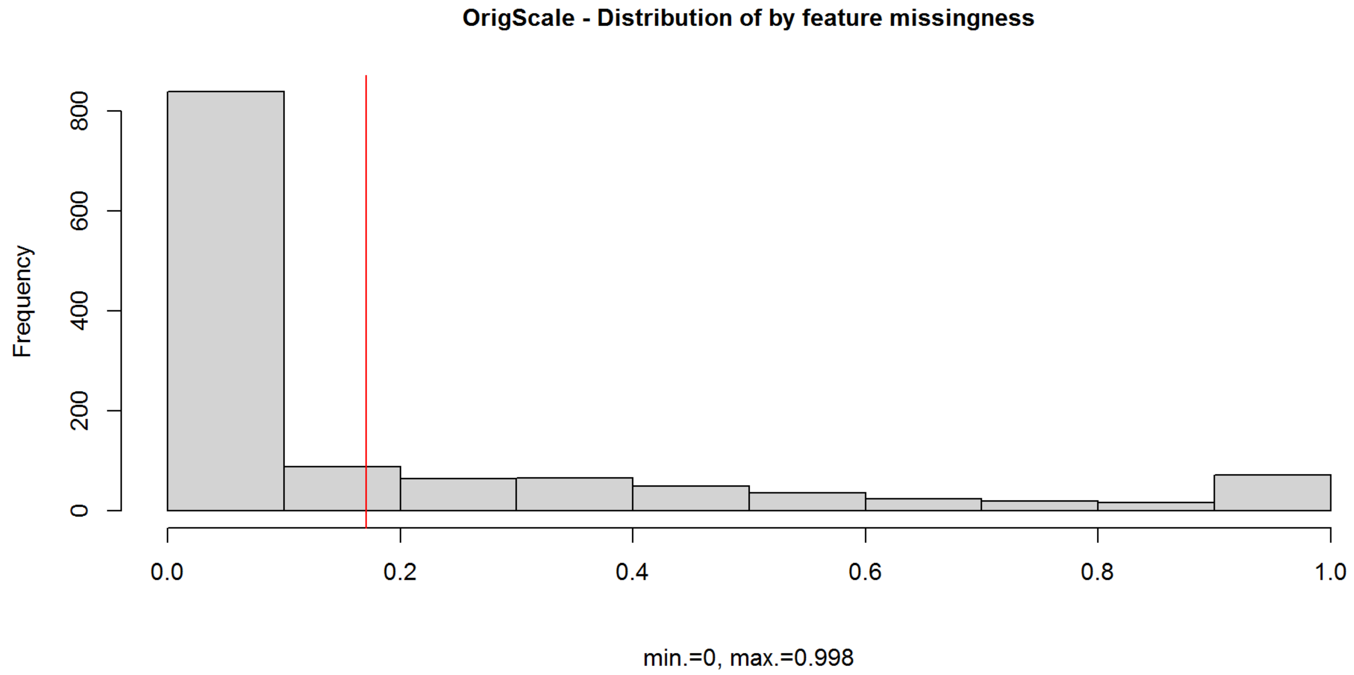


There are 440 features with no data missing.

#### Example sample numbers at different missingness rates

If feature has 10% missingness, data is available for: 516.6 samples.
If feature has 20% missingness, data is available for: 459.2 samples.
If feature has 30% missingness, data is available for: 401.8 samples.
If feature has 40% missingness, data is available for: 344.4 samples.
If feature has 50% missingness, data is available for: 287 samples.

### Summarise outliers (pre filtering)

#### Outliers are defined both as values greater than or less than 5SD from the mean and as values outside the 1st/99th percentile.

```
[1] "Summary of SD outliers by sample (pre filtering):"
```

```
   Min. 1st Qu.  Median    Mean 3rd Qu.    Max. 
  0.000   0.000   2.000   3.352   4.000  35.000 
```

```
[1] "Summary of SD outliers by feature (pre filtering):"
```

```
   Min. 1st Qu.  Median    Mean 3rd Qu.    Max. 
  0.000   0.000   1.000   1.508   2.000   8.000 
```

```
[1] "Summary of percentile outliers by sample (pre filtering):"
```

```
   Min. 1st Qu.  Median    Mean 3rd Qu.    Max. 
   2.00   12.00   18.00   22.94   28.00  102.00 
```

```
[1] "Summary of percentile outliers by feature (pre filtering):"
```

```
   Min. 1st Qu.  Median    Mean 3rd Qu.    Max. 
   0.00   10.00   12.00   10.32   12.00   12.00 
```

### Apply missingness thresholds to data and re-summarise

Sample missingness applied using all features except those classified as xenobiotics.
Samples excluded if more than 20% of features are missing.
Features excluded if they are measured in less than 5 samples.

Number of sample exclusions based on >20% missingness: 0
Sample missingness is calculated after exclusion of 203 xenobiotics, leaving 1073 features. Number of feature exclusions based on >0.9912892 missingness: 22

```
[1] "Summary of sample missingness after cleaning:"
```

```
   Min. 1st Qu.  Median    Mean 3rd Qu.    Max. 
 0.1029  0.1382  0.1531  0.1561  0.1707  0.2496 
```

```
[1] "Summary of feature missingness after cleaning:"
```

```
   Min. 1st Qu.  Median    Mean 3rd Qu.    Max. 
0.00000 0.00000 0.01045 0.15612 0.21646 0.99129 
```

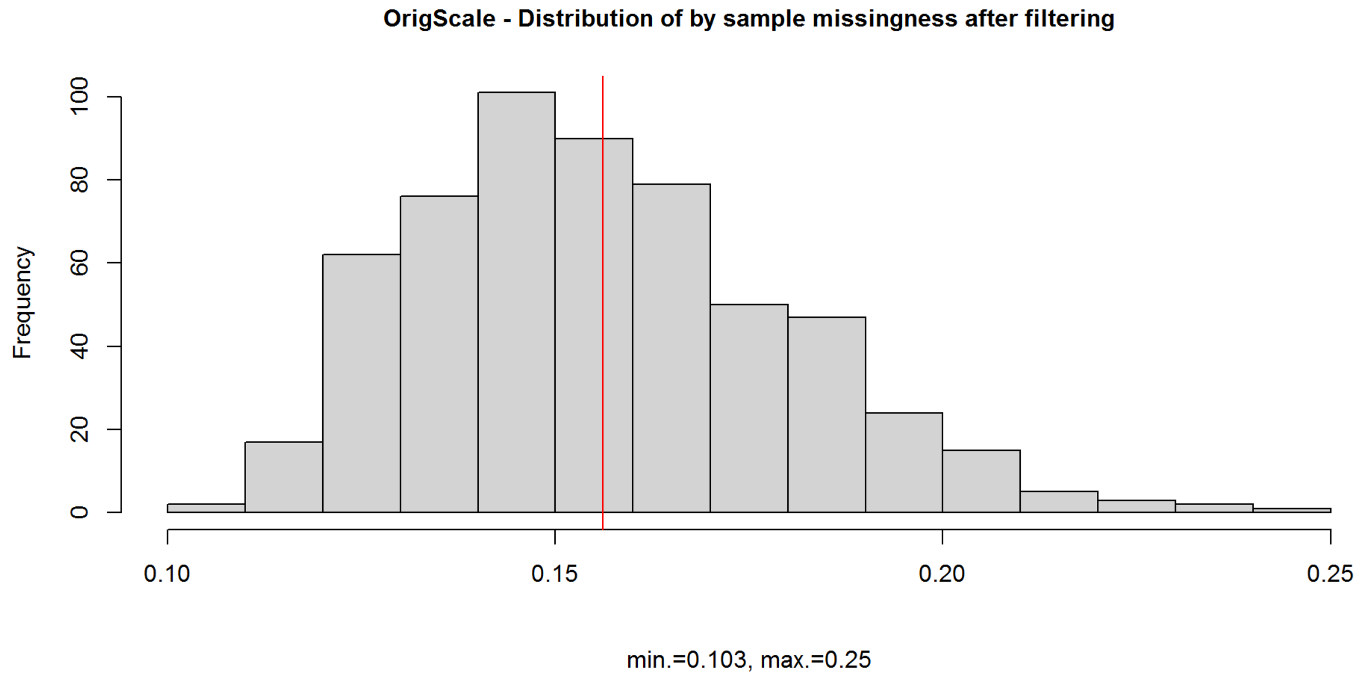


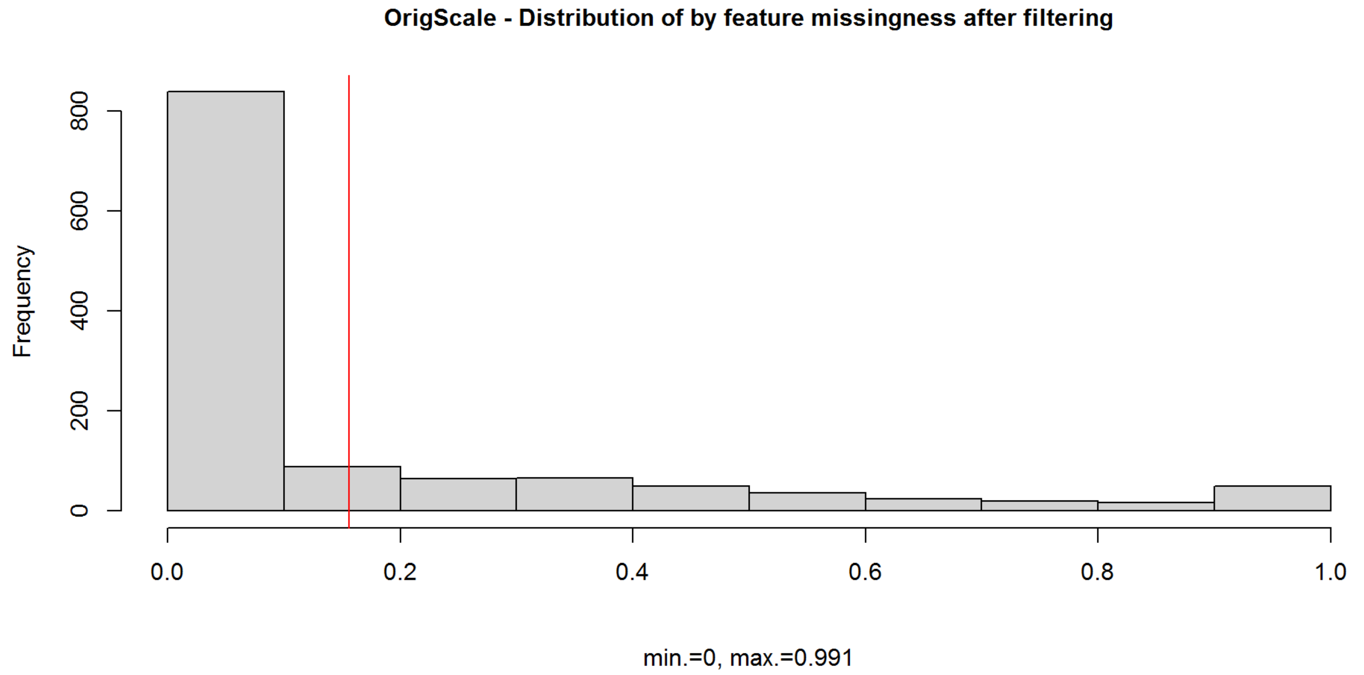


### Total peak area assessment (by sample) (based on filtered OrigScale data)

```
[1] "Summary of total peak area:"
```

```
     Min.   1st Qu.    Median      Mean   3rd Qu.      Max. 
3.142e+10 4.496e+10 4.923e+10 4.937e+10 5.348e+10 8.158e+10 
```

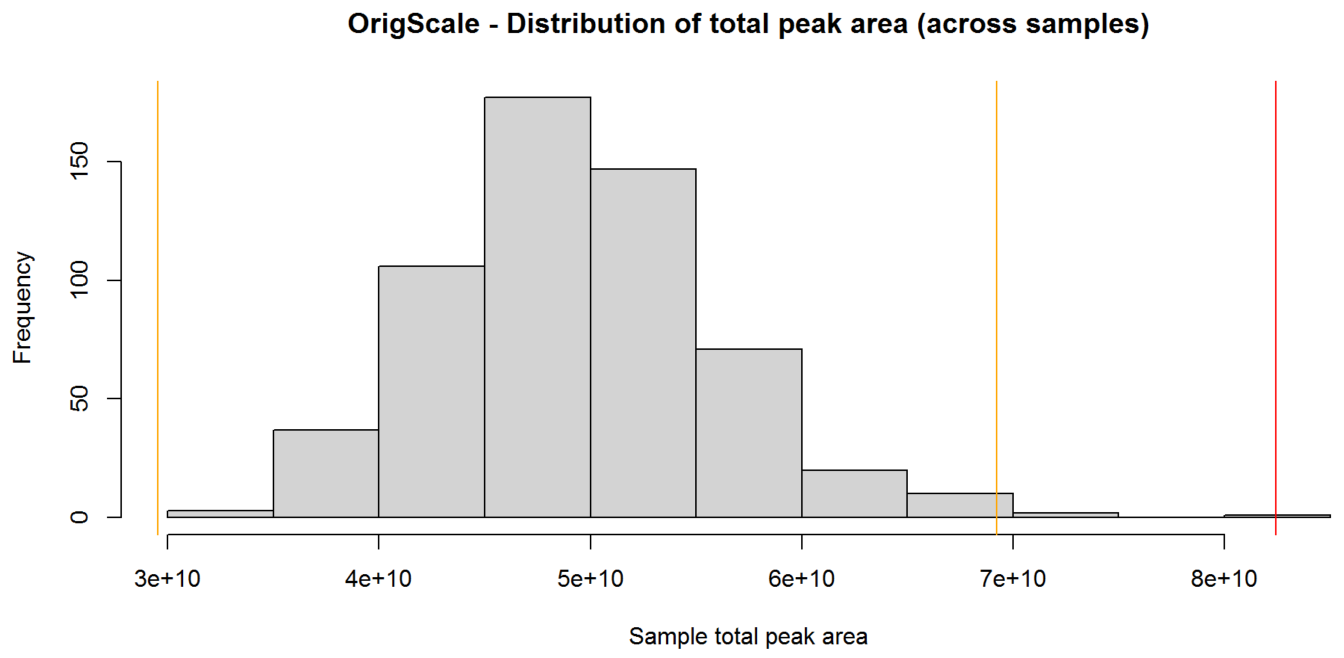


Number of sample exclusions based on total peak area (+/- 3sd from the mean): 5
Number of sample exclusions based on total peak area (+/- 5sd from the mean) (current criteria): 0

### Plot principal components - check for sample outliers

#### Identify independent features based on Spearman's correlation

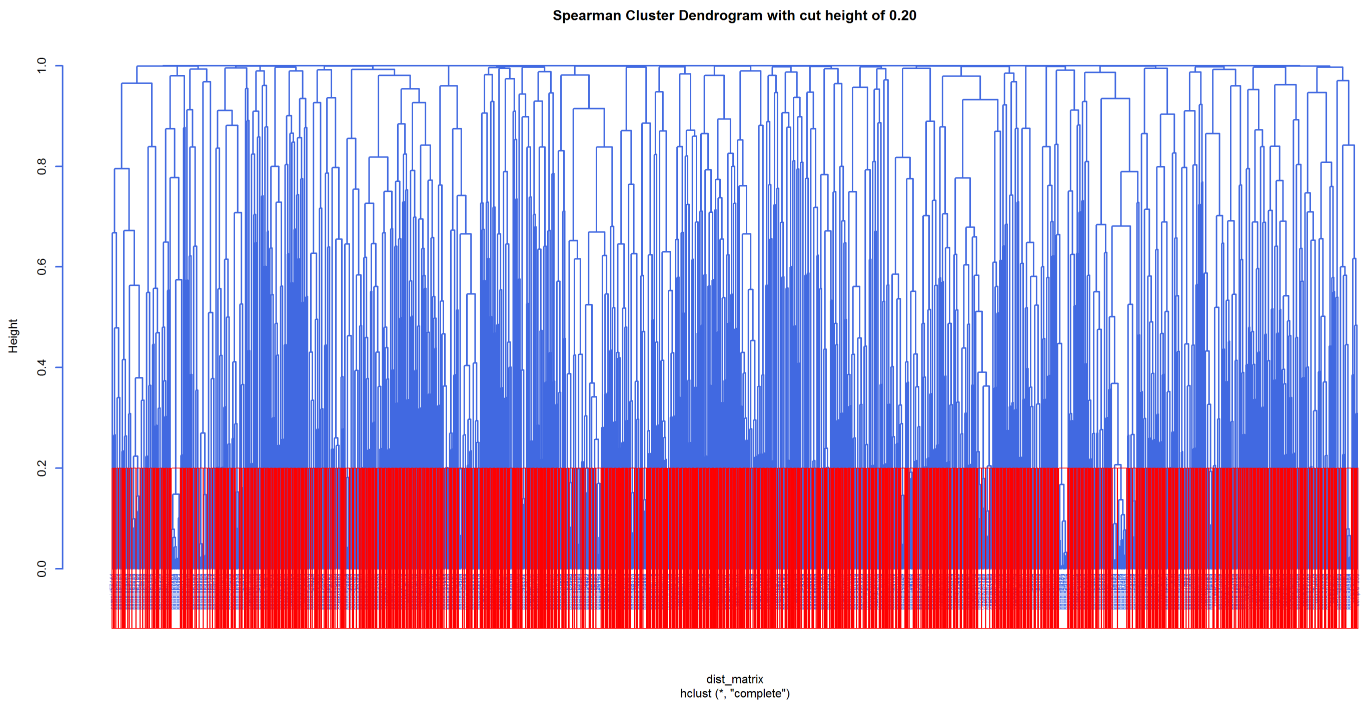


Total number of features with <20% missingness (used for identifying independent features): 927
Number of independent features used for PCA (based on a tree cut height of 0.20): 775

#### Generate PCs

```
[1] "Round 1 PCA - variance explained:"
```

```
ppca calculated PCA
Importance of component(s):
                   PC1    PC2     PC3     PC4     PC5     PC6    PC7     PC8
R2             0.06855 0.0543 0.04121 0.03791 0.03359 0.02894 0.0236 0.02123
Cumulative R2 0.06855 0.1228 0.16405 0.20196 0.23555 0.26450 0.2881 0.30933
                   PC9    PC10    PC11    PC12    PC13   PC14    PC15    PC16
R2             0.01976 0.01848 0.01561 0.01515 0.01417 0.0121 0.01178 0.01108
Cumulative R2 0.32909 0.34757 0.36318 0.37833 0.39249 0.4046 0.41638 0.42746
                  PC17    PC18    PC19     PC20
R2             0.01068 0.01018 0.00987 0.009634
Cumulative R2 0.43815 0.44833 0.45820 0.467837
```

```
 PC1  PC2  PC3  PC4  PC5  PC6  PC7  PC8  PC9 PC10 PC11 PC12 PC13 PC14 PC15 PC16
0.07 0.05 0.04 0.04 0.03 0.03 0.02 0.02 0.02 0.02 0.02 0.02 0.01 0.01 0.01 0.01
PC17 PC18 PC19 PC20
0.01 0.01 0.01 0.01
```

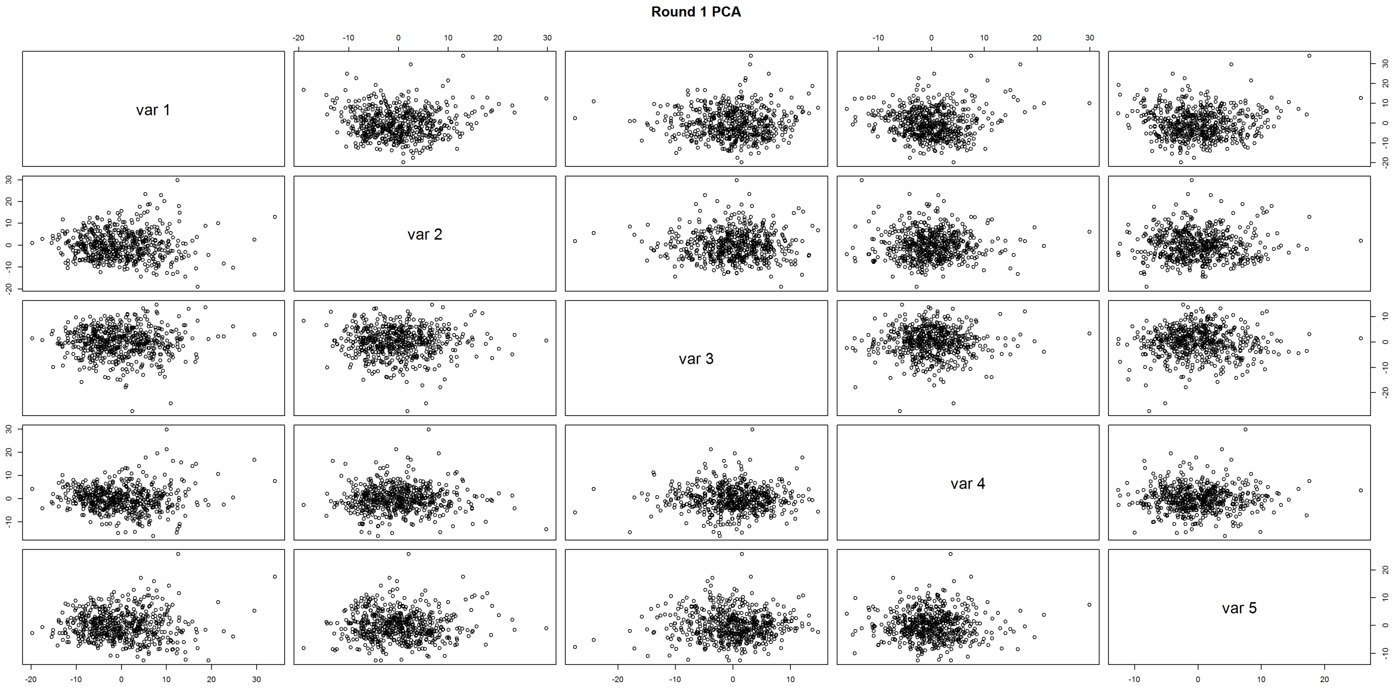


PCA Round 1 - Number of samples to exclude based on PC1 (+/- 5sd from the mean): 0
PCA Round 1 - Number of samples to exclude based on PC2 (+/- 5sd from the mean): 0

```
[1] "No PCA outliers identified. Second round of PCA not needed"
```

PCA Round 2 - Number of samples to exclude based on PC1 (+/- 5sd from the mean): 0
PCA Round 2 - Number of samples to exclude based on PC2 (+/- 5sd from the mean): 0

### Prepare QC'd datasets

```
Warning in orig_sample$client.identifier == extra_fails: longer object length is
not a multiple of shorter object length
```

No. of samples excluded due to failing Nightingale QC: 3

### Data overview post QC

The QC'd data files contain 571 samples and 1254 features.

### Check distributions (based on cleaned OrigScale data)

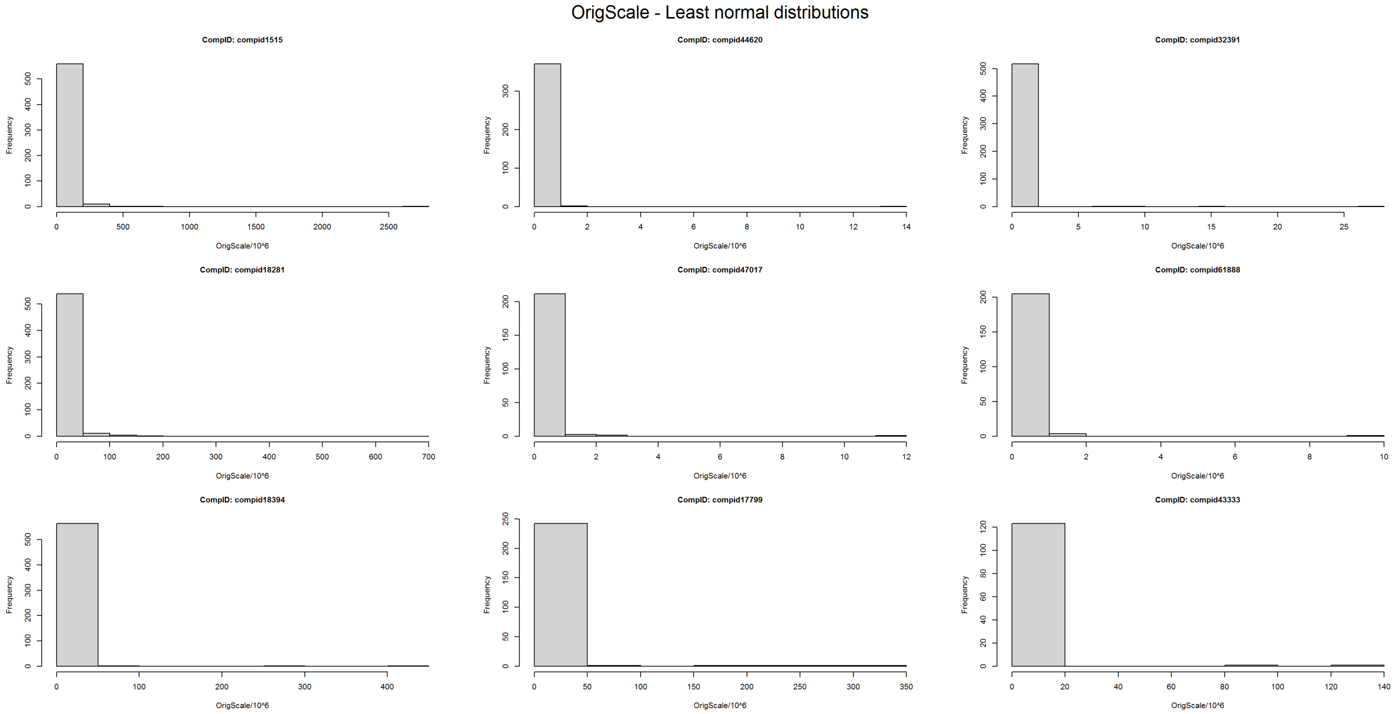


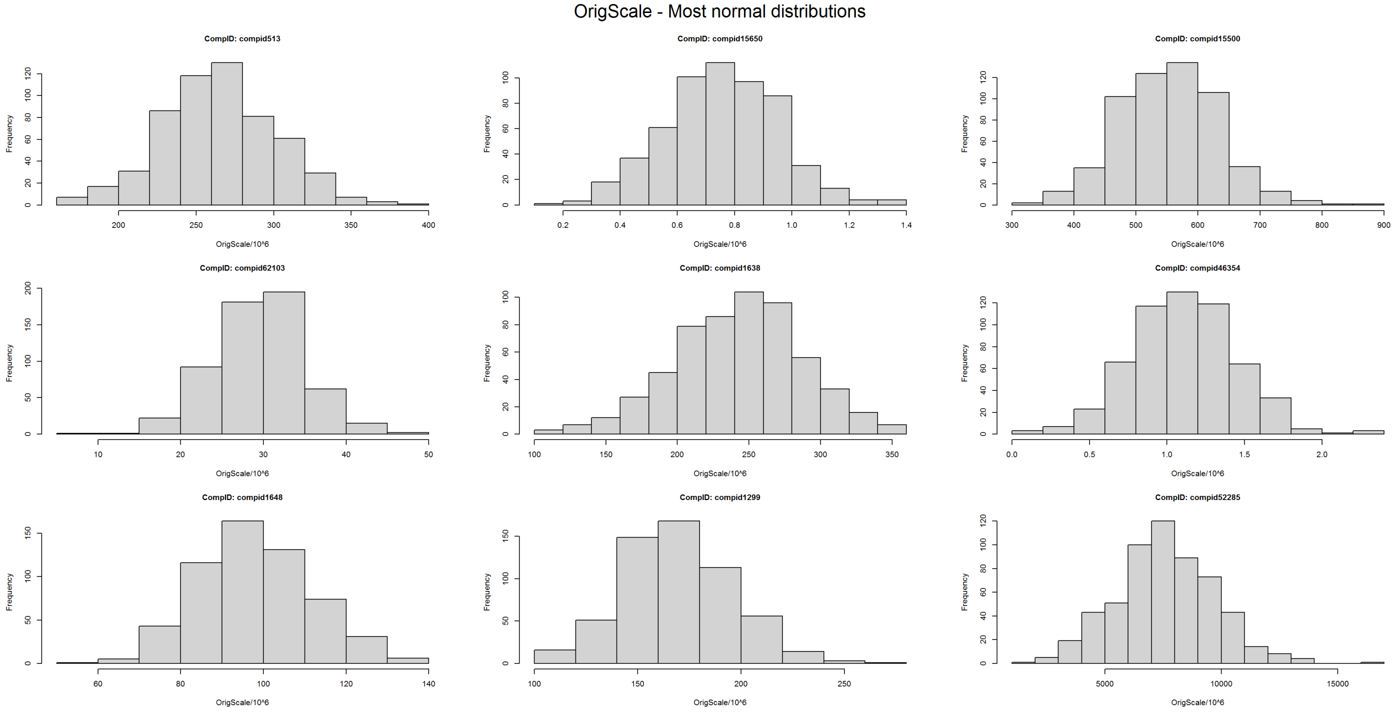


Proportion of features with a normal distribution (w>0.95): 0.1810207

Proportion of features with a normal distribution (p>0.01): 0.0199362

### Summarise missingness (post QC)

```
[1] "Summary of sample missingness:"
```

```
   Min. 1st Qu.  Median    Mean 3rd Qu.    Max. 
 0.1029  0.1380  0.1531  0.1561  0.1707  0.2496 
```

```
[1] "Summary of feature missingness:"
```

```
   Min. 1st Qu.  Median    Mean 3rd Qu.    Max. 
0.00000 0.00000 0.01051 0.15610 0.21629 0.99124 
```

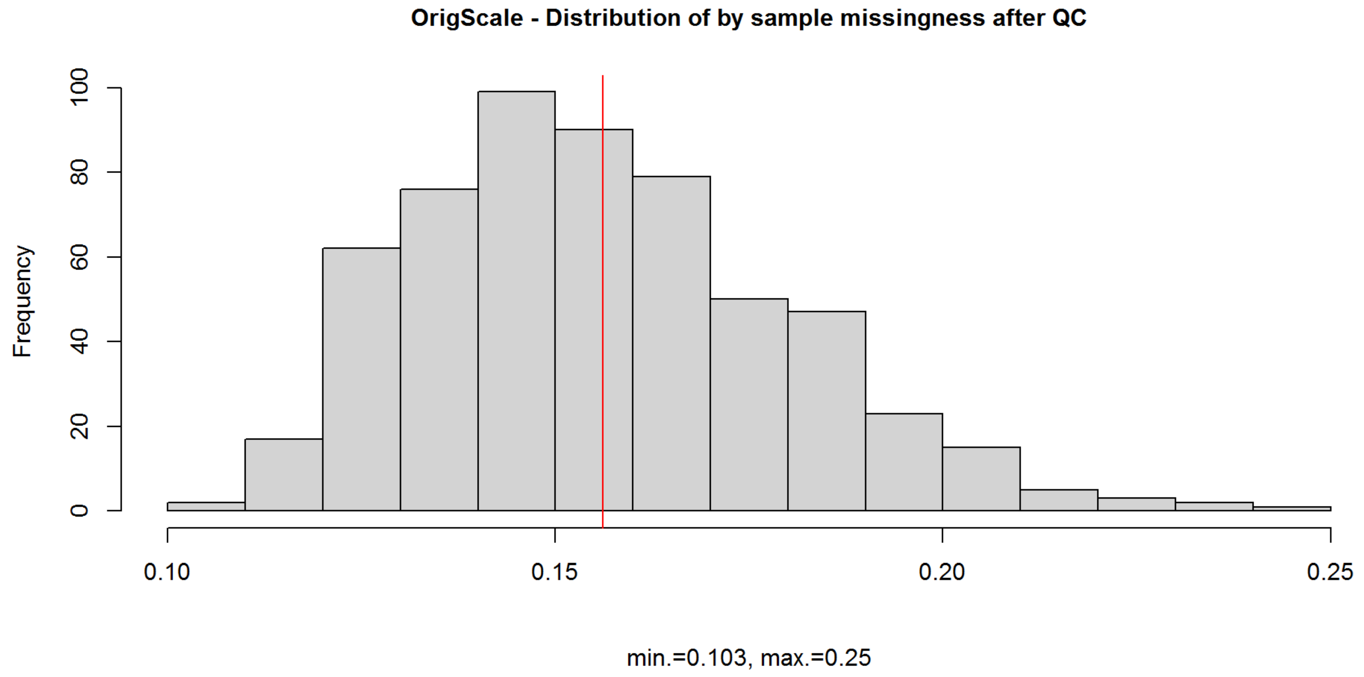


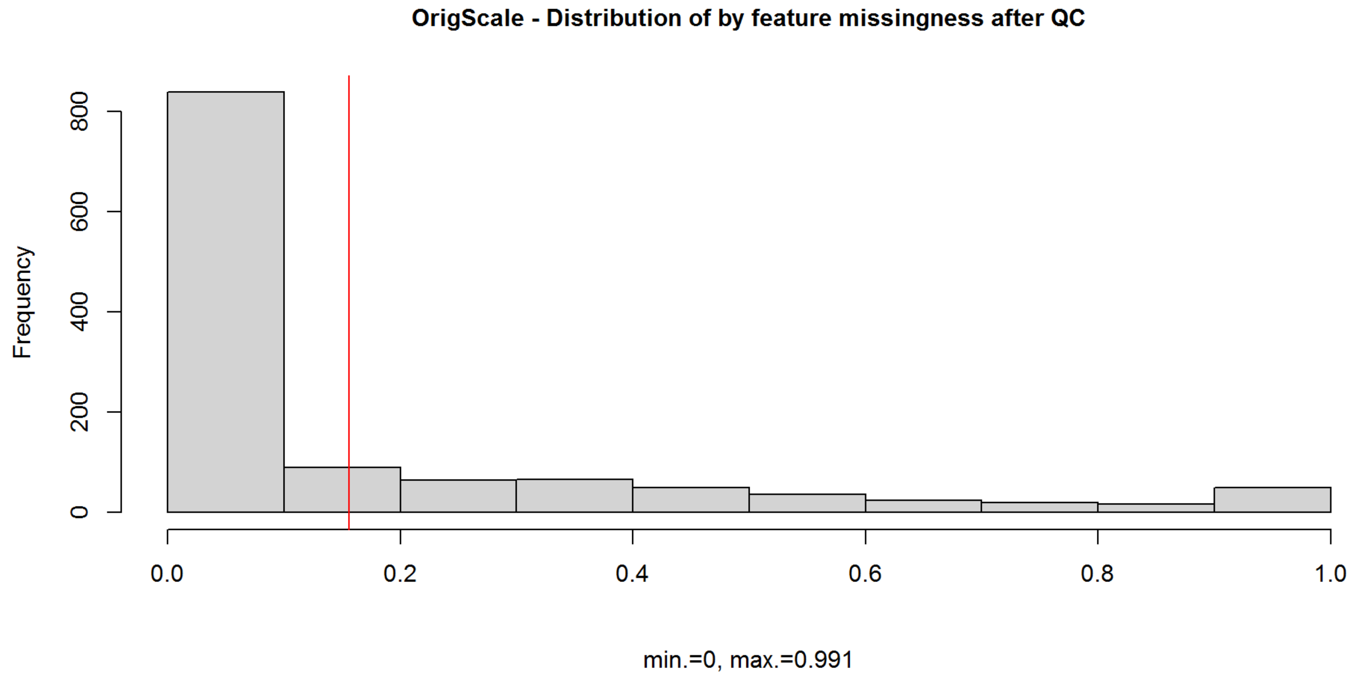


### Summarise outliers (post QC)

### Outliers are defined both as values greater than or less than 5SD from the mean and as values outside the 1st/99th percentile.

```
[1] "Summary of SD outliers by sample (post filtering):"
```

```
   Min. 1st Qu.  Median    Mean 3rd Qu.    Max. 
  0.000   0.000   2.000   3.324   4.000  35.000 
```

```
[1] "Summary of SD outliers by feature (post filtering):"
```

```
   Min. 1st Qu.  Median    Mean 3rd Qu.    Max. 
  0.000   0.000   1.000   1.514   2.000   8.000 
```

```
[1] "Summary of percentile outliers by sample (post QC):"
```

```
   Min. 1st Qu.  Median    Mean 3rd Qu.    Max. 
   2.00   12.00   19.00   22.97   28.00  102.00 
```

```
[1] "Summary of percentile outliers by feature (postQC):"
```

```
   Min. 1st Qu.  Median    Mean 3rd Qu.    Max. 
   2.00   10.00   12.00   10.46   12.00   12.00 
```

```
sessionInfo()
```

```
## R version 4.0.2 (2020-06-22)
## Platform: x86_64-w64-mingw32/x64 (64-bit)
## Running under: Windows Server 2012 R2 x64 (build 9600)
##
## Matrix products: default
##
## locale:
## [1] LC_COLLATE=English_United Kingdom.1252
## [2] LC_CTYPE=English_United Kingdom.1252
## [3] LC_MONETARY=English_United Kingdom.1252
## [4] LC_NUMERIC=C
## [5] LC_TIME=English_United Kingdom.1252
##
## attached base packages:
## [1] parallel  stats     graphics  grDevices utils     datasets  methods
## [8] base
##
## other attached packages:
## [1] pcaMethods_1.80.0   Biobase_2.48.0      BiocGenerics_0.34.0
## [4] psych_2.0.7         ggplot2_3.3.5       data.table_1.14.2
## [7] pwr_1.3-0           knitr_1.36          dplyr_1.0.7
##
## loaded via a namespace (and not attached):
##  [1] Rcpp_1.0.7       highr_0.9        pillar_1.6.4     compiler_4.0.2
##  [5] jquerylib_0.1.4  tools_4.0.2      digest_0.6.28    nlme_3.1-149
##  [9] lattice_0.20-41  evaluate_0.14    lifecycle_1.0.1  tibble_3.1.5
## [13] gtable_0.3.0     pkgconfig_2.0.3  rlang_0.4.11     yaml_2.2.1
## [17] xfun_0.26        fastmap_1.1.0    withr_2.4.2      stringr_1.4.0
## [21] generics_0.1.0   vctrs_0.3.8      grid_4.0.2       tidyselect_1.1.1
## [25] glue_1.4.2       R6_2.5.1         fansi_0.5.0      rmarkdown_2.11
## [29] farver_2.1.0     purrr_0.3.4      magrittr_2.0.1   scales_1.1.1
## [33] ellipsis_0.3.2   htmltools_0.5.2  mnormt_2.0.1     colorspace_2.0-2
## [37] utf8_1.2.2       stringi_1.7.5    munsell_0.5.0    tmvnsim_1.0-2
## [41] crayon_1.4.1
```
