## Supplementary Document 3 for "The metabolomic signature of weight loss in the Diabetes Remission Clinical Trial (DiRECT)"

### Nightingale NMR Metabolite Data QC Report

L.J.Corbin

31/03/2022

#### Data overview

Number of samples in OrigScale file: 574
Number of features in OrigScale file: 227
Number of raw measures: 148
Number of derived measures: 79

#### Power exploration for case/control analysis

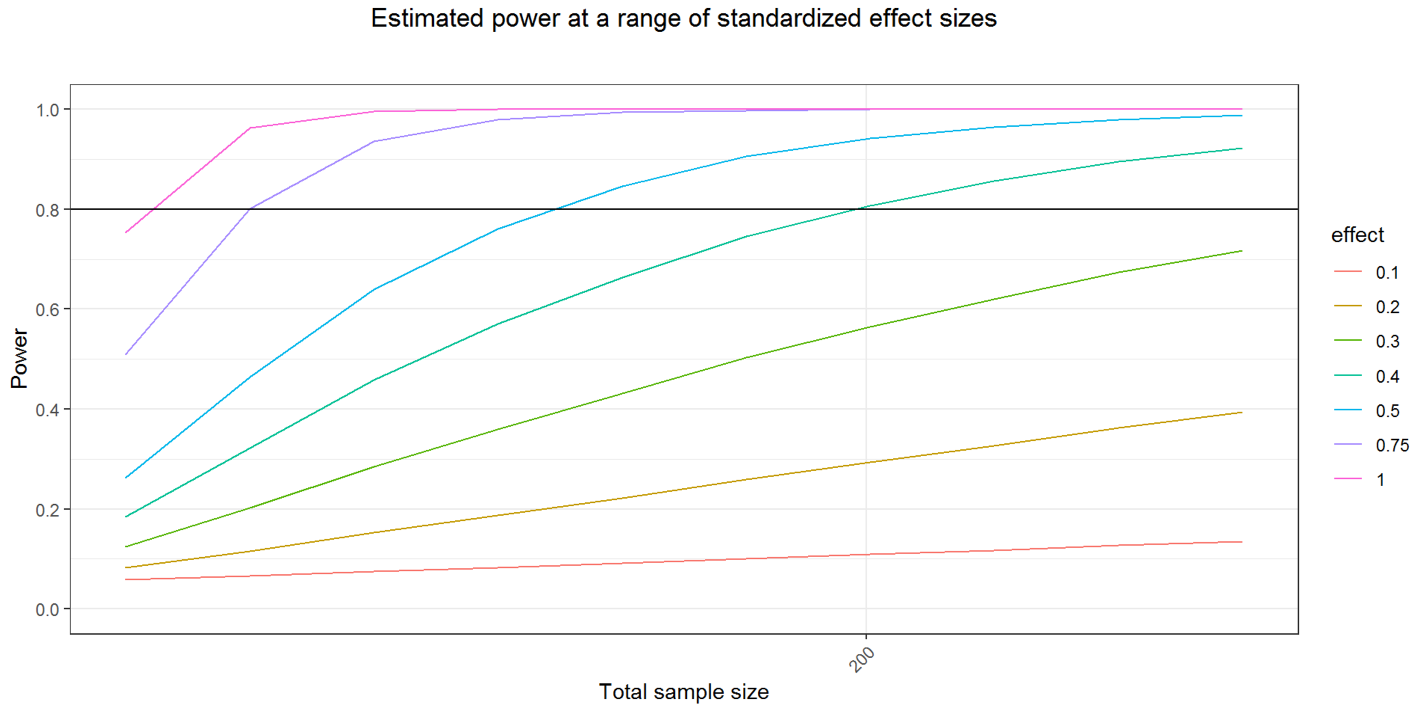


#### Power exploration for continuous outcome analysis

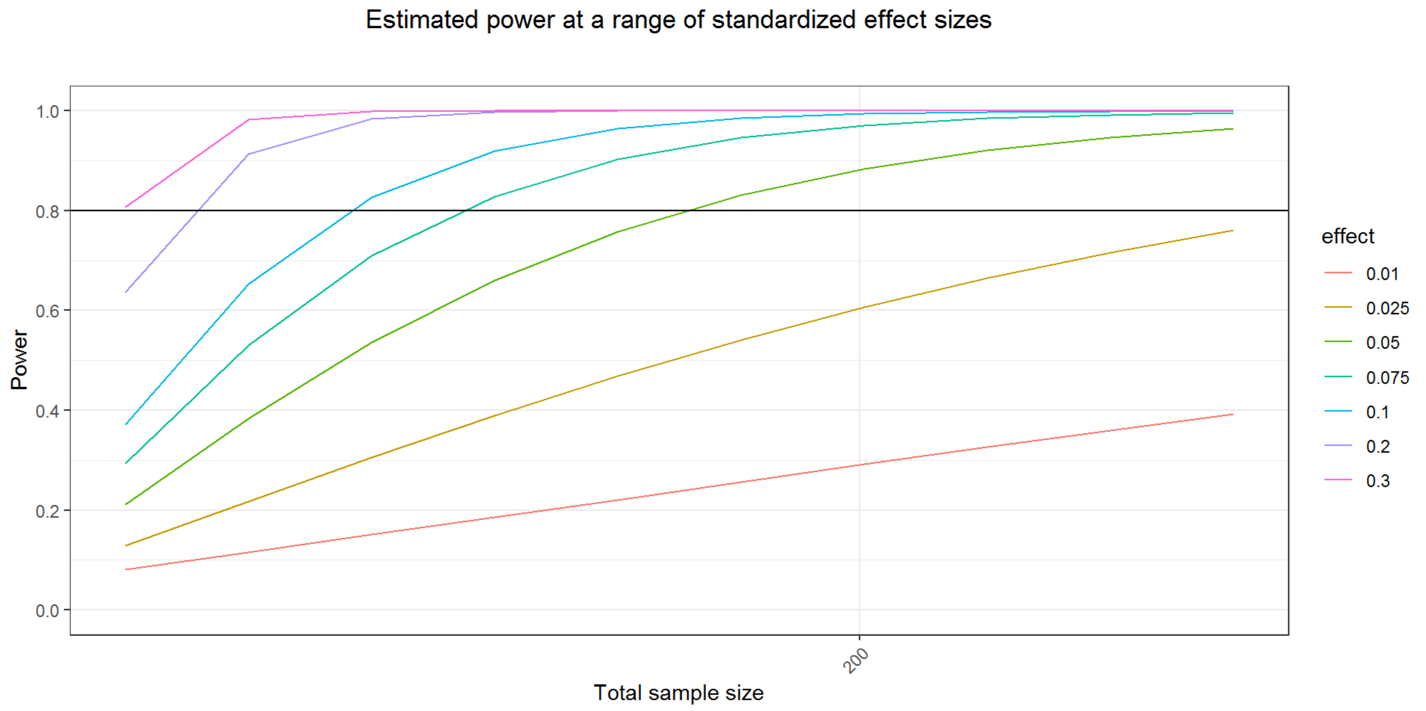


### Summarise missingness (pre filtering)

By sample missingness is calculated after excluding derived measures (i.e. percentages and ratios)

```
[1] "Summary of sample missingness:"
```

```
    Min.  1st Qu.   Median     Mean  3rd Qu.     Max. 
0.000000 0.000000 0.006757 0.006486 0.006757 0.351351 
```

```
[1] "Summary of feature missingness:"
```

```
    Min.  1st Qu.   Median     Mean  3rd Qu.     Max. 
0.000000 0.000000 0.003484 0.019977 0.005227 0.677700 
```

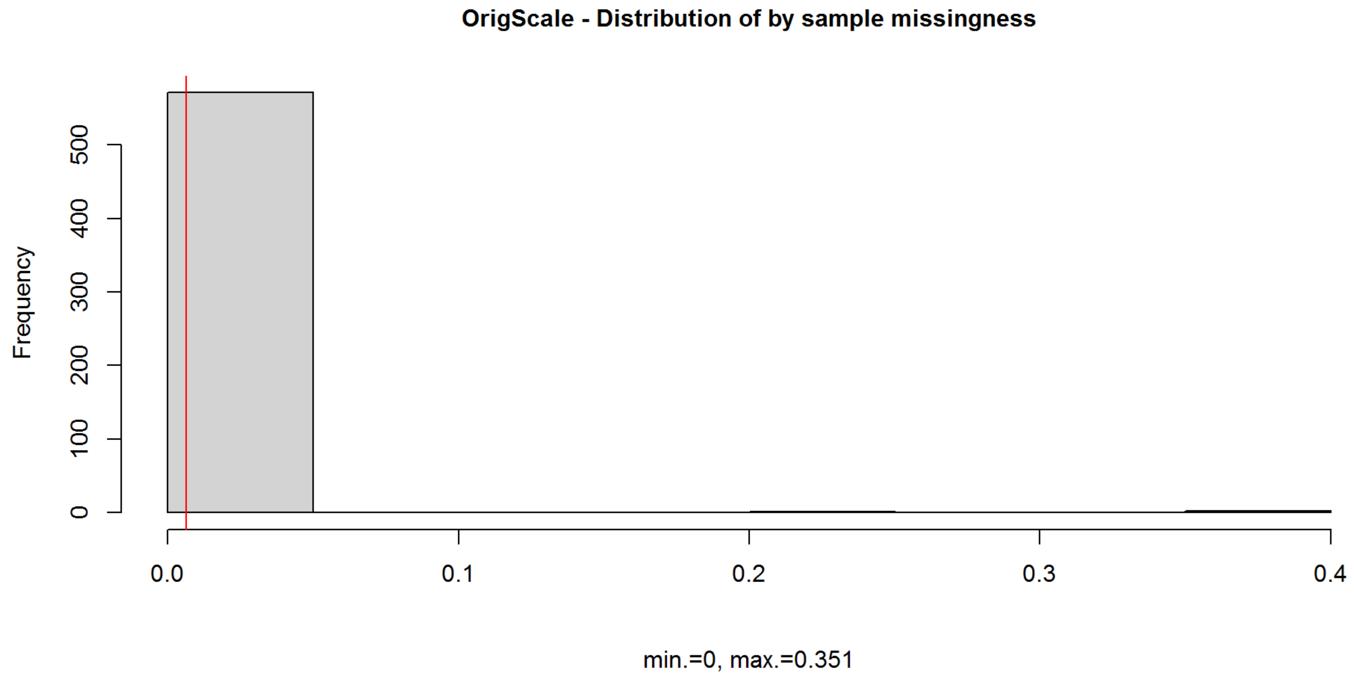


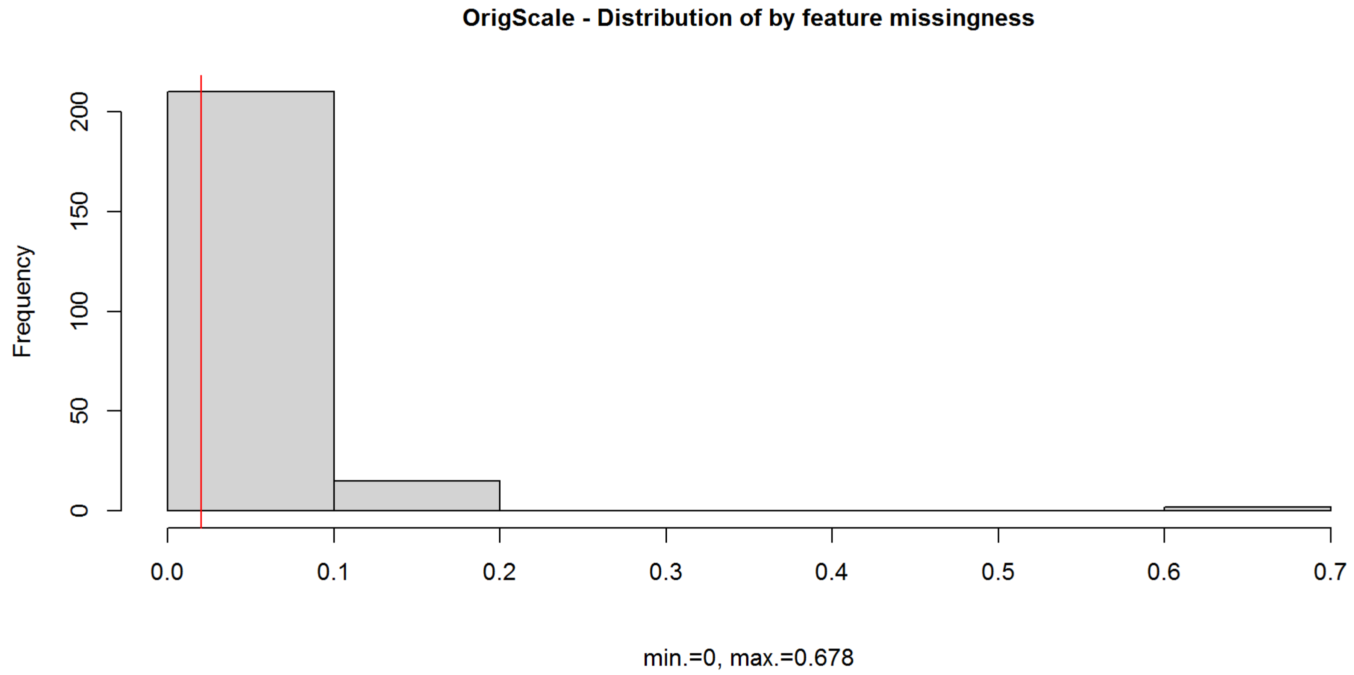


There are 96 features with no data missing.
Of these, 96 are raw features (not derived).

#### Example sample numbers at different missingness rates

If feature has 10% missingness, data is available for: 516.6 samples.
If feature has 20% missingness, data is available for: 459.2 samples.
If feature has 30% missingness, data is available for: 401.8 samples.
If feature has 40% missingness, data is available for: 344.4 samples.
If feature has 50% missingness, data is available for: 287 samples.

### Summarise outliers (pre filtering)

#### Outliers are defined both as values greater than or less than 5SD from the mean and as values outside the 1st/99th percentile.

```
[1] "Summary of SD outliers by sample (pre filtering):"
```

```
   Min. 1st Qu.  Median    Mean 3rd Qu.    Max. 
 0.0000  0.0000  0.0000  0.2561  0.0000 33.0000 
```

```
[1] "Summary of SD outliers by feature (pre filtering):"
```

```
   Min. 1st Qu.  Median    Mean 3rd Qu.    Max. 
 0.0000  0.0000  0.0000  0.6476  1.0000  5.0000 
```

```
[1] "Summary of percentile outliers by sample (pre filtering):"
```

```
   Min. 1st Qu.  Median    Mean 3rd Qu.    Max. 
  0.000   0.000   1.000   8.077   9.000 101.000 
```

```
[1] "Summary of percentile outliers by feature (pre filtering):"
```

```
   Min. 1st Qu.  Median    Mean 3rd Qu.    Max. 
   4.00   12.00   12.00   20.42   13.00  100.00 
```

### Apply missingness thresholds to data and re-summarise

Samples excluded if more than 20% of features are missing.
By sample missingness is calculated after excluding derived measures (i.e. percentages and ratios).
Features excluded if they are missing in more than 20% samples.

Number of sample exclusions based on >20% missingness: 3
Number of feature exclusions based on >20% missingness: 2

```
[1] "Summary of sample missingness after cleaning:"
```

```
     Min.   1st Qu.    Median      Mean   3rd Qu.      Max. 
0.0000000 0.0000000 0.0000000 0.0003336 0.0000000 0.0136054 
```

```
[1] "Summary of feature missingness after cleaning:"
```

```
   Min. 1st Qu.  Median    Mean 3rd Qu.    Max. 
0.00000 0.00000 0.00000 0.01189 0.00000 0.14536 
```

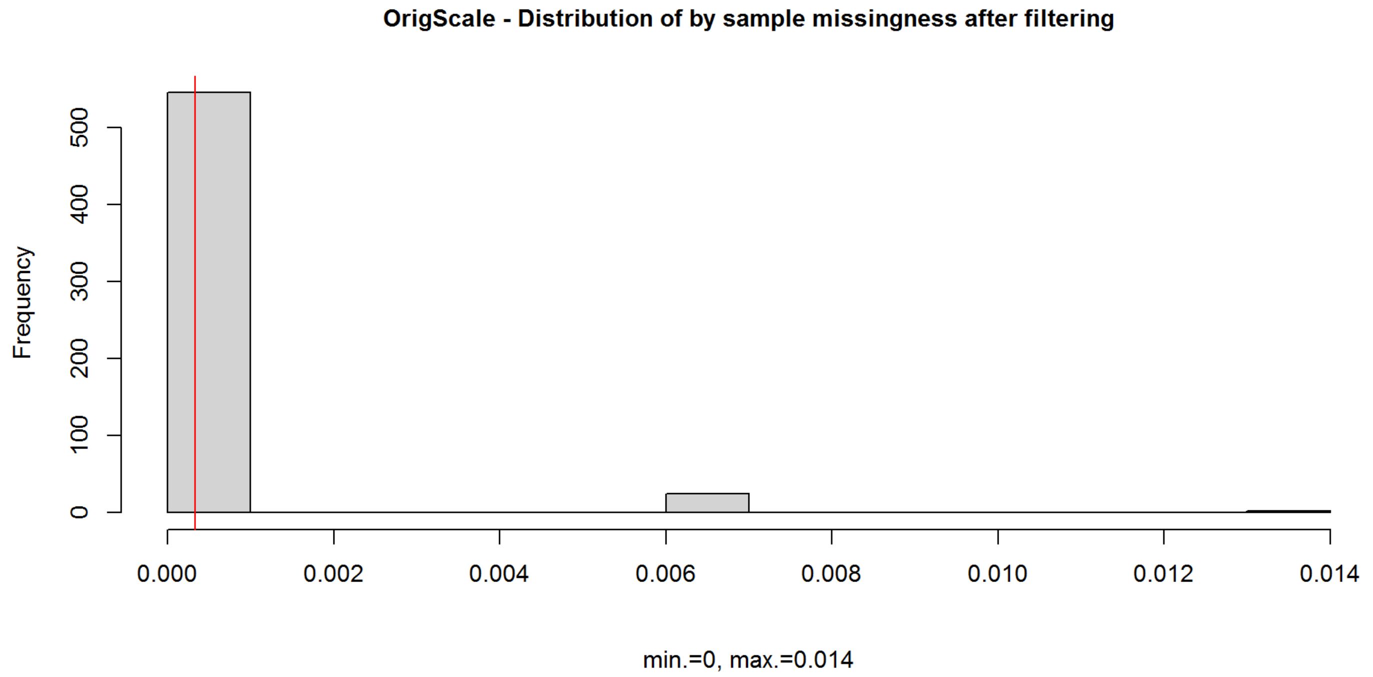


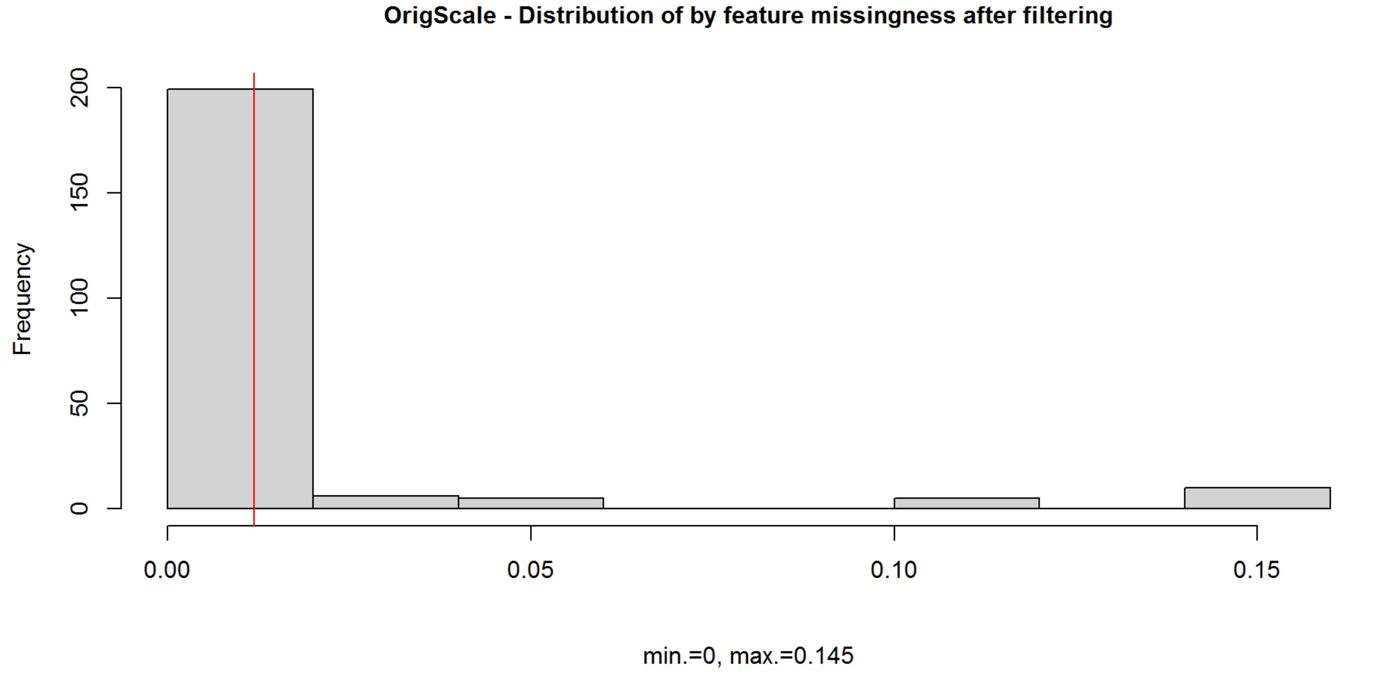


### Total metabolite levels (by sample) (based on filtered data)

Calculated after excluding derived measures (i.e. percentages and ratios).

```
[1] "Summary of total peak area:"
```

```
   Min. 1st Qu.  Median    Mean 3rd Qu.    Max. 
  7.535  14.562  16.837  17.287  19.765  33.992 
```

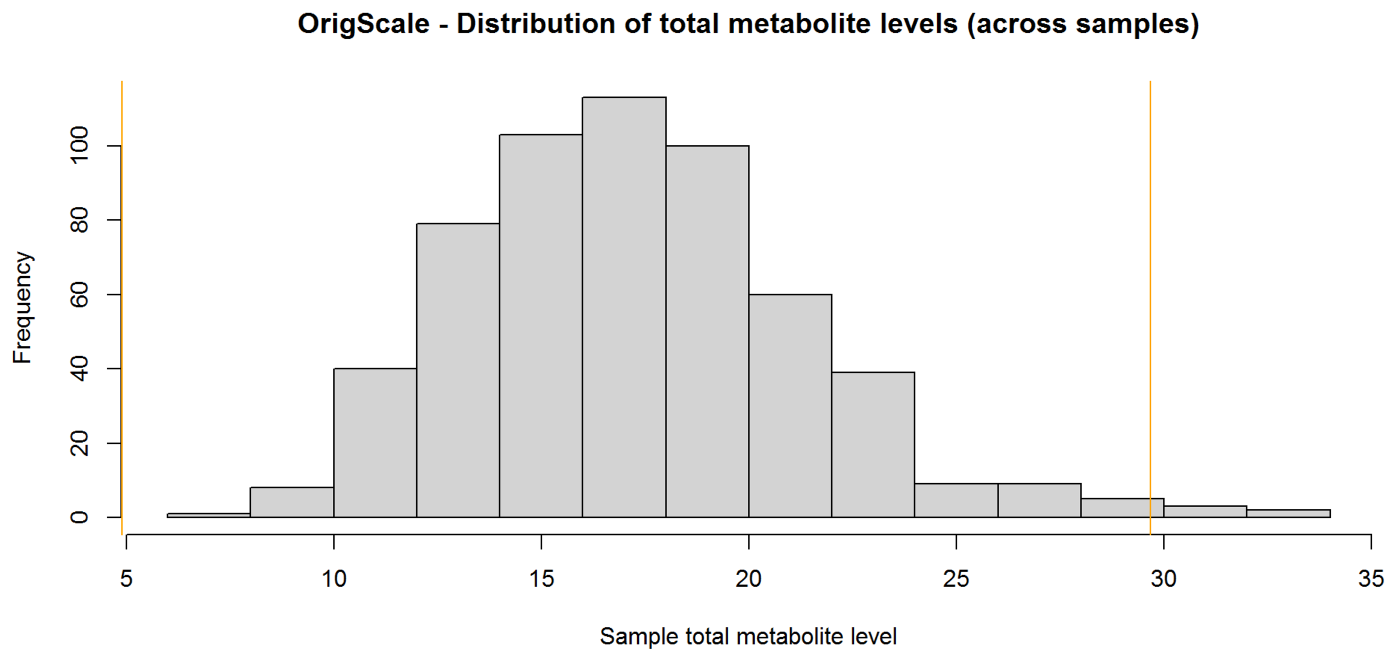


Number of sample exclusions based on total metabolite level (+/- 3sd from the mean): 7
Number of sample exclusions based on total metabolite level (+/- 5sd from the mean) (current criteria): 0

### Plot principal components - check for sample outliers

#### Identify independent features based on Spearman's correlation

Tree generated after excluding derived measures (i.e. percentages and ratios).

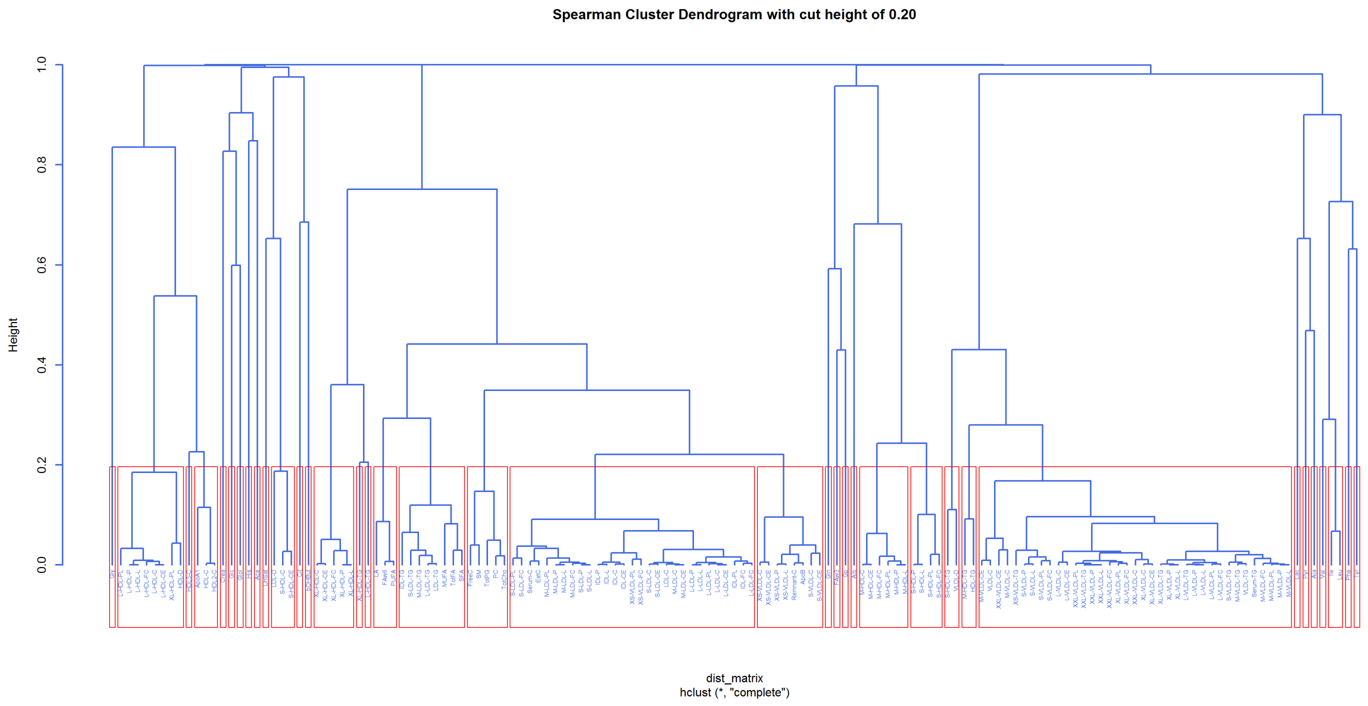


Total number of raw features: 147
Number of independent features (based on a tree cut height of 0.20): 37

#### Generate PCs

PCA run on unique features from tree with complete data (i.e. no missing data points).

```
[1] "Round 1 PCA - variance explained:"
```

```
ppca calculated PCA
Importance of component(s):
                 PC1    PC2    PC3     PC4     PC5     PC6     PC7     PC8
R2            0.2517 0.1364 0.1076 0.06138 0.05788 0.04691 0.04091 0.03654
Cumulative R2 0.2517 0.3881 0.4957 0.55709 0.61497 0.66188 0.70278 0.73932
                  PC9    PC10    PC11    PC12    PC13    PC14    PC15    PC16
R2            0.03498 0.02909 0.02482 0.02174 0.02073 0.01844 0.01598 0.01371
Cumulative R2 0.77430 0.80339 0.82821 0.84995 0.87067 0.88912 0.90509 0.91881
                 PC17    PC18    PC19     PC20
R2            0.01287 0.01253 0.01053 0.009075
Cumulative R2 0.93167 0.94421 0.95474 0.963812
```

```
 PC1  PC2  PC3  PC4  PC5  PC6  PC7  PC8  PC9 PC10 PC11 PC12 PC13 PC14 PC15 PC16
0.25 0.14 0.11 0.06 0.06 0.05 0.04 0.04 0.03 0.03 0.02 0.02 0.02 0.02 0.02 0.01
PC17 PC18 PC19 PC20
0.01 0.01 0.01 0.01
```

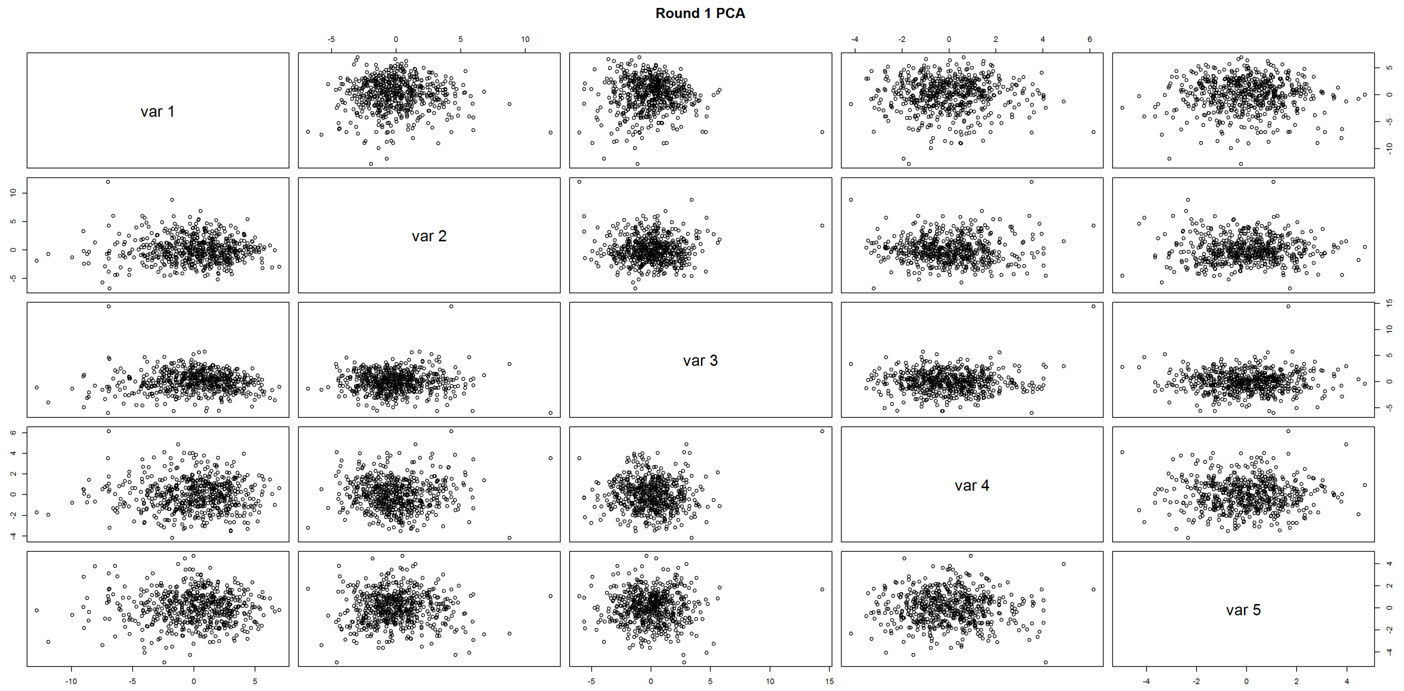


PCA Round 1 - Number of samples to exclude based on PC1 (+/- 5sd from the mean): 0

PCA Round 1 - Number of samples to exclude based on PC2 (+/- 5sd from the mean): 1

```
[1] "Round 2 PCA - variance explained:"
ppca calculated PCA
Importance of component(s):
                 PC1    PC2    PC3     PC4     PC5     PC6    PC7    PC8
R2            0.2532 0.1333 0.1070 0.06182 0.05817 0.04717 0.0408 0.0369
Cumulative R2 0.2532 0.3865 0.4935 0.55528 0.61345 0.66063 0.7014 0.7383
                  PC9    PC10    PC11    PC12    PC13    PC14    PC15    PC16
R2            0.03525 0.02919 0.02492 0.02179 0.02071 0.01846 0.01599 0.01376
Cumulative R2 0.77357 0.80276 0.82768 0.84947 0.87018 0.88864 0.90463 0.91839
                PC17   PC18    PC19     PC20
R2            0.0129 0.0126 0.01053 0.009142
Cumulative R2 0.9313 0.9439 0.95443 0.963568
```

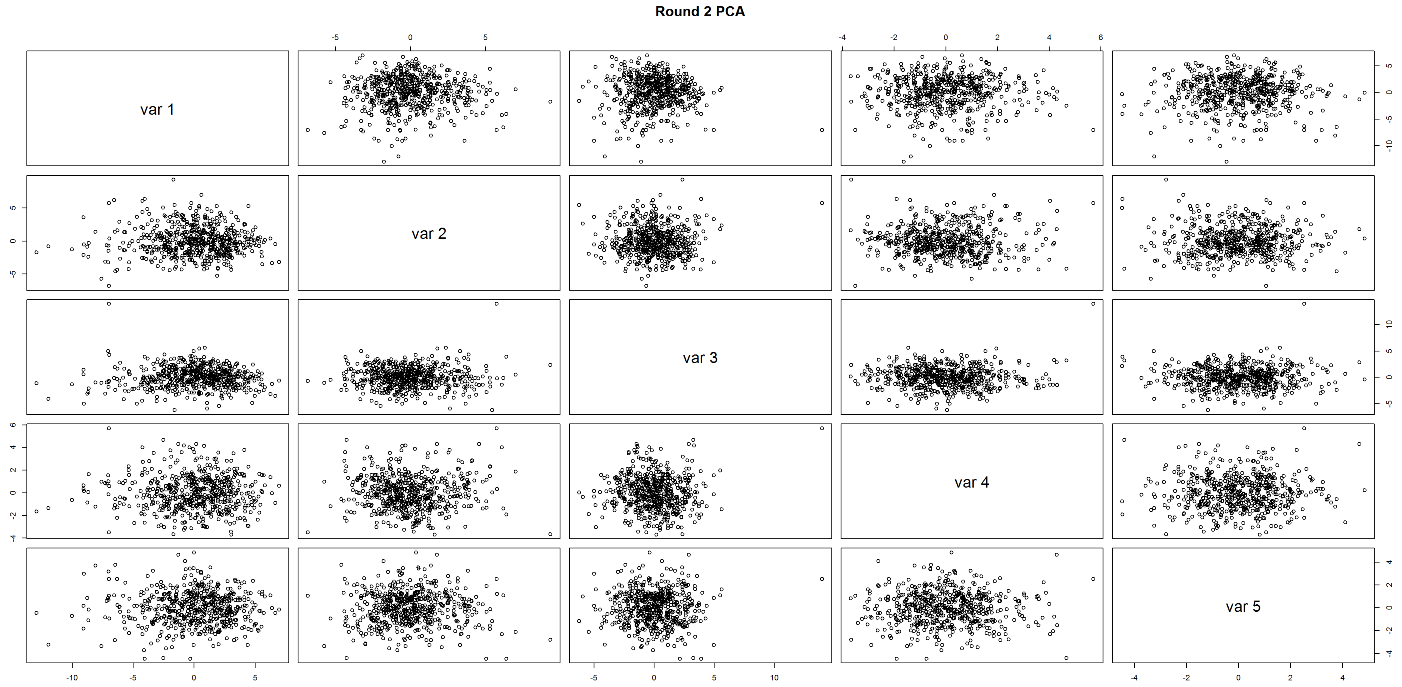


PCA Round 2 - Number of samples to exclude based on PC1 (+/- 5sd from the mean): 0
PCA Round 2 - Number of samples to exclude based on PC2 (+/- 5sd from the mean): 0

### Prepare QC’d datasets

No. of samples excluded due to failing Nightingale QC: 3

### Data overview post QC

The QC’d data files contain 567 samples and 225 features.

### Check distributions (based on cleaned data)

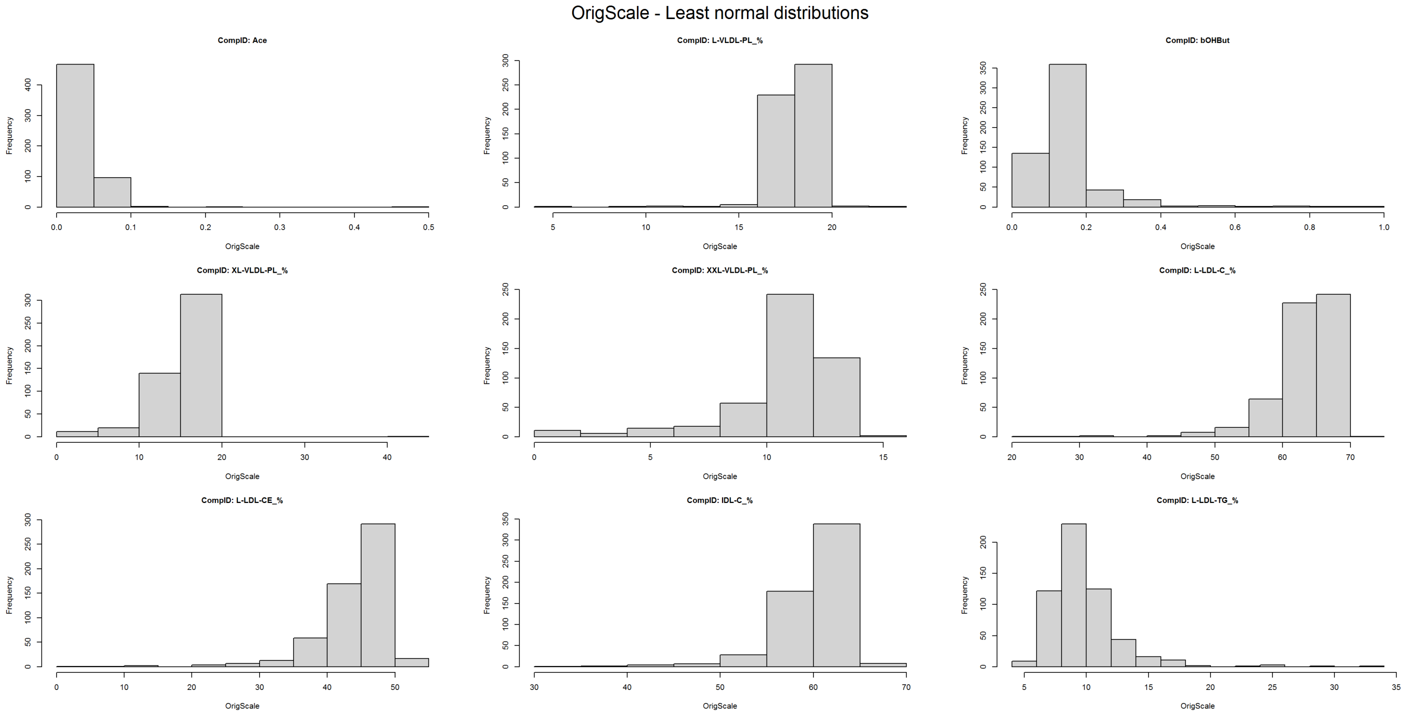


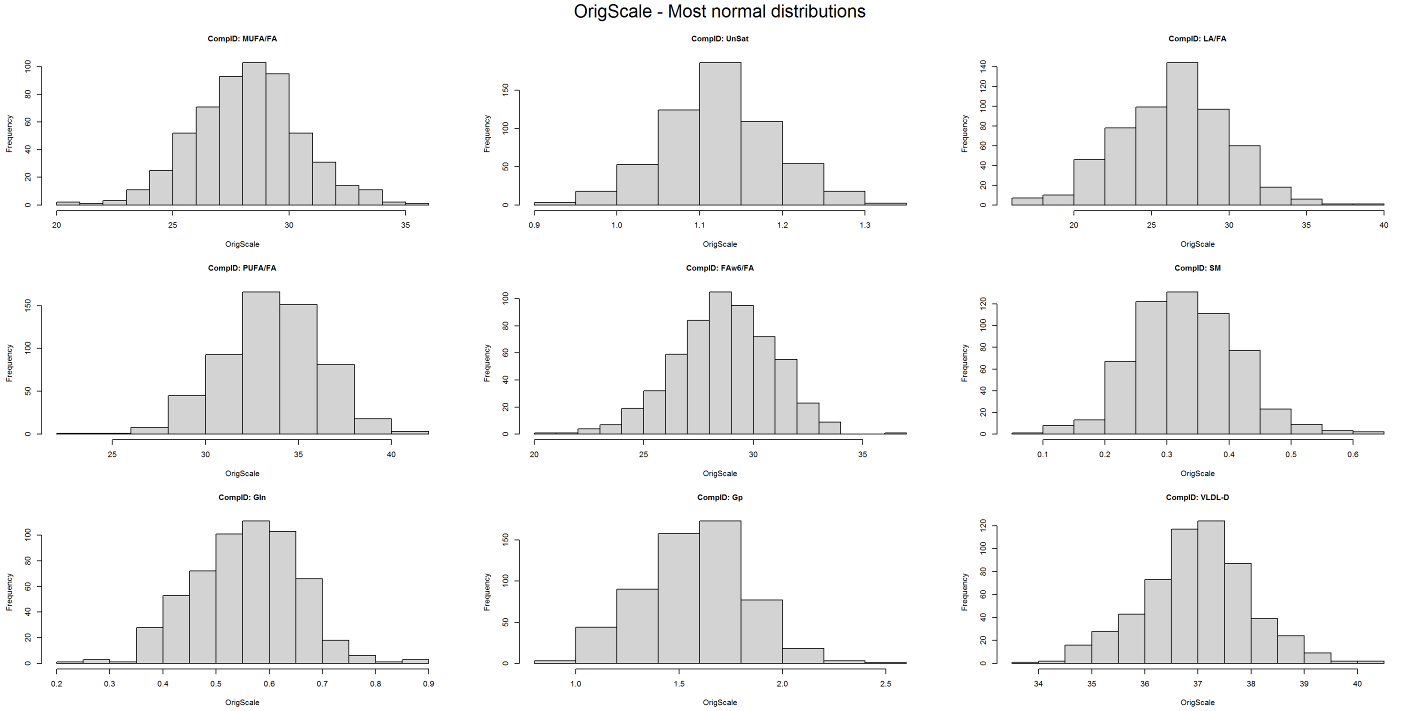


Proportion of features with a normal distribution (w>0.95): 0.6
Proportion of features with a normal distribution (p>0.01): 0.0488889

### Summarise missingness (post QC)

```
[1] "Summary of sample missingness:"
```

```
   Min. 1st Qu.  Median    Mean 3rd Qu.    Max. 
0.00000 0.00000 0.00000 0.01188 0.02222 0.09333 
```

```
[1] "Summary of feature missingness:"
```

```
   Min. 1st Qu.  Median    Mean 3rd Qu.    Max. 
0.00000 0.00000 0.00000 0.01188 0.00000 0.14462 
```

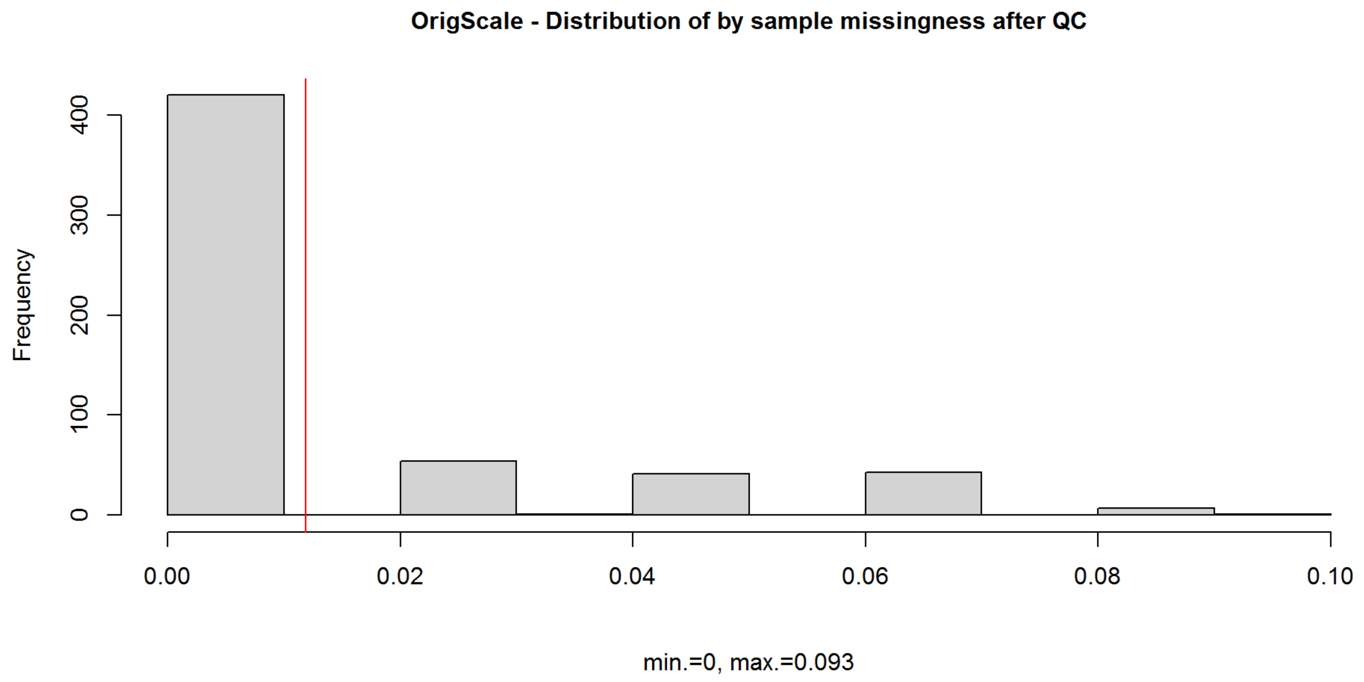


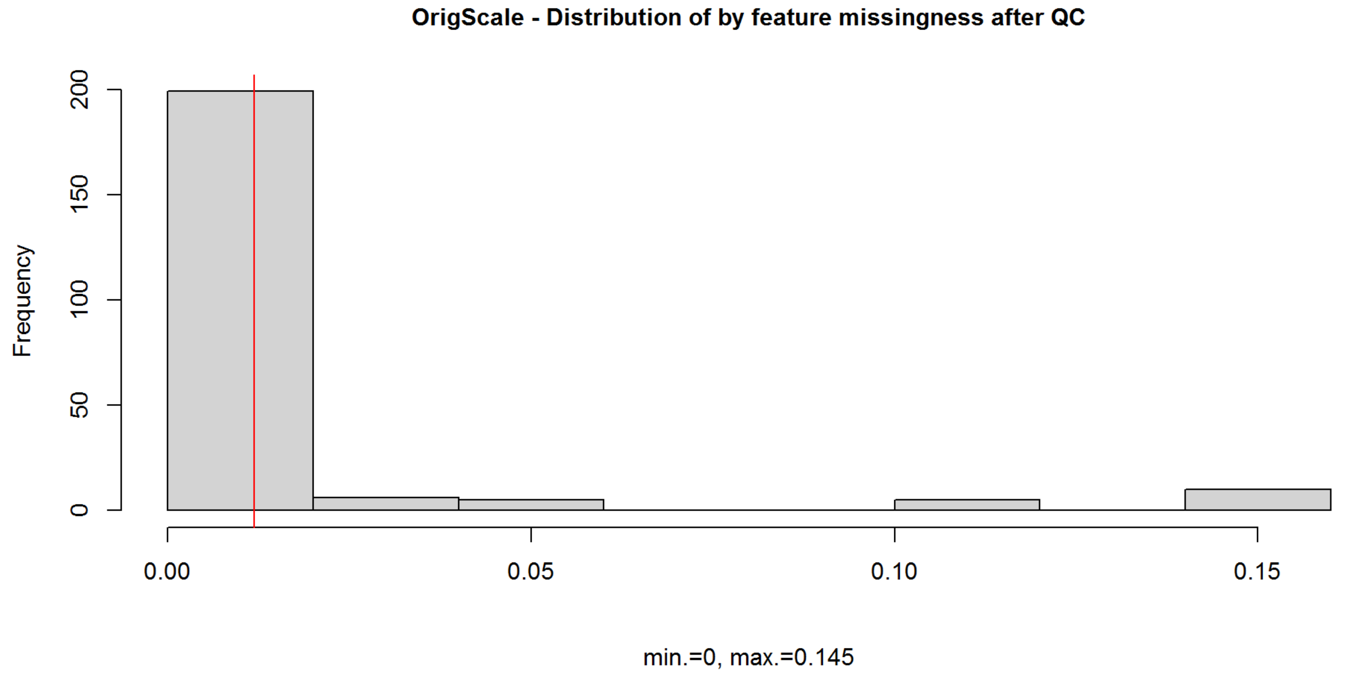


### Summarise outliers (post QC)

#### Outliers are defined both as values greater than or less than 5SD from the mean and as values outside the 1st/99th percentile.

```
[1] "Summary of SD outliers by sample (post filtering):"
```

```
   Min. 1st Qu.  Median    Mean 3rd Qu.    Max. 
 0.0000  0.0000  0.0000  0.2099  0.0000 33.0000 
```

```
[1] "Summary of SD outliers by feature (post filtering):"
```

```
   Min. 1st Qu.  Median    Mean 3rd Qu.    Max. 
 0.0000  0.0000  0.0000  0.5289  1.0000  5.0000 
```

```
[1] "Summary of percentile outliers by sample (post QC):"
```

```
   Min. 1st Qu.  Median    Mean 3rd Qu.    Max. 
  0.000   0.000   1.000   7.961  10.000 102.000 
```

```
[1] "Summary of percentile outliers by feature (postQC):"
```

```
   Min. 1st Qu.  Median    Mean 3rd Qu.    Max. 
  10.00   12.00   12.00   20.06   13.00   97.00 
```

```
sessionInfo()
```

```
## R version 4.0.2 (2020-06-22)
## Platform: x86_64-w64-mingw32/x64 (64-bit)
## Running under: Windows Server 2012 R2 x64 (build 9600)
## 
## Matrix products: default
## 
## locale:
## [1] LC_COLLATE=English_United Kingdom.1252 
## [2] LC_CTYPE=English_United Kingdom.1252   
## [3] LC_MONETARY=English_United Kingdom.1252
## [4] LC_NUMERIC=C                           
## [5] LC_TIME=English_United Kingdom.1252    
## 
## attached base packages:
## [1] parallel  stats     graphics  grDevices utils     datasets  methods  
## [8] base     
## 
## other attached packages:
## [1] pcaMethods_1.80.0   Biobase_2.48.0      BiocGenerics_0.34.0
## [4] psych_2.0.7         ggplot2_3.3.5       data.table_1.14.2  
## [7] pwr_1.3-0           knitr_1.36          dplyr_1.0.7        
## 
## loaded via a namespace (and not attached):
##  [1] Rcpp_1.0.7       highr_0.9        pillar_1.6.4     compiler_4.0.2  
##  [5] jquerylib_0.1.4  tools_4.0.2      digest_0.6.28    nlme_3.1-149    
##  [9] lattice_0.20-41  evaluate_0.14    lifecycle_1.0.1  tibble_3.1.5    
## [13] gtable_0.3.0     pkgconfig_2.0.3  rlang_0.4.11     yaml_2.2.1      
## [17] xfun_0.26        fastmap_1.1.0    withr_2.4.2      stringr_1.4.0   
## [21] generics_0.1.0   vctrs_0.3.8      grid_4.0.2       tidyselect_1.1.1
## [25] glue_1.4.2       R6_2.5.1         fansi_0.5.0      rmarkdown_2.11  
## [29] farver_2.1.0     purrr_0.3.4      magrittr_2.0.1   scales_1.1.1    
## [33] ellipsis_0.3.2   htmltools_0.5.2  mnormt_2.0.1     colorspace_2.0-2
## [37] utf8_1.2.2       stringi_1.7.5    munsell_0.5.0    tmvnsim_1.0-2   
## [41] crayon_1.4.1
```
