## Supplementary Document 4 - Figure S2A for "The metabolomic signature of weight loss in the Diabetes Remission Clinical Trial (DiRECT)"

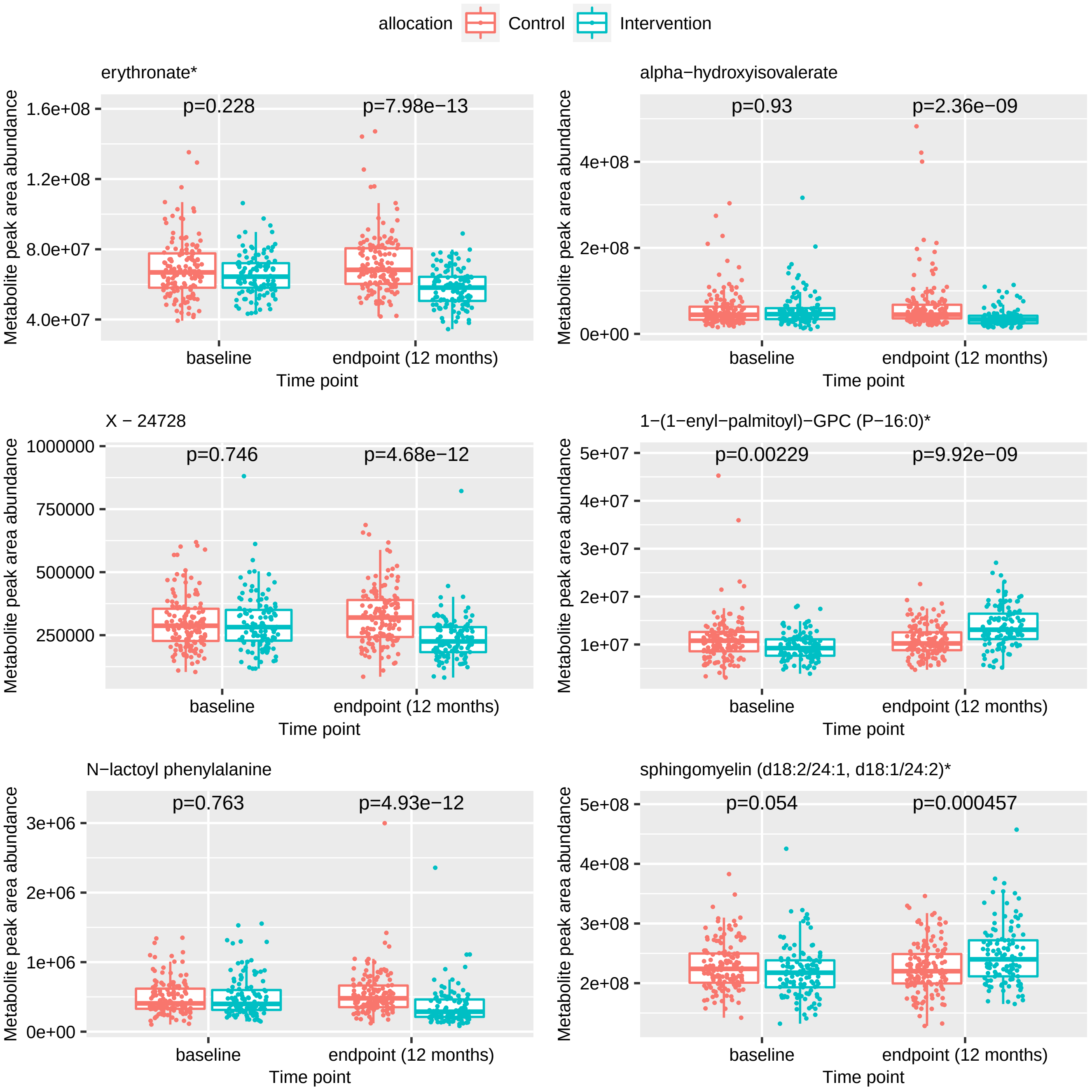

allocation
Control
Intervention
erythronate*
p=0.228
p=7.98e−13
1.6e+08
1.2e+08
8.0e+07
4.0e+07
Metabolite peak area abundance
baseline
endpoint (12 months)
Time point
alpha−hydroxyisovalerate
p=0.93
p=2.36e−09
4e+08
2e+08
0e+00
X − 24728
p=0.746
p=4.68e−12
1000000
750000
500000
250000
1−(1−enyl−palmitoyl)−GPC (P−16:0)*
p=0.00229
p=9.92e−09
5e+07
4e+07
3e+07
2e+07
1e+07
N−lactoyl phenylalanine
p=0.763
p=4.93e−12
3e+06
2e+06
1e+06
0e+00
sphingomyelin (d18:2/24:1, d18:1/24:2)*
p=0.054
p=0.000457
5e+08
4e+08
3e+08
2e+08

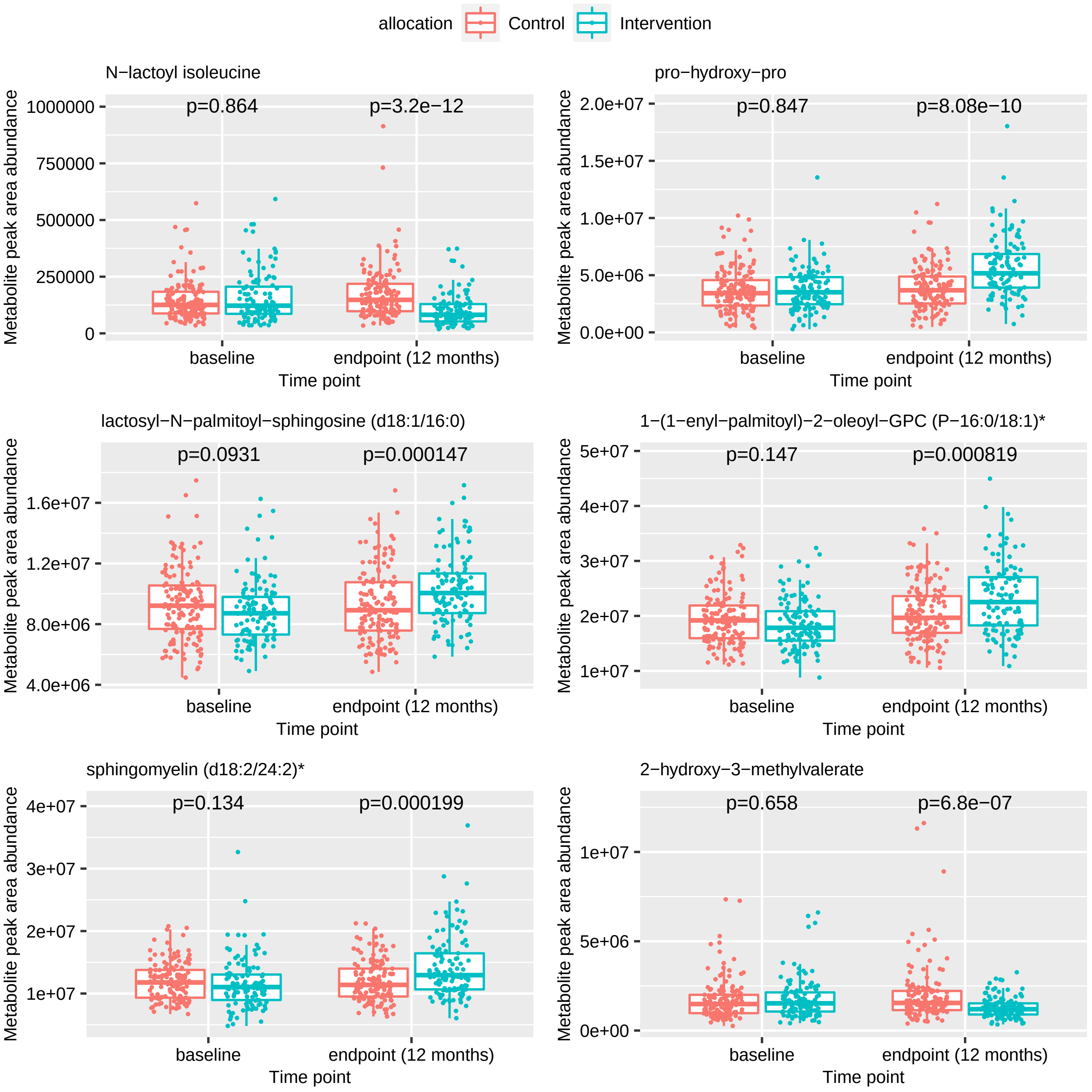

allocation
Control
Intervention
N−lactoyl isoleucine
p=0.864
p=3.2e−12
1000000
750000
500000
250000
0
pro−hydroxy−pro
p=0.847
p=8.08e−10
2.0e+07
1.5e+07
1.0e+07
5.0e+06
0.0e+00
lactosyl−N−palmitoyl−sphingosine (d18:1/16:0)
p=0.0931
p=0.000147
1.6e+07
1.2e+07
8.0e+06
4.0e+06
1−(1−enyl−palmitoyl)−2−oleoyl−GPC (P−16:0/18:1)*
p=0.147
p=0.000819
5e+07
4e+07
3e+07
2e+07
1e+07
sphingomyelin (d18:2/24:2)*
p=0.134
p=0.000199
4e+07
3e+07
2e+07
1e+07
2−hydroxy−3−methylvalerate
p=0.658
p=6.8e−07
1e+07
5e+06
0e+00
Metabolite peak area abundance
baseline
endpoint (12 months)
Time point

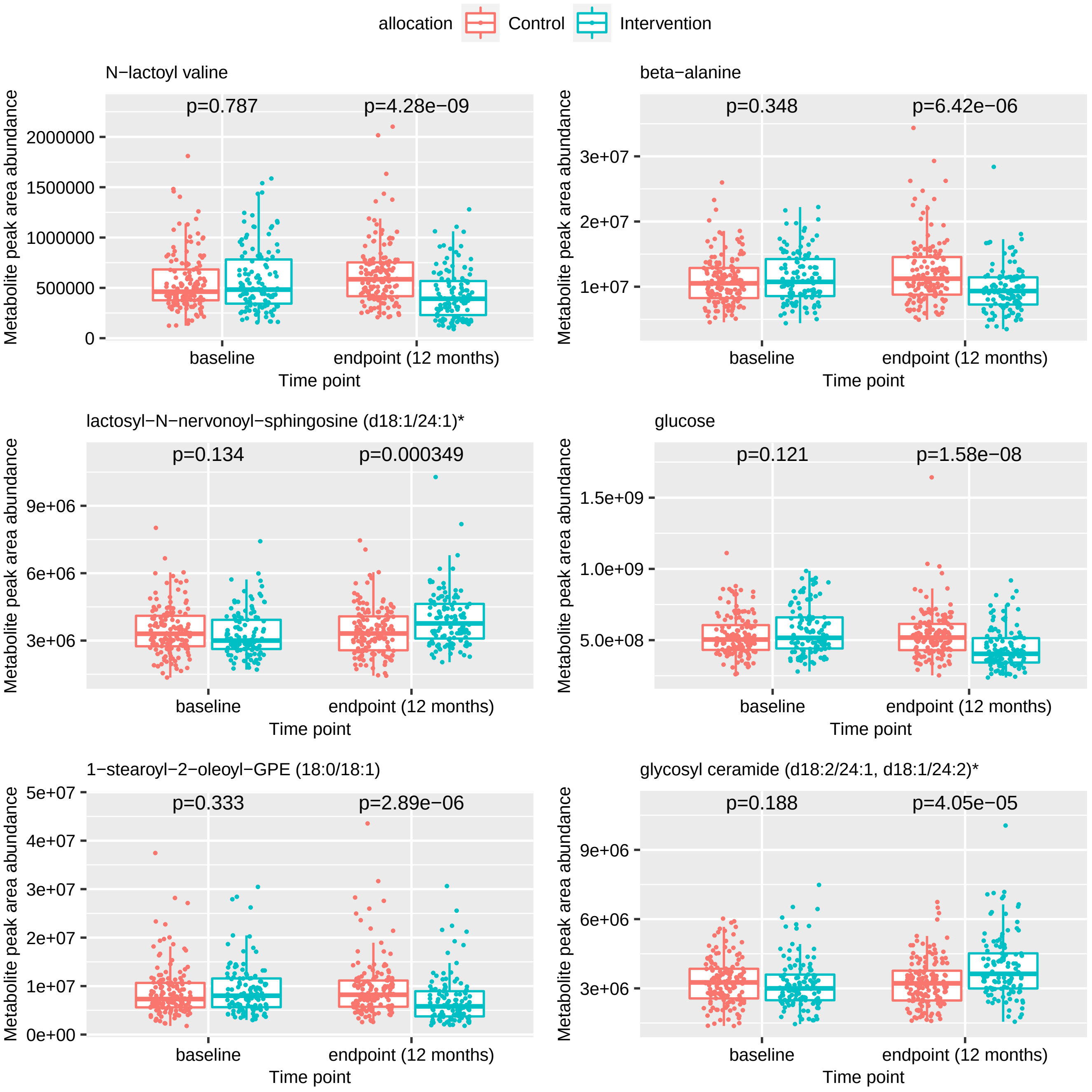

allocation
Control
Intervention
N−lactoyl valine
p=0.787
p=4.28e−09
beta−alanine
p=0.348
p=6.42e−06
lactosyl−N−nervonoyl−sphingosine (d18:1/24:1)*
p=0.134
p=0.000349
glucose
p=0.121
p=1.58e−08
1−stearoyl−2−oleoyl−GPE (18:0/18:1)
p=0.333
p=2.89e−06
glycosyl ceramide (d18:2/24:1, d18:1/24:2)*
p=0.188
p=4.05e−05
Metabolite peak area abundance
baseline
endpoint (12 months)
Time point

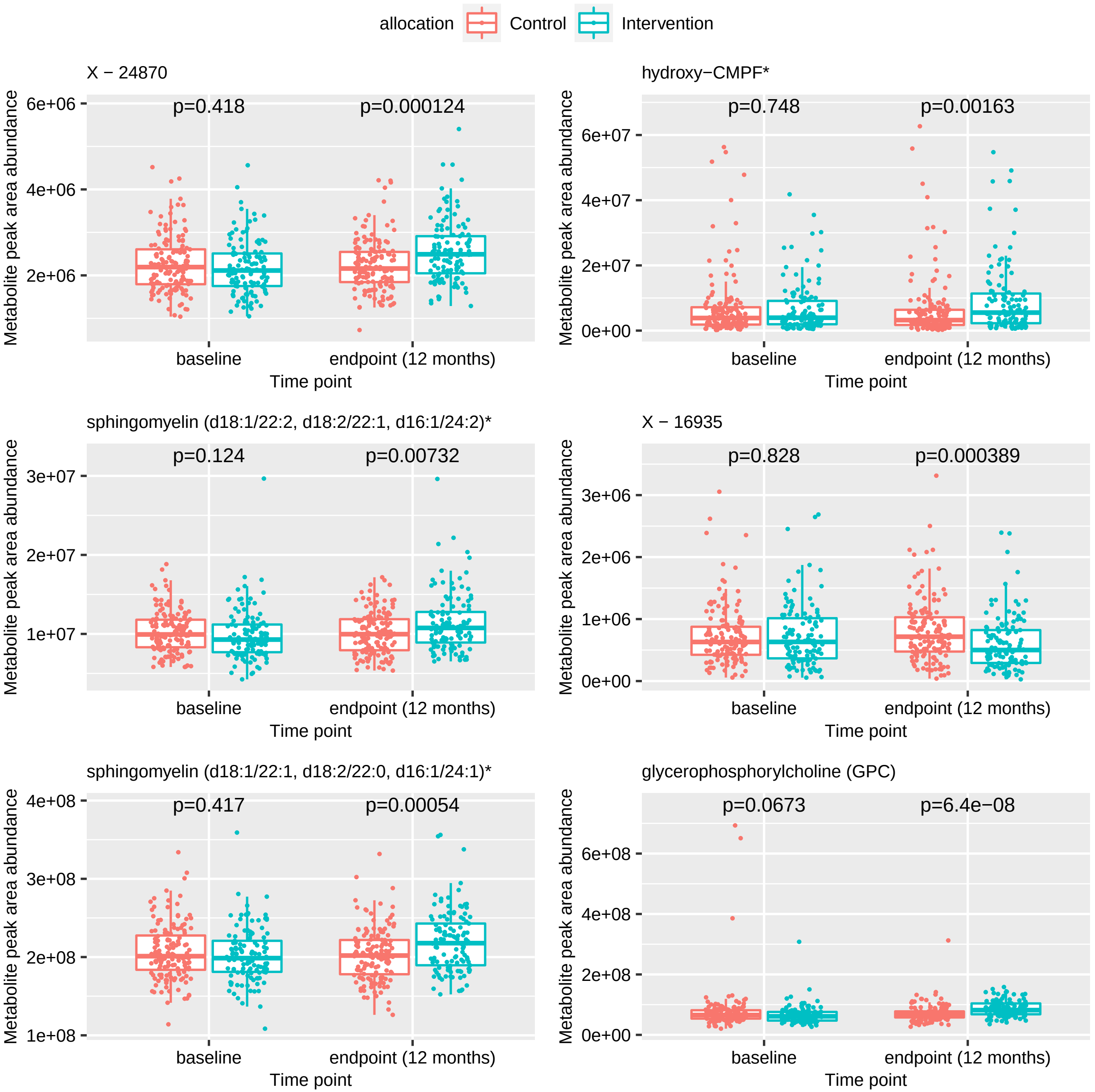
allocation
Control
Intervention
X − 24870
p=0.418
p=0.000124
hydroxy−CMPF*
p=0.748
p=0.00163
sphingomyelin (d18:1/22:2, d18:2/22:1, d16:1/24:2)*
p=0.124
p=0.00732
X − 16935
p=0.828
p=0.000389
sphingomyelin (d18:1/22:1, d18:2/22:0, d16:1/24:1)*
p=0.417
p=0.00054
glycerophosphorylcholine (GPC)
p=0.0673
p=6.4e−08
Metabolite peak area abundance
Time point
baseline
endpoint (12 months)
6e+06
4e+06
2e+06
6e+07
4e+07
2e+07
0e+00
3e+07
2e+07
1e+07
3e+06
2e+06
1e+06
4e+08
3e+08
2e+08
1e+08
6e+08
4e+08
2e+08

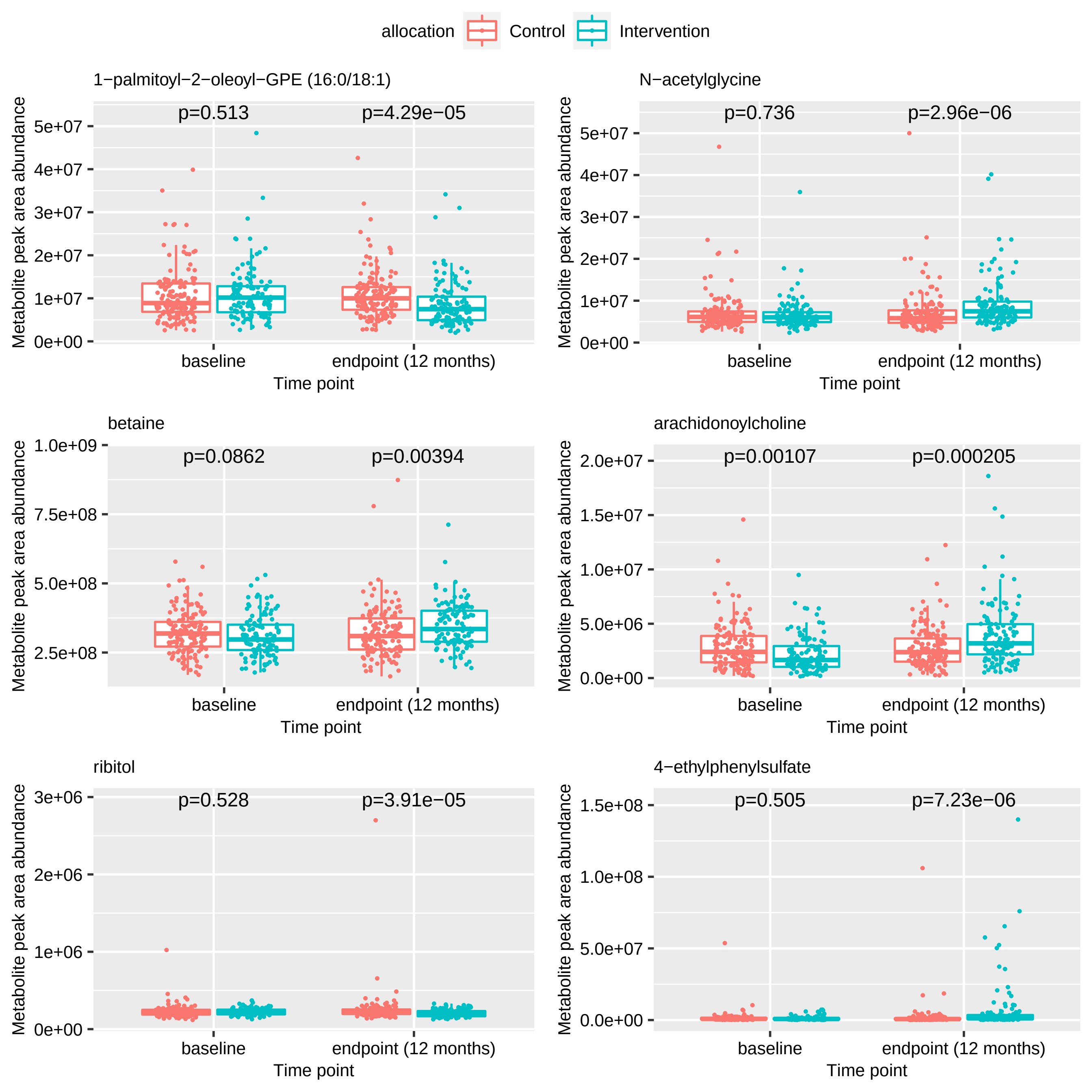

allocation
Control
Intervention
1−palmitoyl−2−oleoyl−GPE (16:0/18:1)
p=0.513
p=4.29e−05
N−acetylglycine
p=0.736
p=2.96e−06
betaine
p=0.0862
p=0.00394
arachidonoylcholine
p=0.00107
p=0.000205
ribitol
p=0.528
p=3.91e−05
4−ethylphenylsulfate
p=0.505
p=7.23e−06
Metabolite peak area abundance
Time point
baseline
endpoint (12 months)

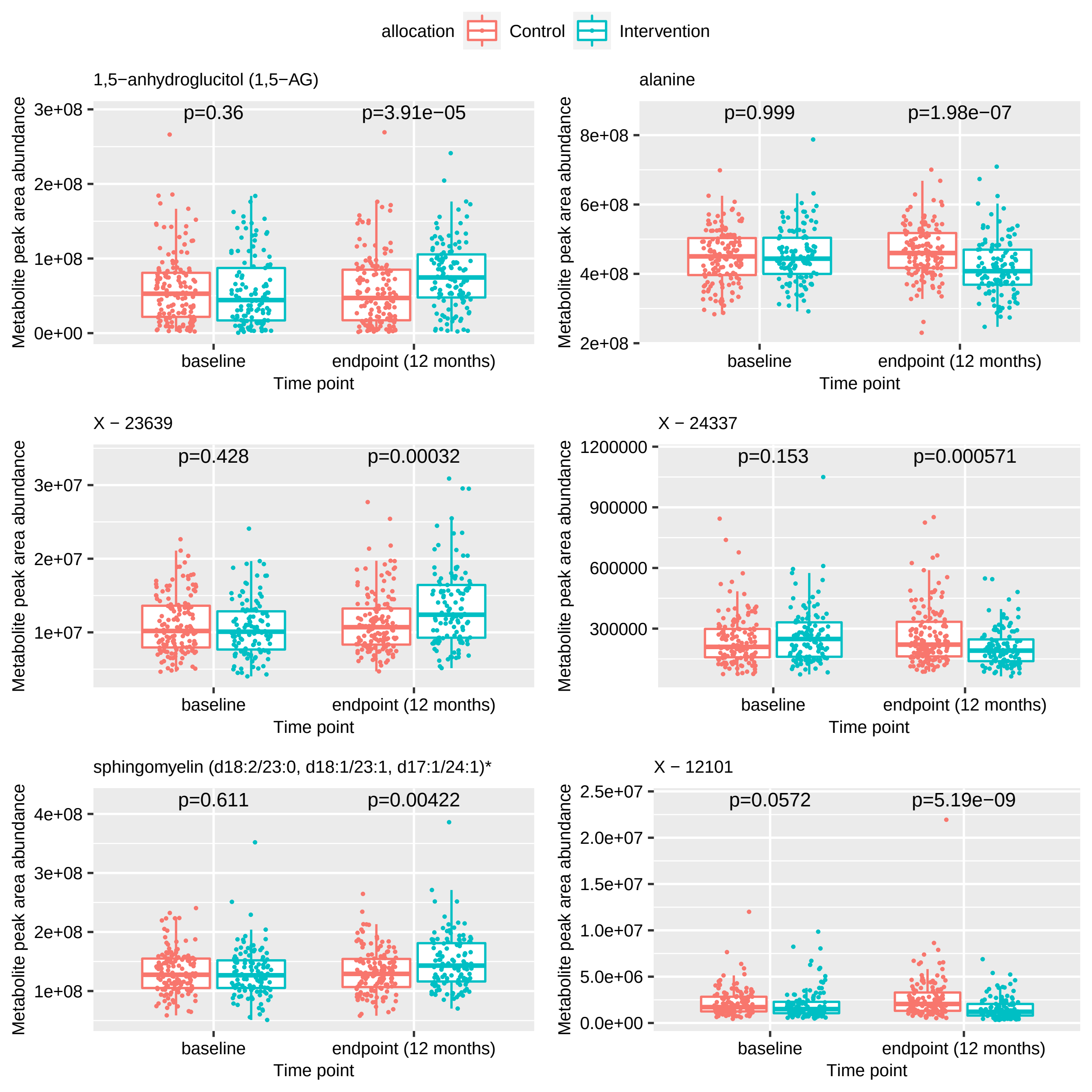
allocation
Control
Intervention
1,5−anhydroglucitol (1,5−AG)
p=0.36
p=3.91e−05
alanine
p=0.999
p=1.98e−07
X − 23639
p=0.428
p=0.00032
X − 24337
p=0.153
p=0.000571
sphingomyelin (d18:2/23:0, d18:1/23:1, d17:1/24:1)*
p=0.611
p=0.00422
X − 12101
p=0.0572
p=5.19e−09
Metabolite peak area abundance
Time point
baseline
endpoint (12 months)

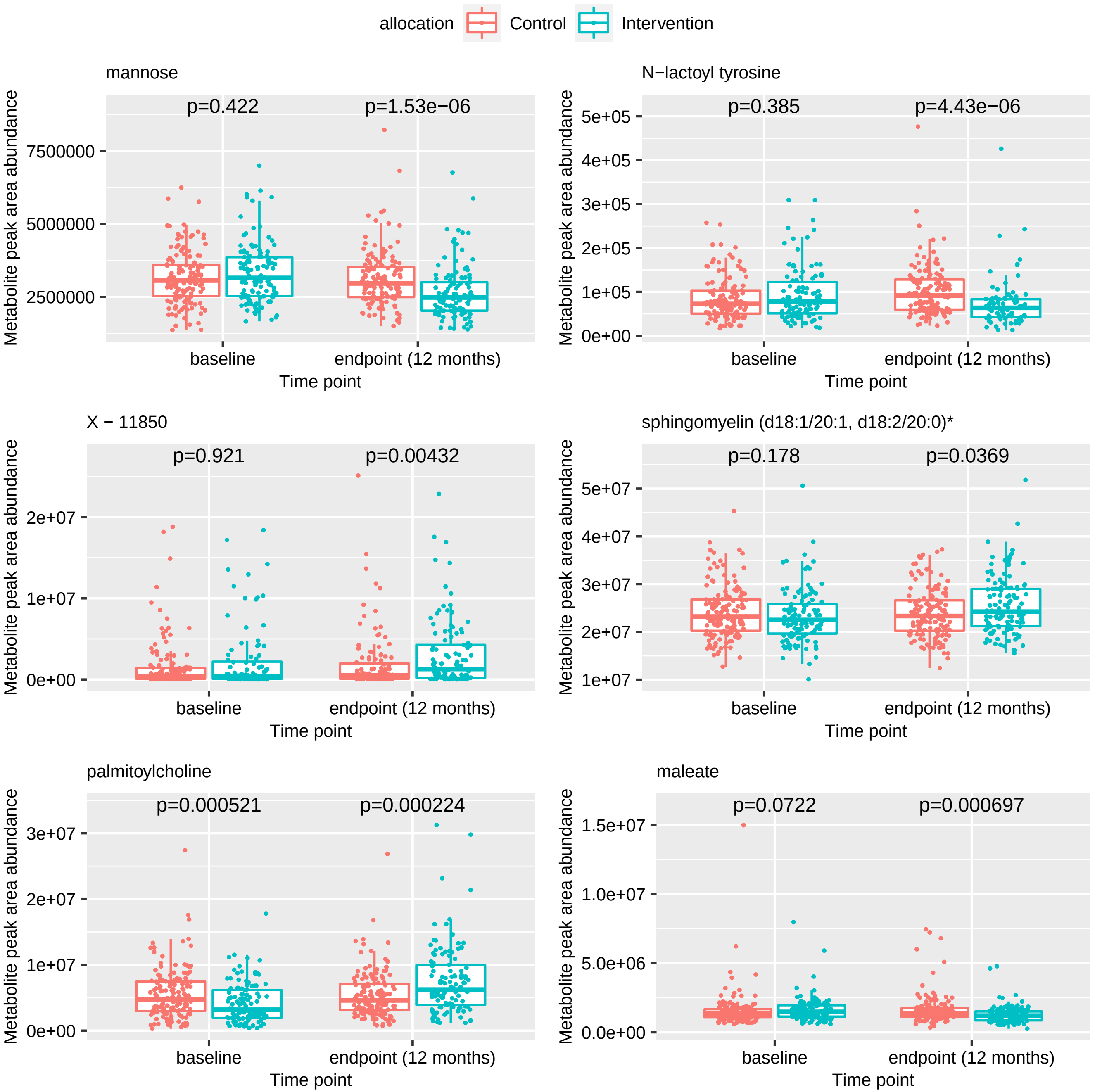
allocation
Control
Intervention
mannose
p=0.422
p=1.53e−06
7500000
5000000
2500000
N−lactoyl tyrosine
p=0.385
p=4.43e−06
5e+05
4e+05
3e+05
2e+05
1e+05
0e+00
X − 11850
p=0.921
p=0.00432
2e+07
1e+07
0e+00
sphingomyelin (d18:1/20:1, d18:2/20:0)*
p=0.178
p=0.0369
5e+07
4e+07
3e+07
2e+07
1e+07
palmitoylcholine
p=0.000521
p=0.000224
3e+07
2e+07
1e+07
0e+00
maleate
p=0.0722
p=0.000697
1.5e+07
1.0e+07
5.0e+06
0.0e+00
Metabolite peak area abundance
baseline
endpoint (12 months)
Time point

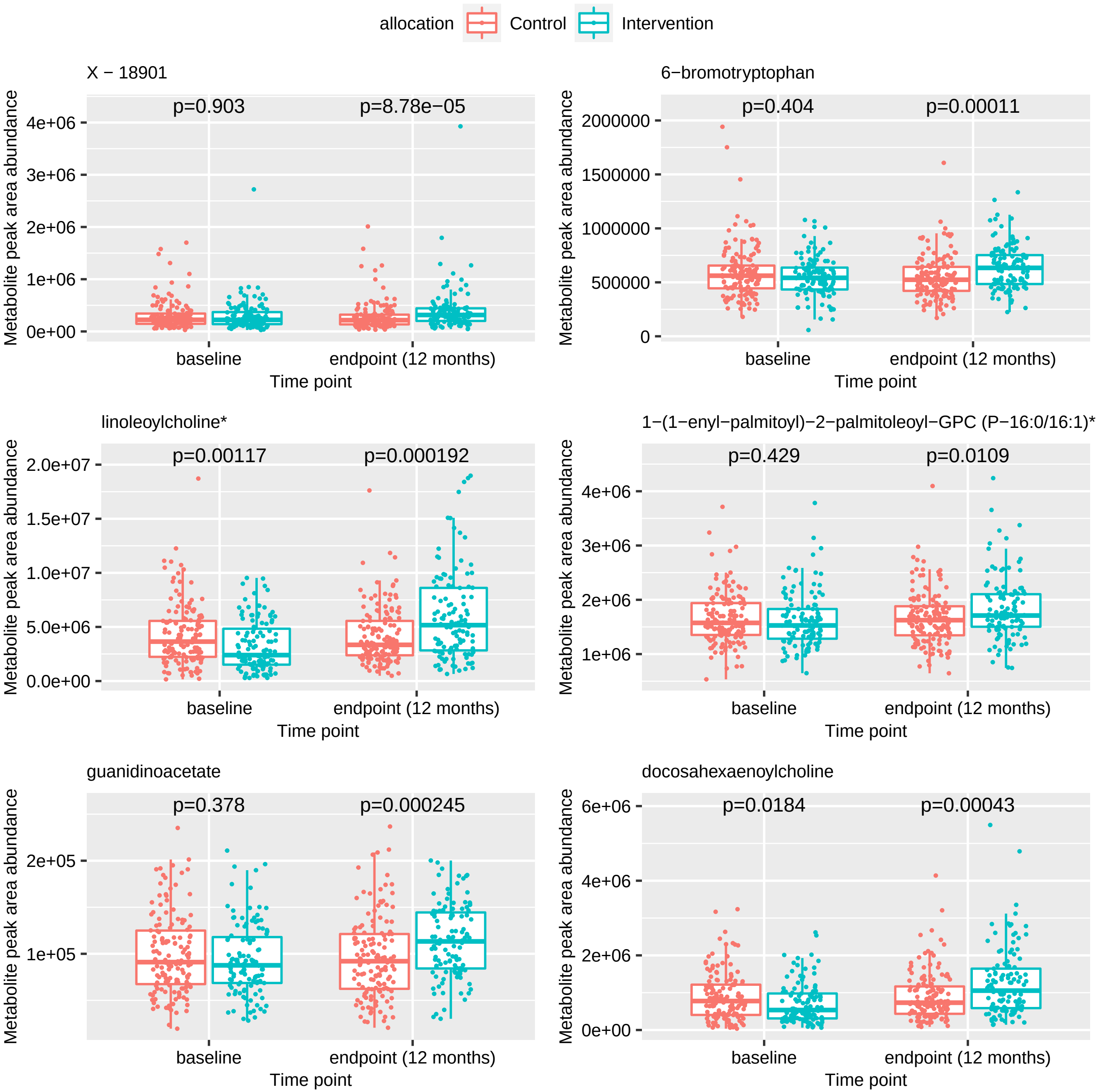
allocation
Control
Intervention
X − 18901
p=0.903
p=8.78e−05
6−bromotryptophan
p=0.404
p=0.00011
linoleoylcholine*
p=0.00117
p=0.000192
1−(1−enyl−palmitoyl)−2−palmitoleoyl−GPC (P−16:0/16:1)*
p=0.429
p=0.0109
guanidinoacetate
p=0.378
p=0.000245
docosahexaenoylcholine
p=0.0184
p=0.00043
Metabolite peak area abundance
baseline
endpoint (12 months)
Time point

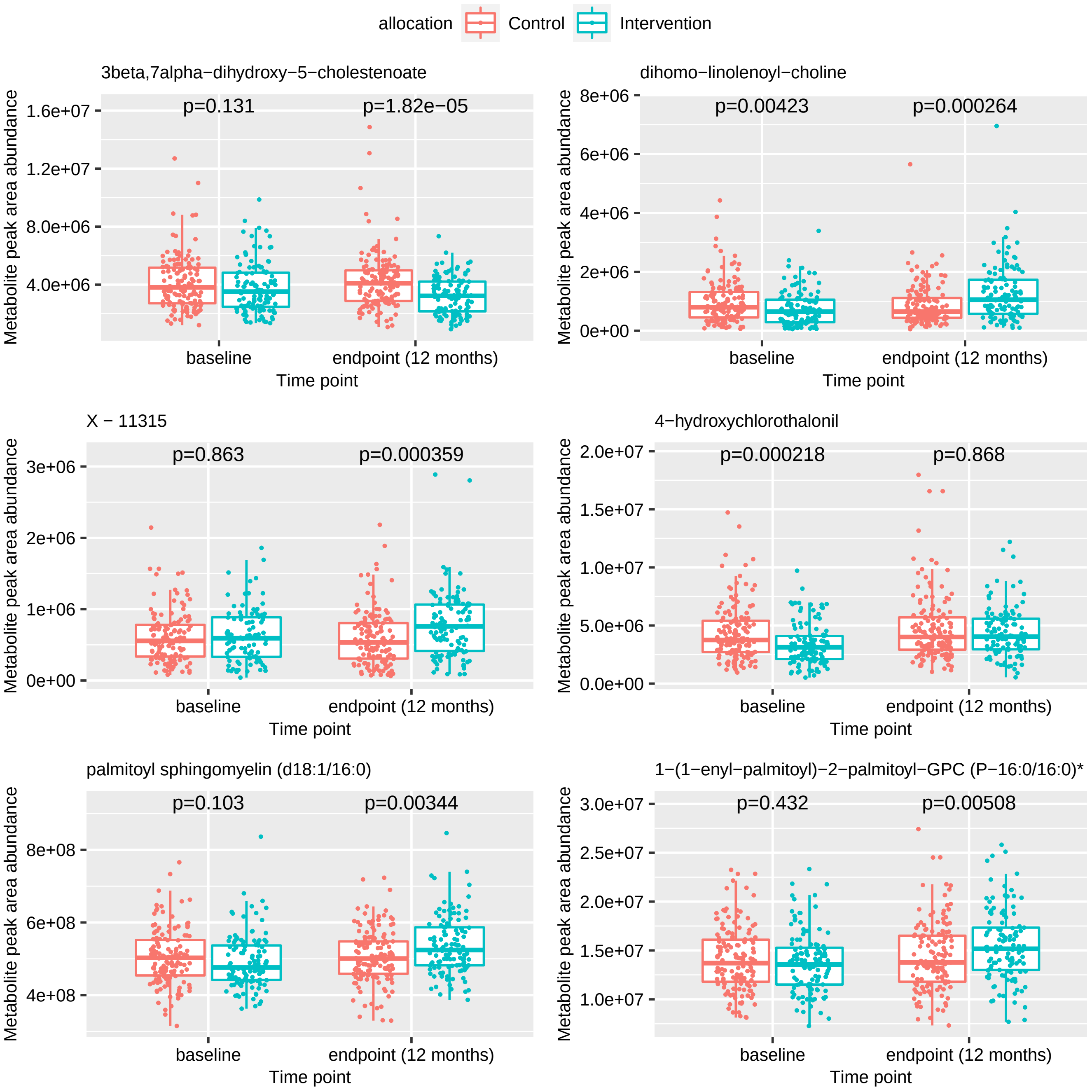

allocation
Control
Intervention
3beta,7alpha−dihydroxy−5−cholestenoate
p=0.131
p=1.82e−05
dihomo−linolenoyl−choline
p=0.00423
p=0.000264
X − 11315
p=0.863
p=0.000359
4−hydroxychlorothalonil
p=0.000218
p=0.868
palmitoyl sphingomyelin (d18:1/16:0)
p=0.103
p=0.00344
1−(1−enyl−palmitoyl)−2−palmitoyl−GPC (P−16:0/16:0)*
p=0.432
p=0.00508
Metabolite peak area abundance
baseline
endpoint (12 months)
Time point

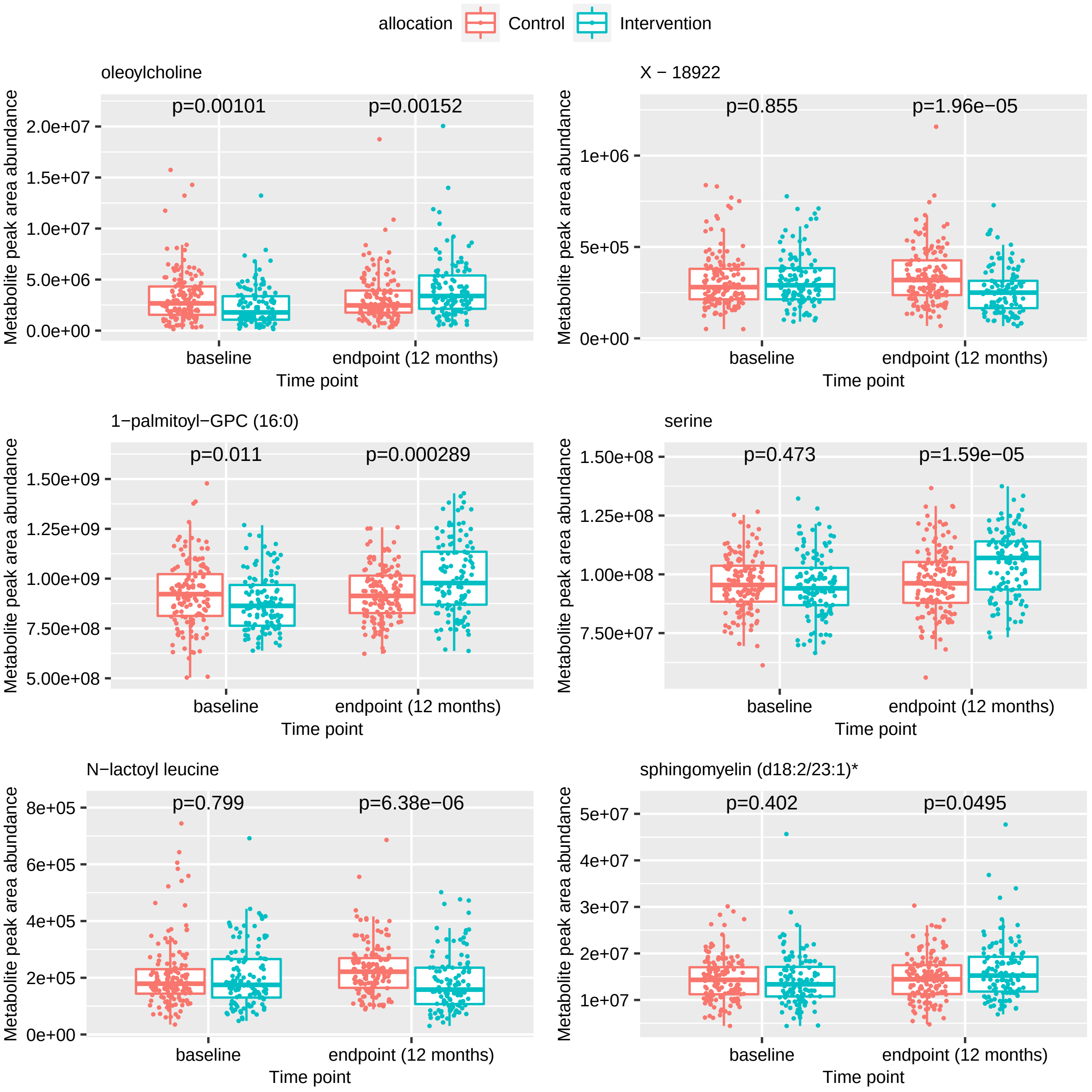
allocation
Control
Intervention
oleoylcholine
p=0.00101
p=0.00152
Metabolite peak area abundance
2.0e+07
1.5e+07
1.0e+07
5.0e+06
0.0e+00
baseline
endpoint (12 months)
Time point
X − 18922
p=0.855
p=1.96e−05
1e+06
5e+05
0e+00
1−palmitoyl−GPC (16:0)
p=0.011
p=0.000289
1.50e+09
1.25e+09
1.00e+09
7.50e+08
5.00e+08
serine
p=0.473
p=1.59e−05
1.50e+08
1.25e+08
1.00e+08
7.50e+07
N−lactoyl leucine
p=0.799
p=6.38e−06
8e+05
6e+05
4e+05
2e+05
0e+00
sphingomyelin (d18:2/23:1)*
p=0.402
p=0.0495
5e+07
4e+07
3e+07
2e+07
1e+07

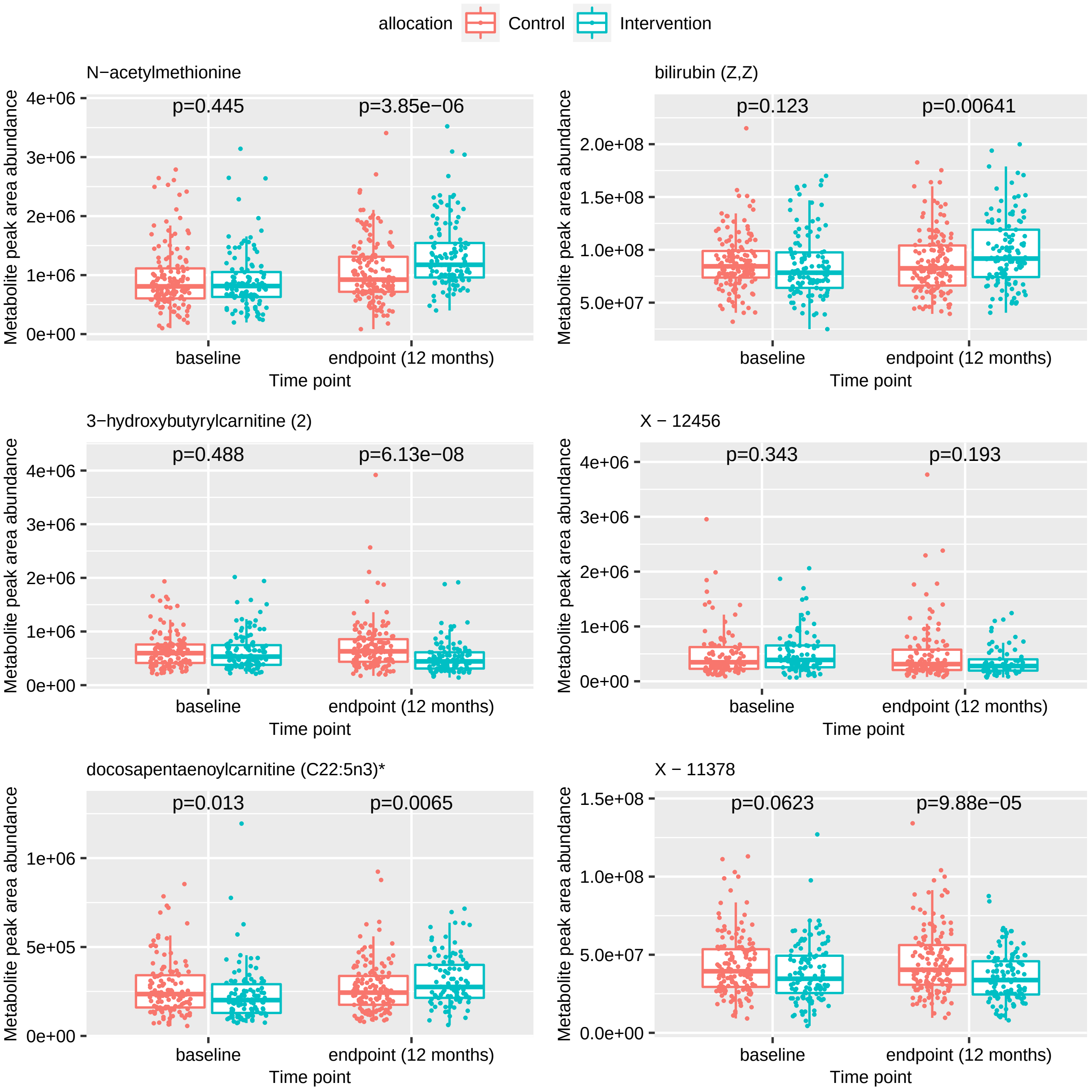
allocation
Control
Intervention
N−acetylmethionine
p=0.445
p=3.85e−06
bilirubin (Z,Z)
p=0.123
p=0.00641
3−hydroxybutyrylcarnitine (2)
p=0.488
p=6.13e−08
X − 12456
p=0.343
p=0.193
docosapentaenoylcarnitine (C22:5n3)*
p=0.013
p=0.0065
X − 11378
p=0.0623
p=9.88e−05
Metabolite peak area abundance
baseline
endpoint (12 months)
Time point

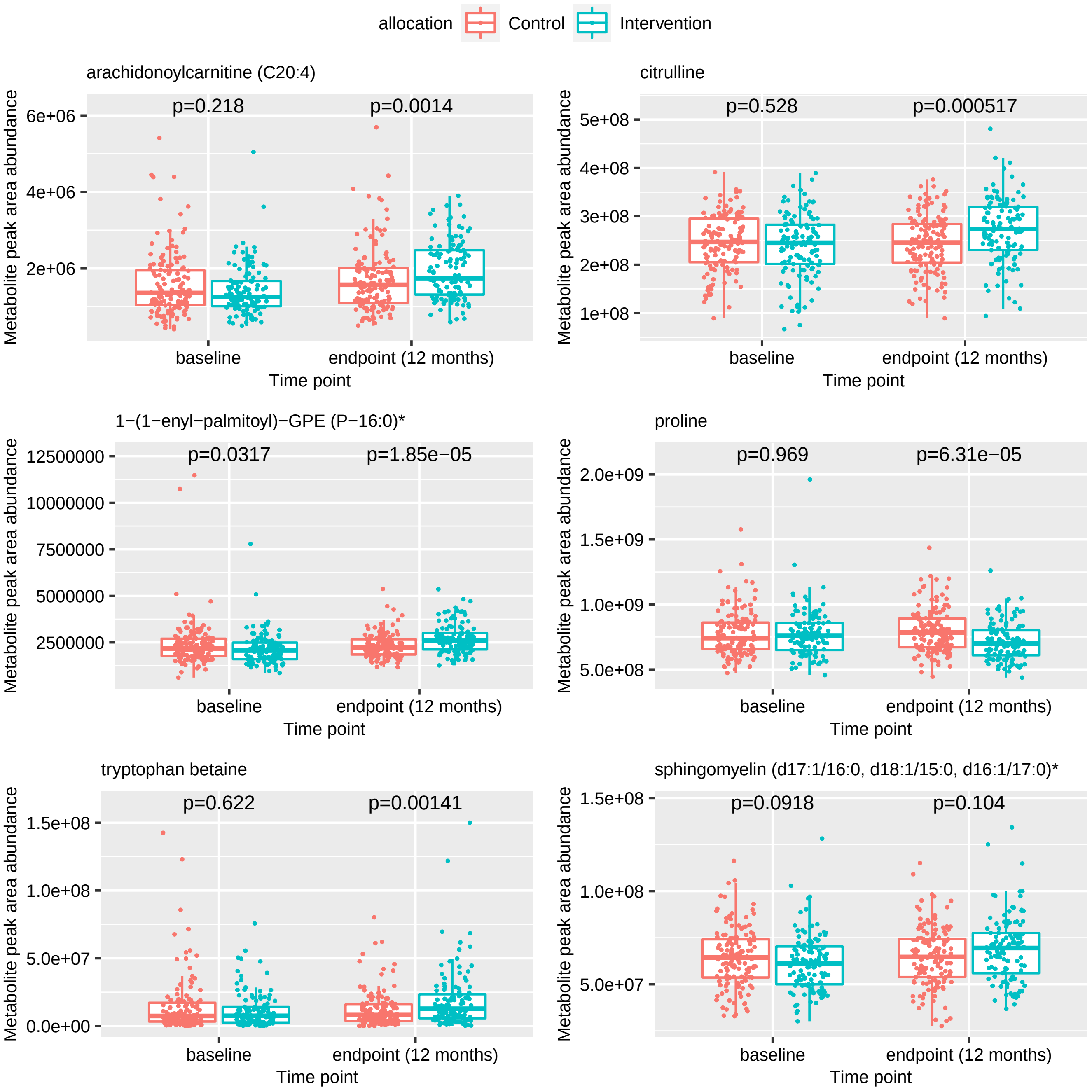

allocation
Control
Intervention
arachidonoylcarnitine (C20:4)
p=0.218
p=0.0014
citrulline
p=0.528
p=0.000517
1−(1−enyl−palmitoyl)−GPE (P−16:0)*
p=0.0317
p=1.85e−05
proline
p=0.969
p=6.31e−05
tryptophan betaine
p=0.622
p=0.00141
sphingomyelin (d17:1/16:0, d18:1/15:0, d16:1/17:0)*
p=0.0918
p=0.104
Metabolite peak area abundance
baseline
endpoint (12 months)
Time point

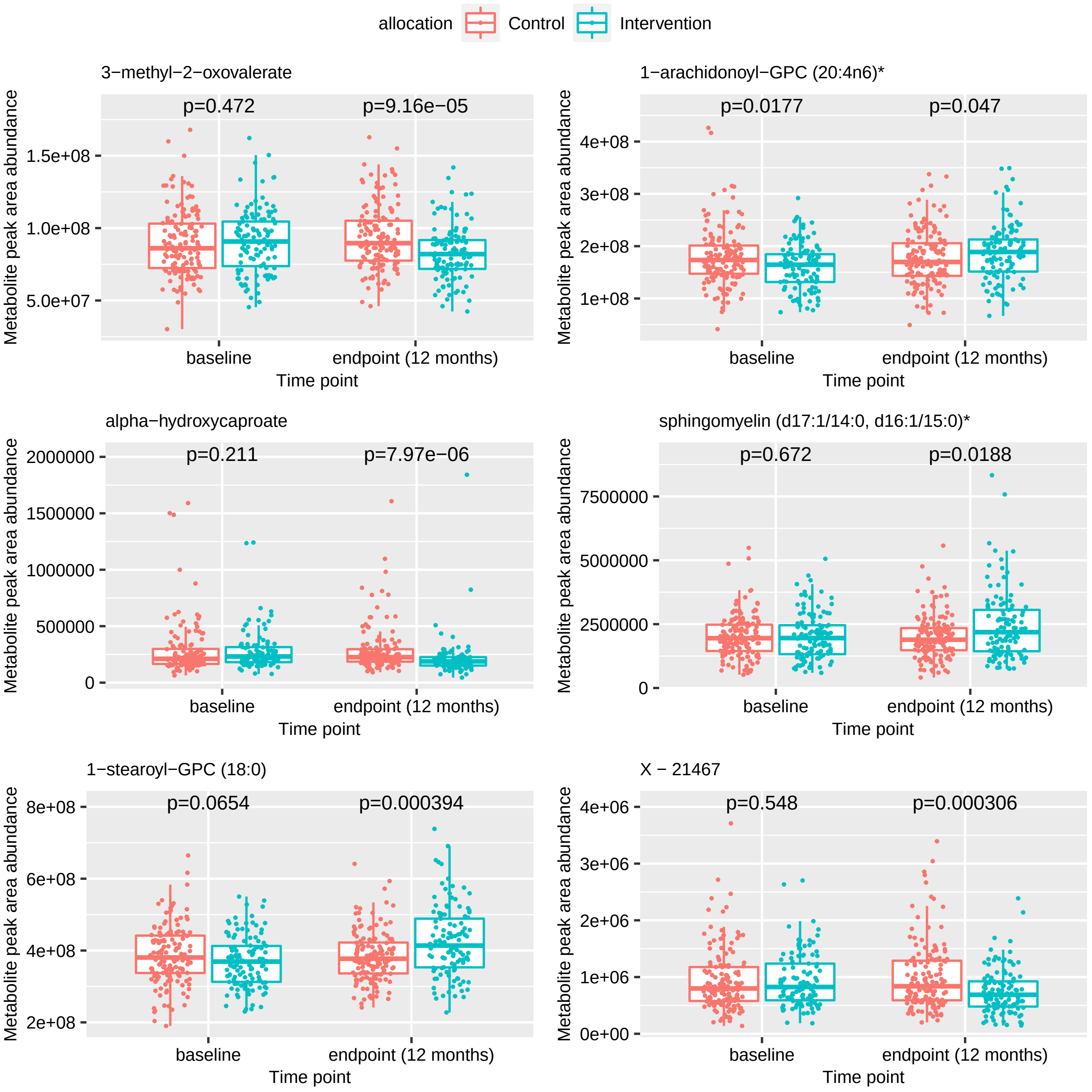

allocation
Control
Intervention
3−methyl−2−oxovalerate
p=0.472
p=9.16e−05
1−arachidonoyl−GPC (20:4n6)*
p=0.0177
p=0.047
alpha−hydroxycaproate
p=0.211
p=7.97e−06
sphingomyelin (d17:1/14:0, d16:1/15:0)*
p=0.672
p=0.0188
1−stearoyl−GPC (18:0)
p=0.0654
p=0.000394
X − 21467
p=0.548
p=0.000306
Metabolite peak area abundance
baseline
endpoint (12 months)
Time point

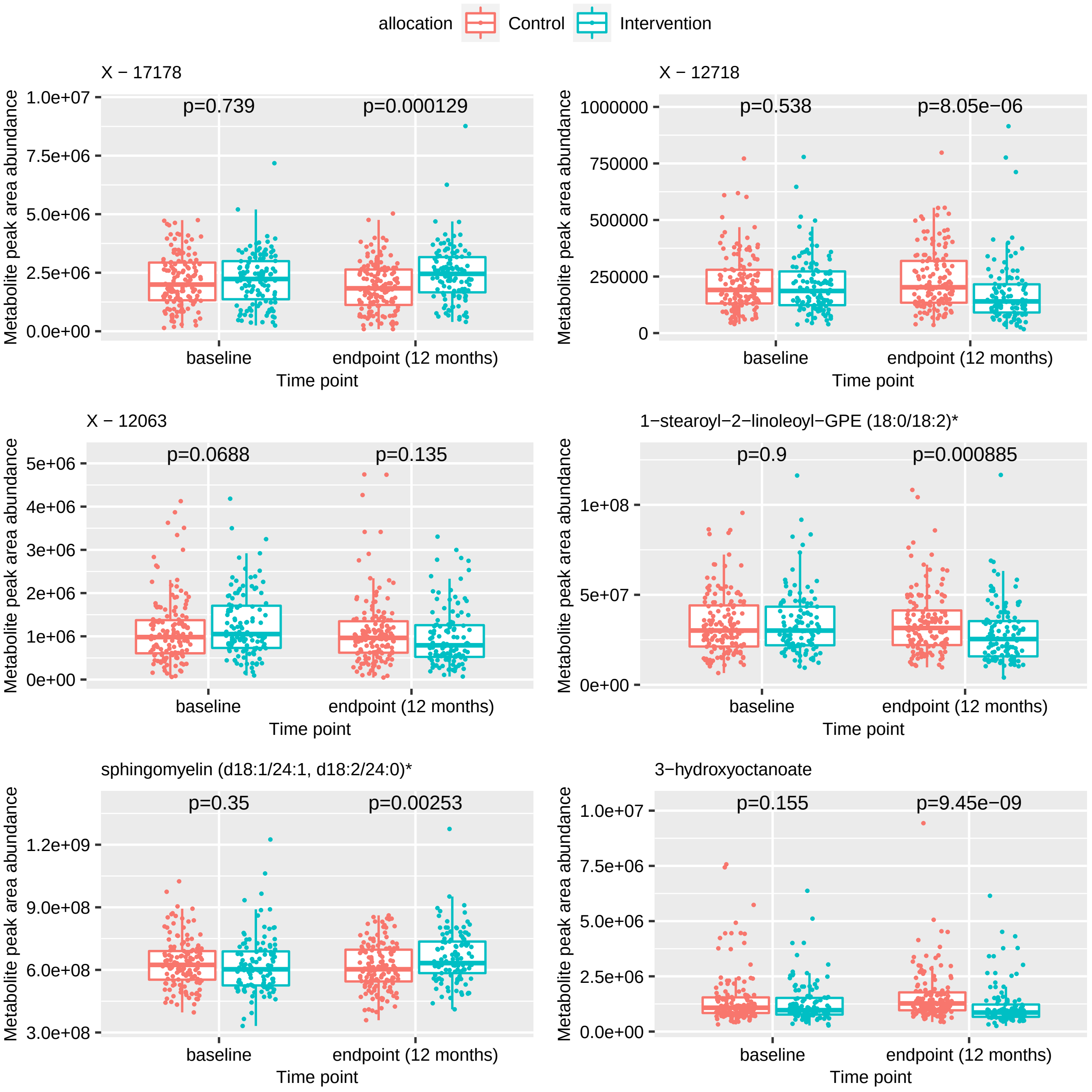

allocation
Control
Intervention
X − 17178
p=0.739
p=0.000129
X − 12718
p=0.538
p=8.05e−06
X − 12063
p=0.0688
p=0.135
1−stearoyl−2−linoleoyl−GPE (18:0/18:2)*
p=0.9
p=0.000885
sphingomyelin (d18:1/24:1, d18:2/24:0)*
p=0.35
p=0.00253
3−hydroxyoctanoate
p=0.155
p=9.45e−09
Metabolite peak area abundance
Time point
baseline
endpoint (12 months)

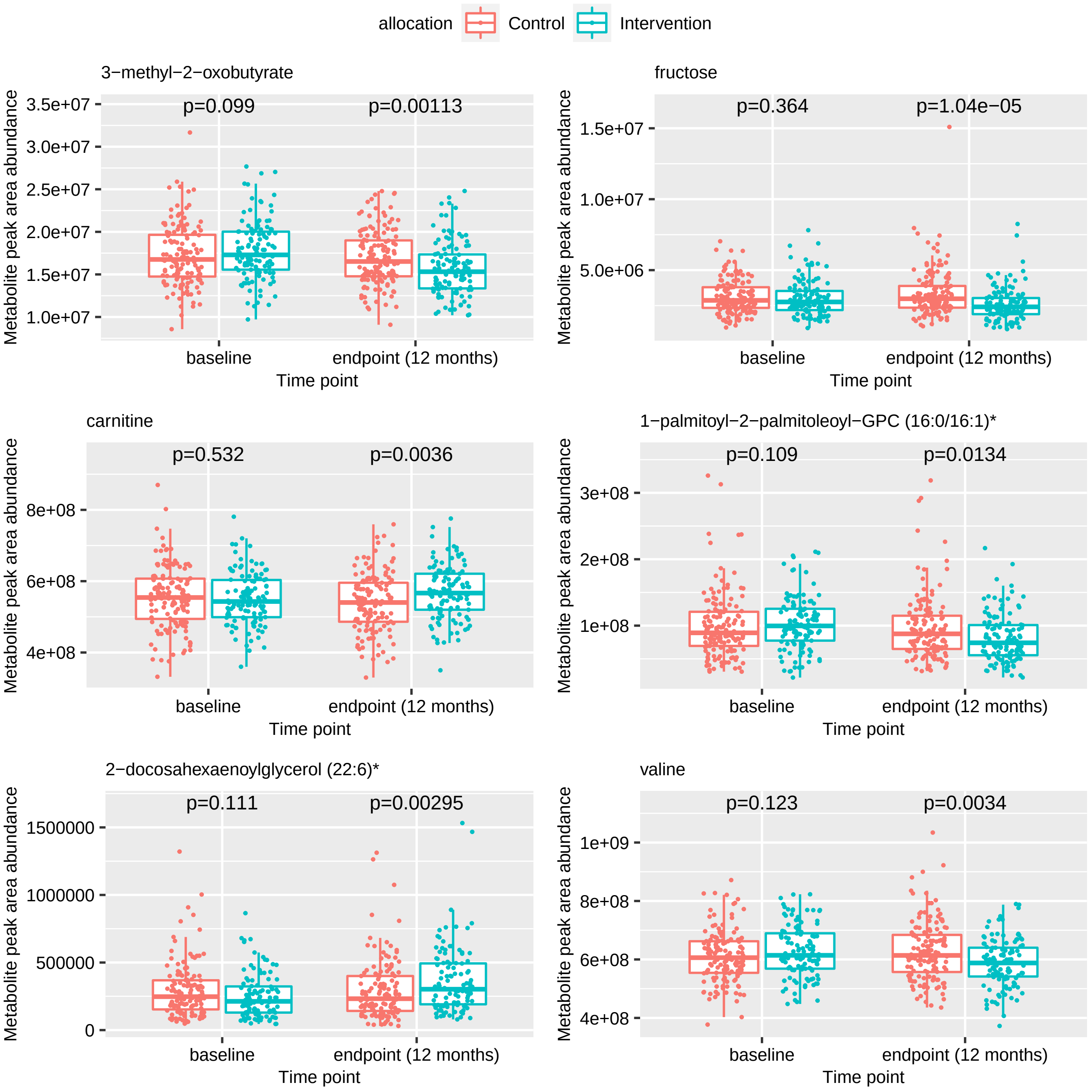
allocation
Control
Intervention
3−methyl−2−oxobutyrate
p=0.099
p=0.00113
fructose
p=0.364
p=1.04e−05
carnitine
p=0.532
p=0.0036
1−palmitoyl−2−palmitoleoyl−GPC (16:0/16:1)*
p=0.109
p=0.0134
2−docosahexaenoylglycerol (22:6)*
p=0.111
p=0.00295
valine
p=0.123
p=0.0034
Metabolite peak area abundance
Time point
baseline
endpoint (12 months)

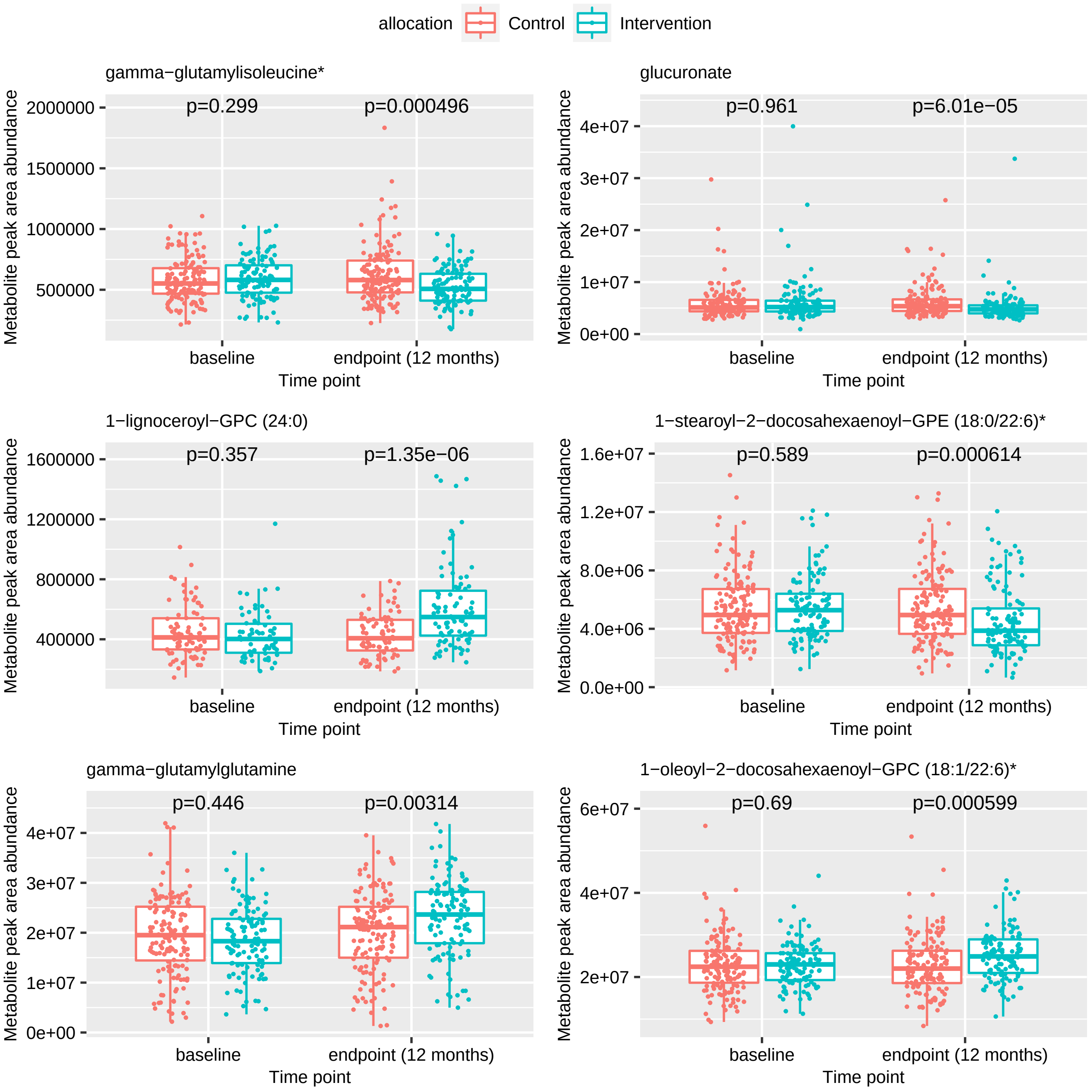

allocation
Control
Intervention
gamma−glutamylisoleucine*
p=0.299
p=0.000496
glucuronate
p=0.961
p=6.01e−05
1−lignoceroyl−GPC (24:0)
p=0.357
p=1.35e−06
1−stearoyl−2−docosahexaenoyl−GPE (18:0/22:6)*
p=0.589
p=0.000614
gamma−glutamylglutamine
p=0.446
p=0.00314
1−oleoyl−2−docosahexaenoyl−GPC (18:1/22:6)*
p=0.69
p=0.000599
Metabolite peak area abundance
baseline
endpoint (12 months)
Time point

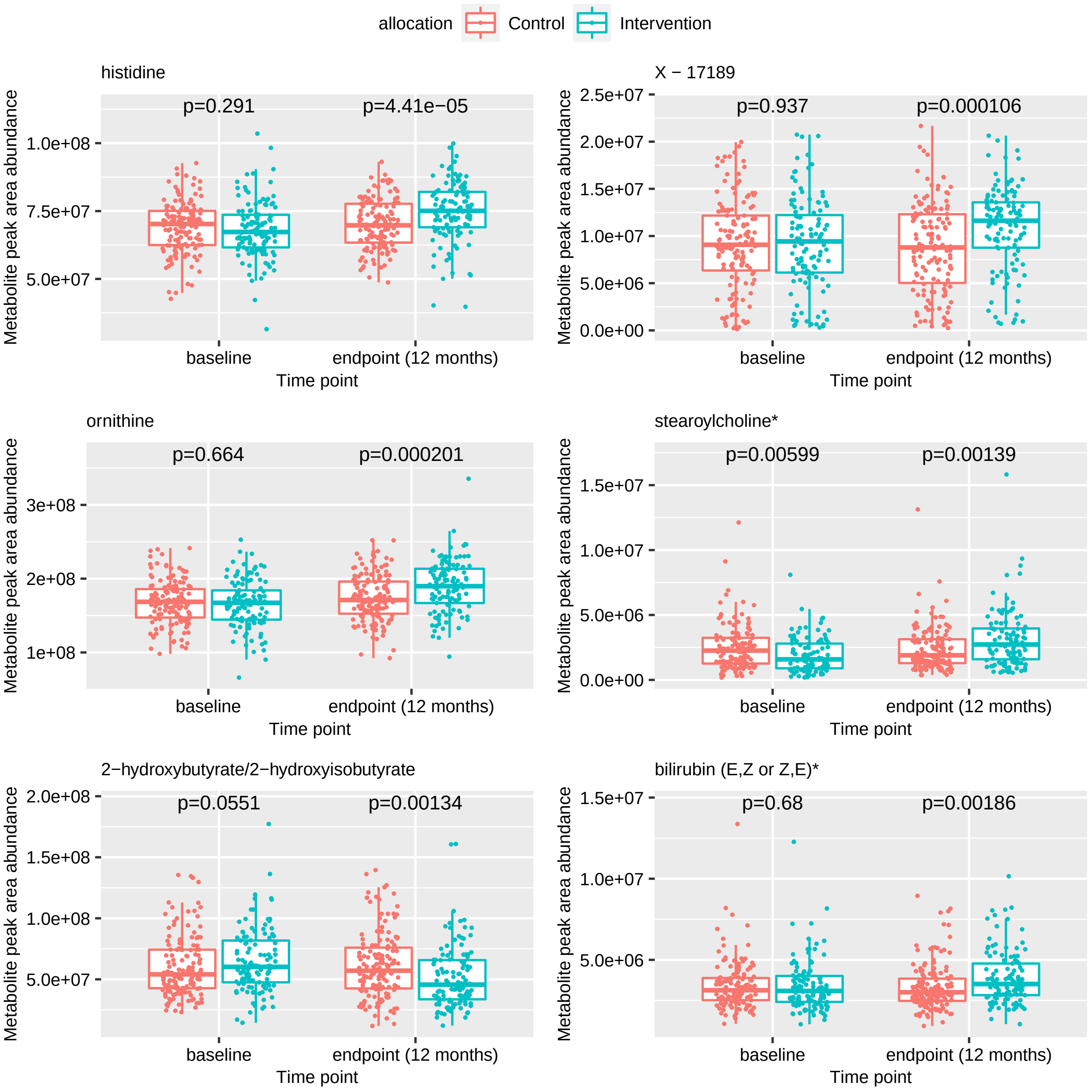

allocation
Control
Intervention
histidine
p=0.291
p=4.41e−05
1.0e+08
7.5e+07
5.0e+07
Metabolite peak area abundance
baseline
endpoint (12 months)
Time point
X − 17189
p=0.937
p=0.000106
2.5e+07
2.0e+07
1.5e+07
1.0e+07
5.0e+06
0.0e+00
ornithine
p=0.664
p=0.000201
3e+08
2e+08
1e+08
stearoylcholine*
p=0.00599
p=0.00139
1.5e+07
1.0e+07
5.0e+06
0.0e+00
2−hydroxybutyrate/2−hydroxyisobutyrate
p=0.0551
p=0.00134
2.0e+08
1.5e+08
1.0e+08
5.0e+07
bilirubin (E,Z or Z,E)*
p=0.68
p=0.00186
1.5e+07
1.0e+07
5.0e+06

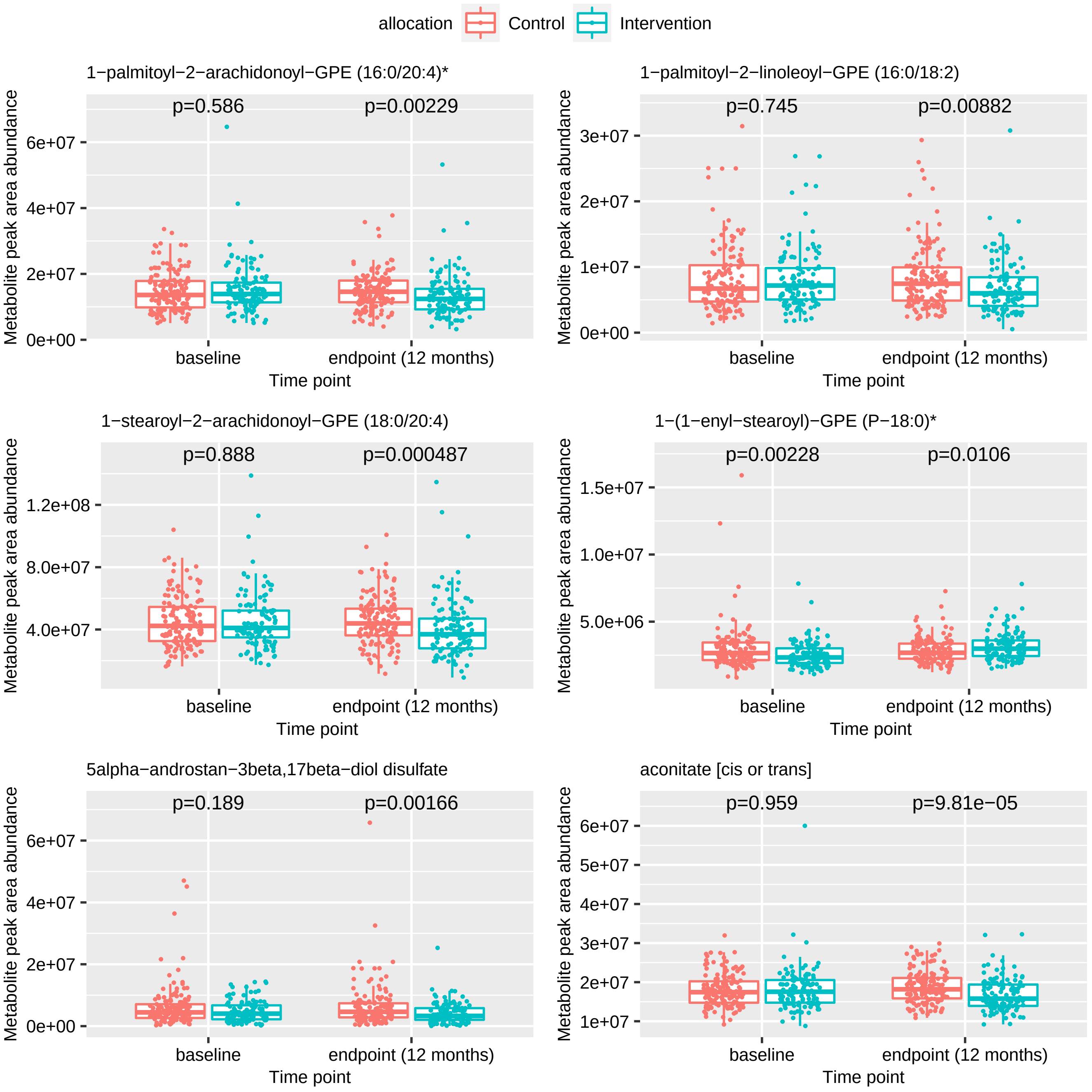
allocation
Control
Intervention
1−palmitoyl−2−arachidonoyl−GPE (16:0/20:4)*
p=0.586
p=0.00229
1−palmitoyl−2−linoleoyl−GPE (16:0/18:2)
p=0.745
p=0.00882
1−stearoyl−2−arachidonoyl−GPE (18:0/20:4)
p=0.888
p=0.000487
1−(1−enyl−stearoyl)−GPE (P−18:0)*
p=0.00228
p=0.0106
5alpha−androstan−3beta,17beta−diol disulfate
p=0.189
p=0.00166
aconitate [cis or trans]
p=0.959
p=9.81e−05
Metabolite peak area abundance
baseline
endpoint (12 months)
Time point

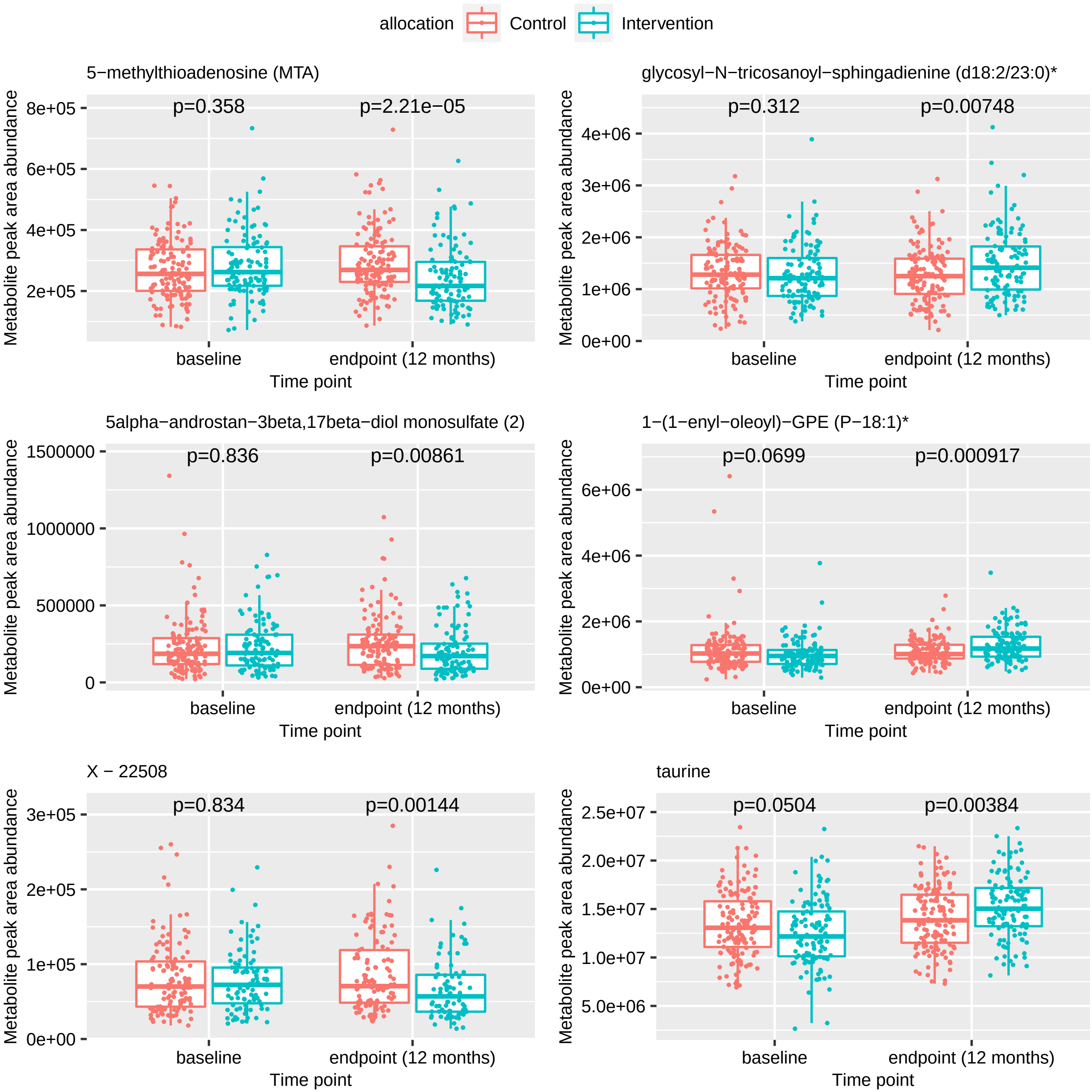
allocation
Control
Intervention
5−methylthioadenosine (MTA)
p=0.358
p=2.21e−05
glycosyl−N−tricosanoyl−sphingadienine (d18:2/23:0)*
p=0.312
p=0.00748
5alpha−androstan−3beta,17beta−diol monosulfate (2)
p=0.836
p=0.00861
1−(1−enyl−oleoyl)−GPE (P−18:1)*
p=0.0699
p=0.000917
X − 22508
p=0.834
p=0.00144
taurine
p=0.0504
p=0.00384
Metabolite peak area abundance
baseline
endpoint (12 months)
Time point

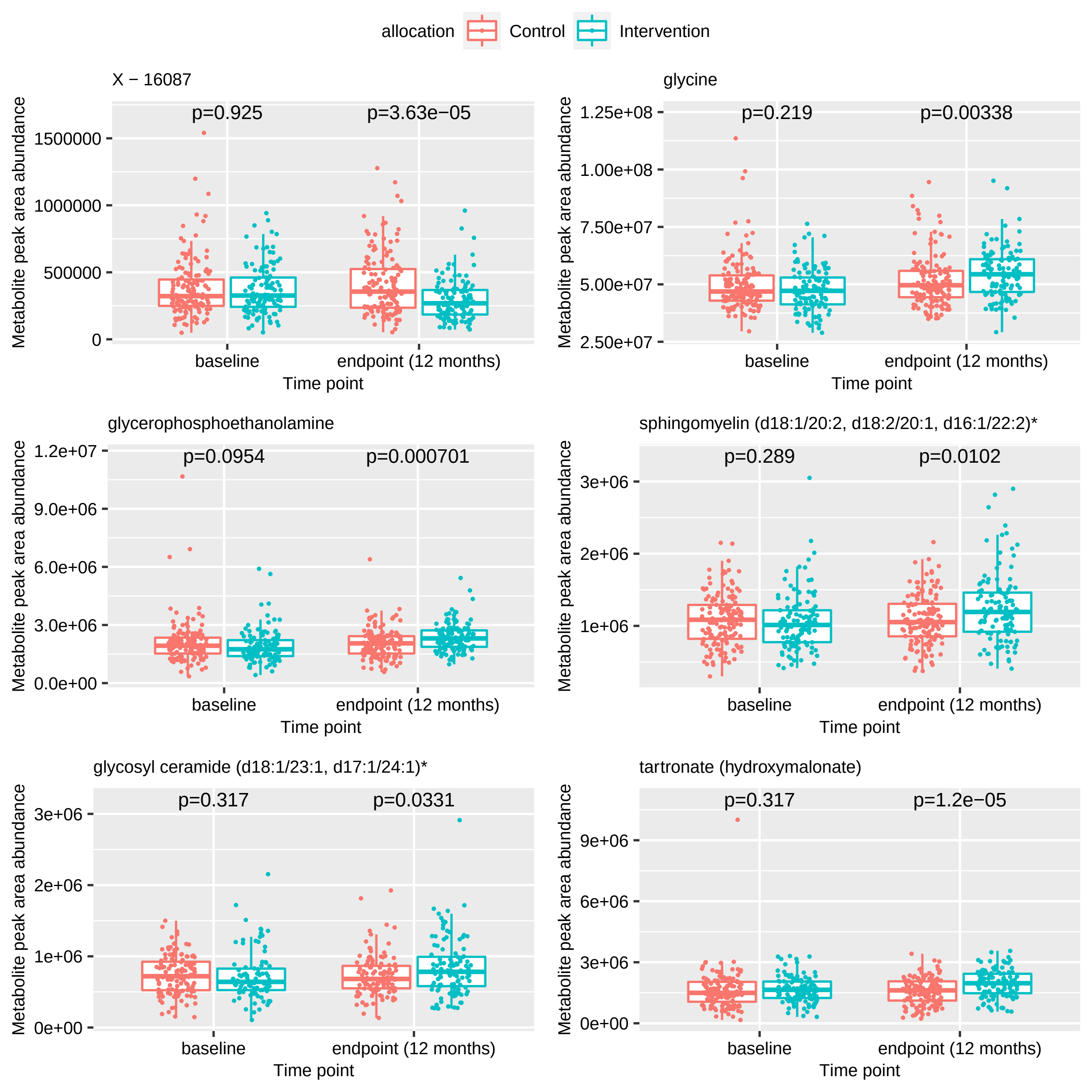

allocation
Control
Intervention
X − 16087
p=0.925
p=3.63e−05
glycine
p=0.219
p=0.00338
glycerophosphoethanolamine
p=0.0954
p=0.000701
sphingomyelin (d18:1/20:2, d18:2/20:1, d16:1/22:2)*
p=0.289
p=0.0102
glycosyl ceramide (d18:1/23:1, d17:1/24:1)*
p=0.317
p=0.0331
tartronate (hydroxymalonate)
p=0.317
p=1.2e−05
Metabolite peak area abundance
Time point
baseline
endpoint (12 months)
0
500000
1000000
1500000
2.50e+07
5.00e+07
7.50e+07
1.00e+08
1.25e+08
0.0e+00
3.0e+06
6.0e+06
9.0e+06
1.2e+07
1e+06
2e+06
3e+06
0e+00
1e+06
2e+06
3e+06
0e+00
3e+06
6e+06
9e+06

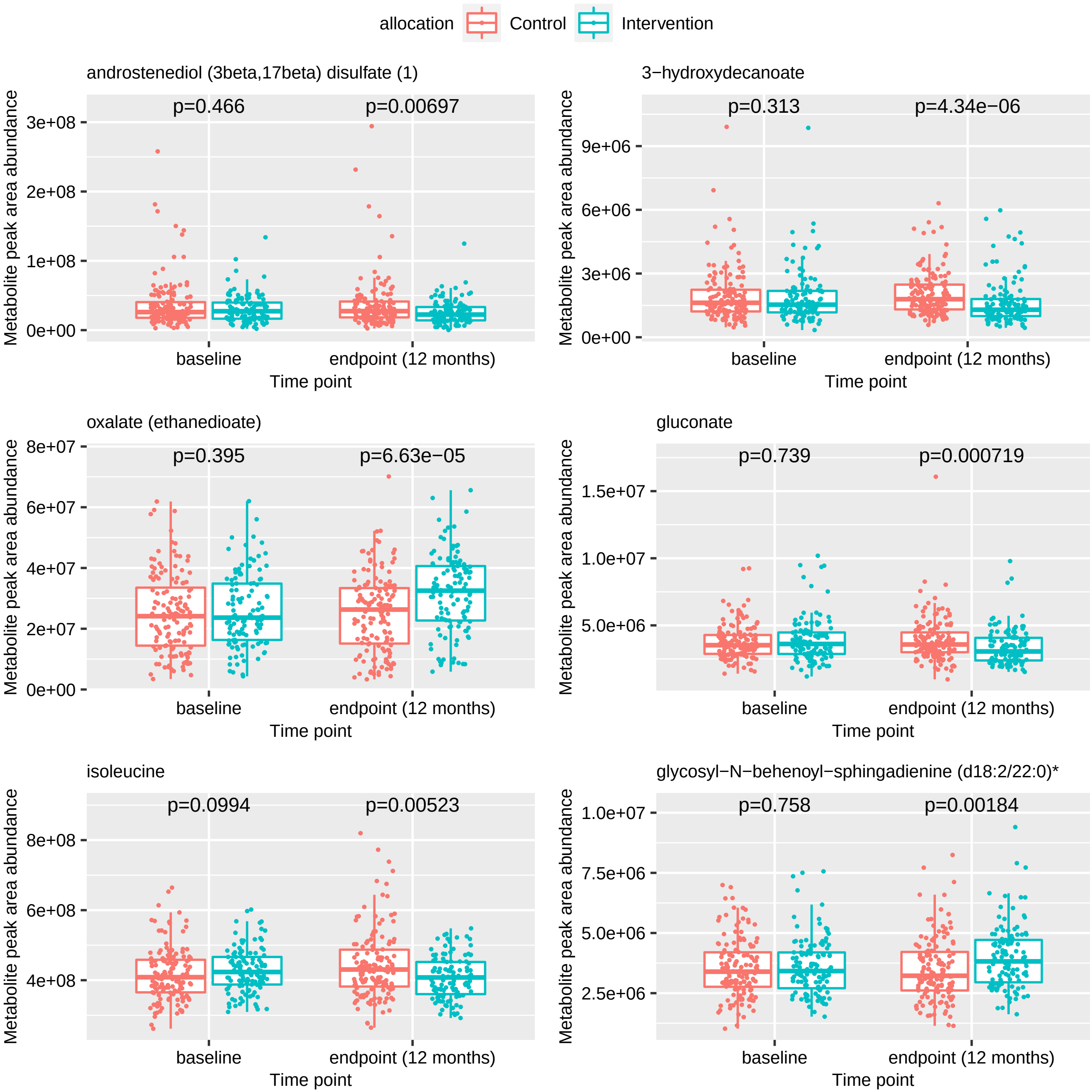

allocation
Control
Intervention
androstenediol (3beta,17beta) disulfate (1)
p=0.466
p=0.00697
3e+08
2e+08
1e+08
0e+00
3−hydroxydecanoate
p=0.313
p=4.34e−06
9e+06
6e+06
3e+06
0e+00
oxalate (ethanedioate)
p=0.395
p=6.63e−05
8e+07
6e+07
4e+07
2e+07
0e+00
gluconate
p=0.739
p=0.000719
1.5e+07
1.0e+07
5.0e+06
isoleucine
p=0.0994
p=0.00523
8e+08
6e+08
4e+08
glycosyl−N−behenoyl−sphingadienine (d18:2/22:0)*
p=0.758
p=0.00184
1.0e+07
7.5e+06
5.0e+06
2.5e+06
Metabolite peak area abundance
baseline
endpoint (12 months)
Time point

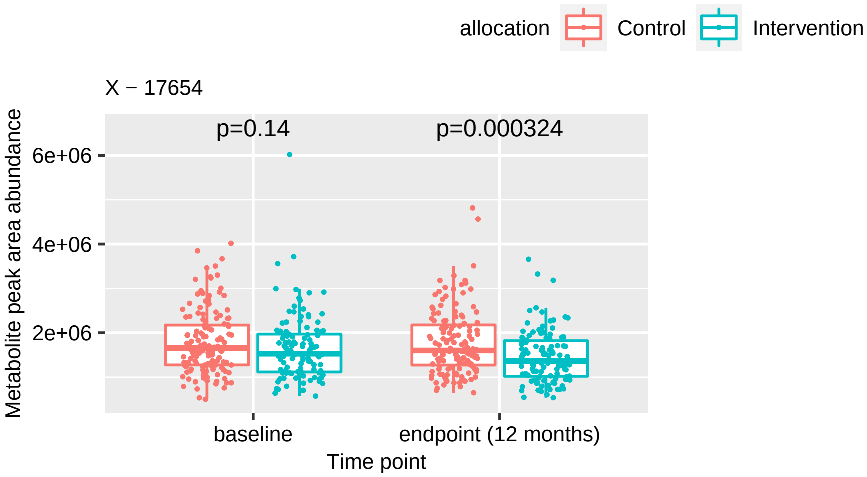

allocation
Control
Intervention
X − 17654
p=0.14
p=0.000324
Metabolite peak area abundance
6e+06
4e+06
2e+06
baseline
endpoint (12 months)
Time point
