## Supplementary figures and images for "The metabolomic signature of weight loss in the Diabetes Remission Clinical Trial (DiRECT)"

### Supplementary Document 5 - Figure S2B

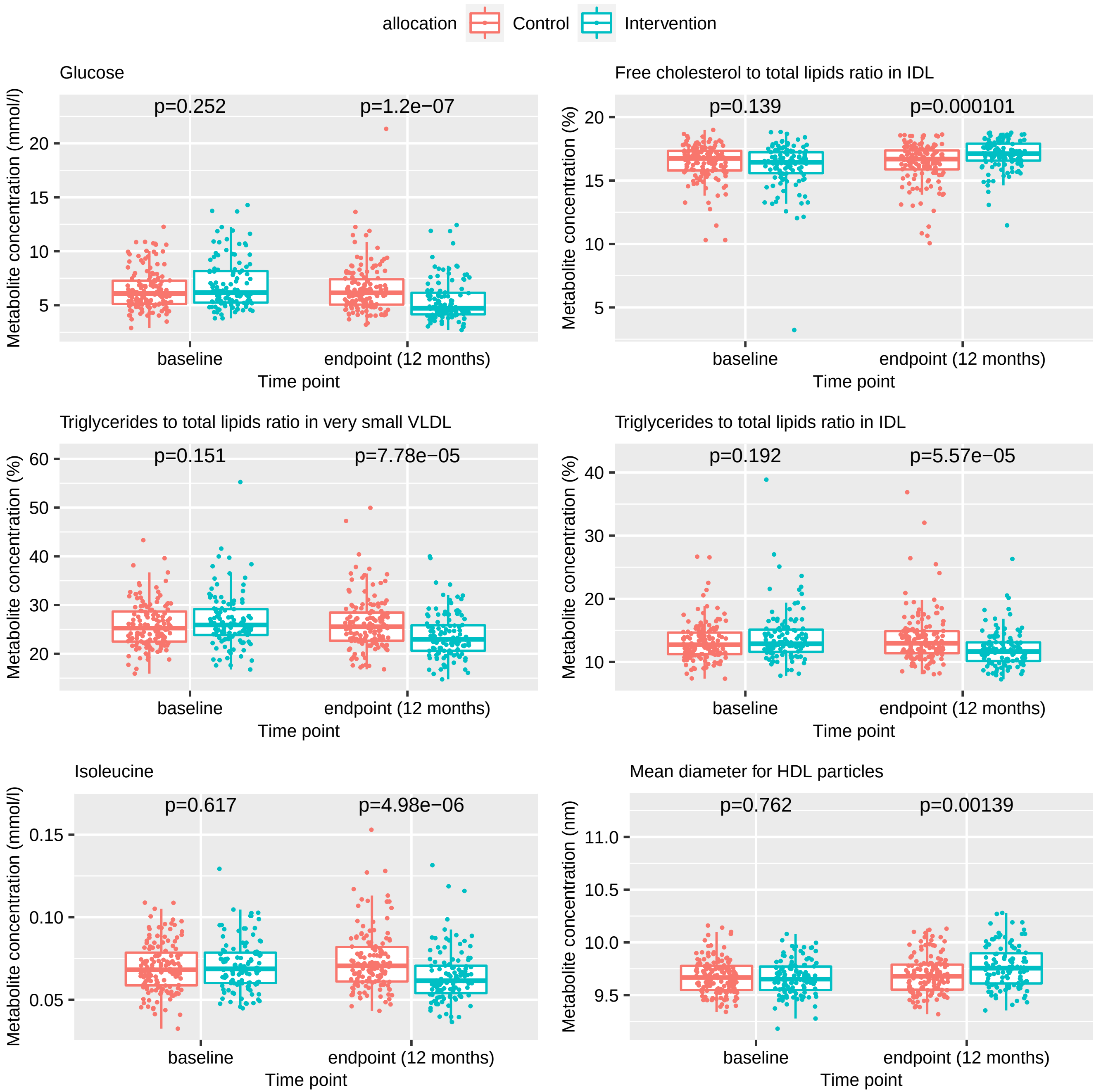

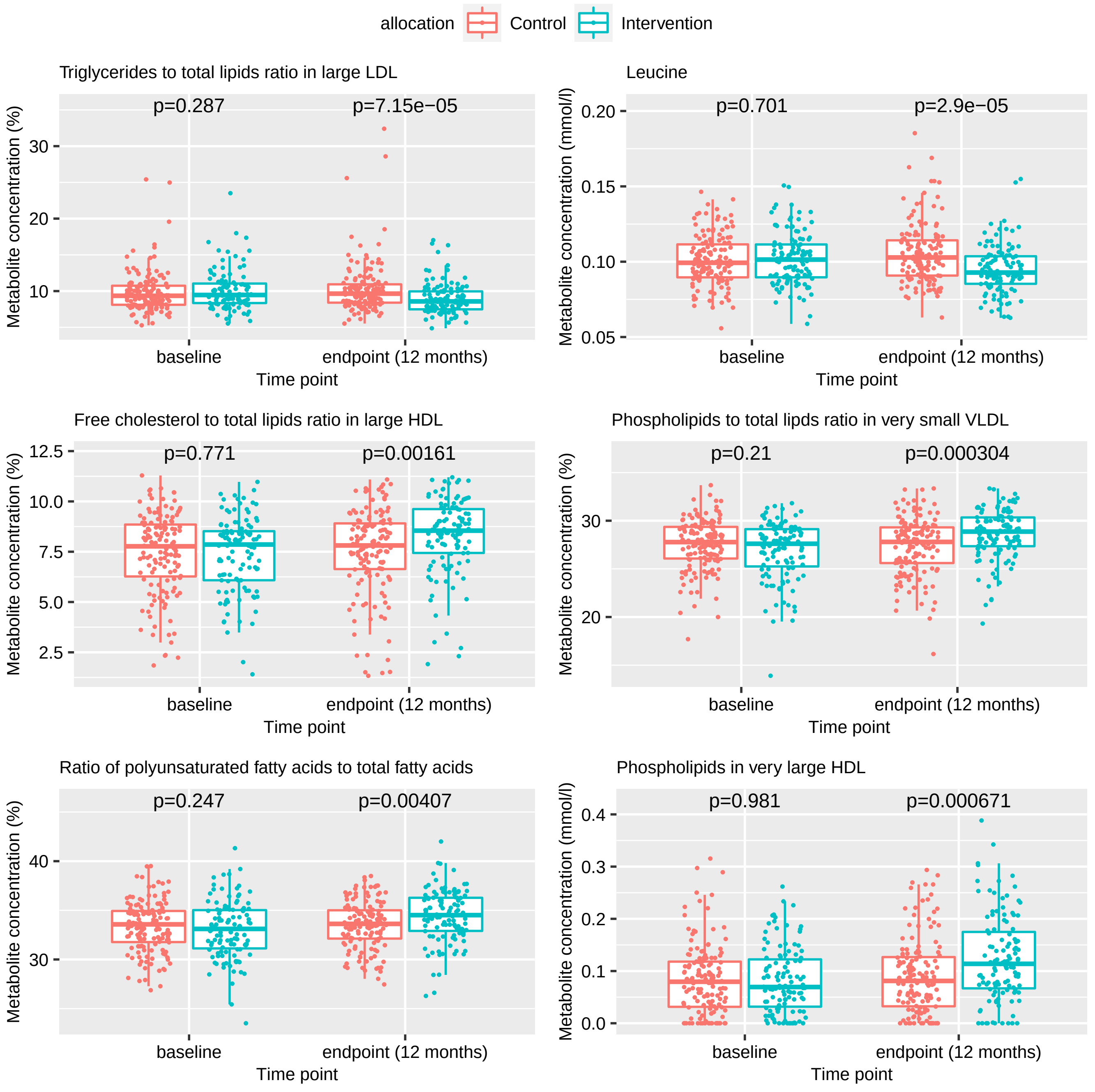

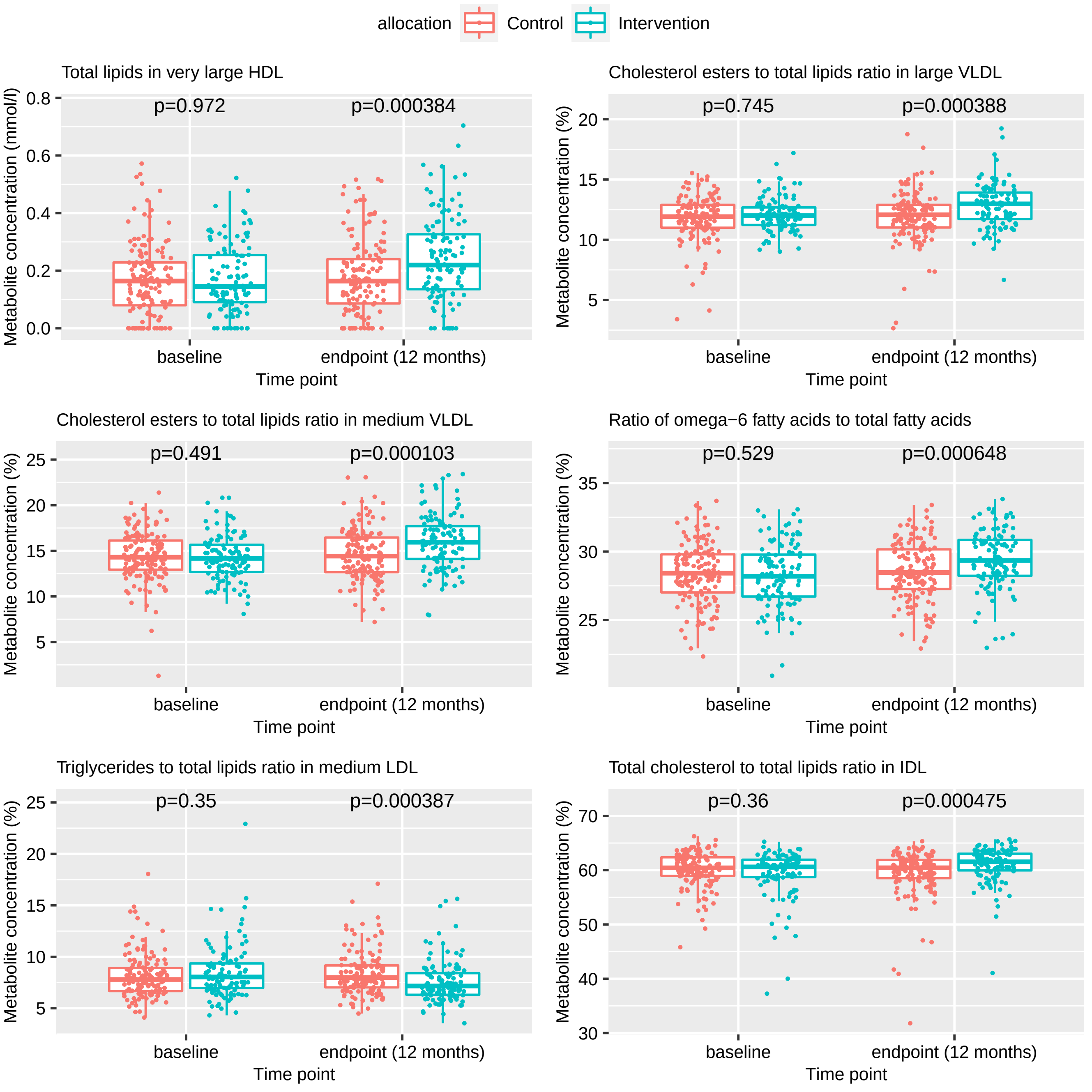

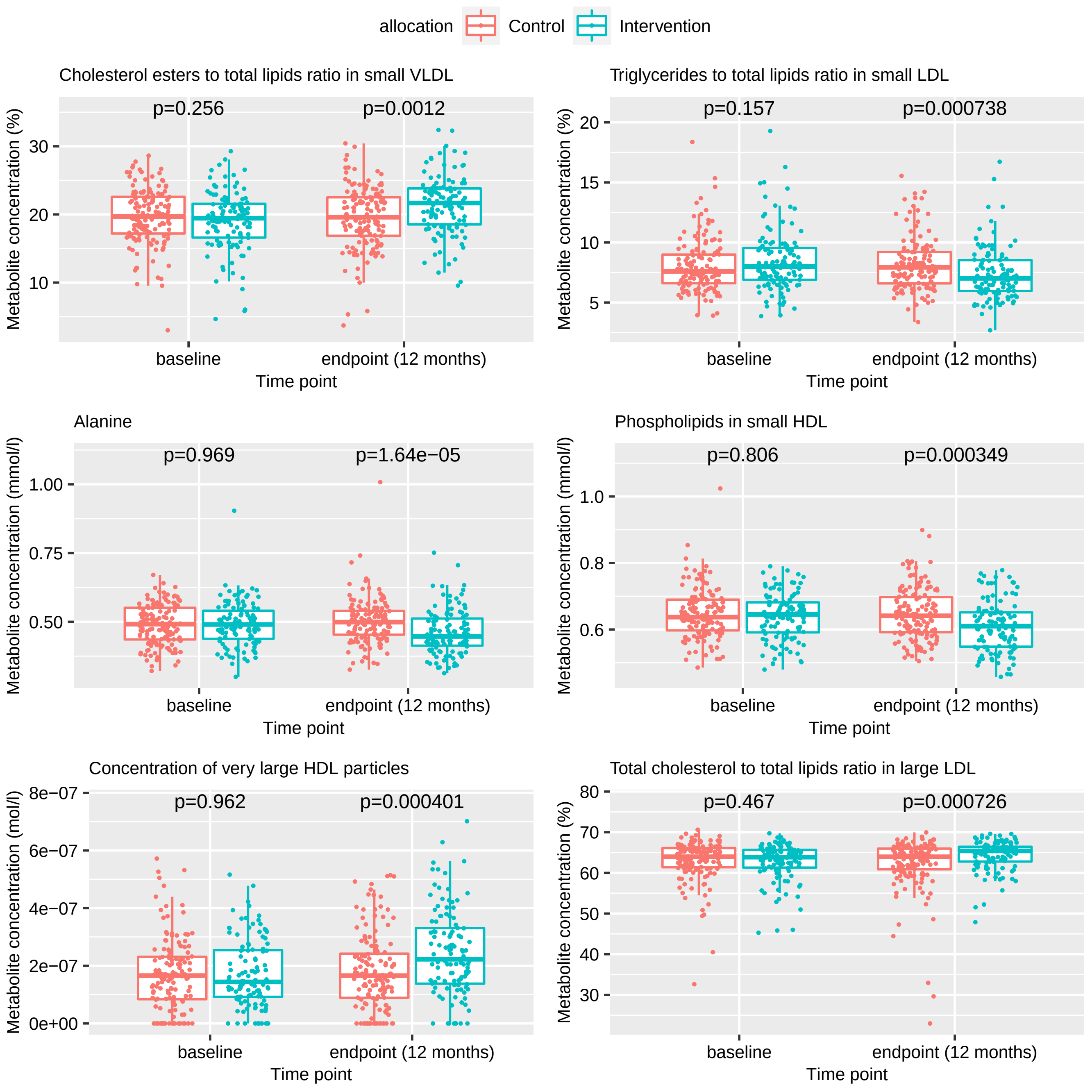

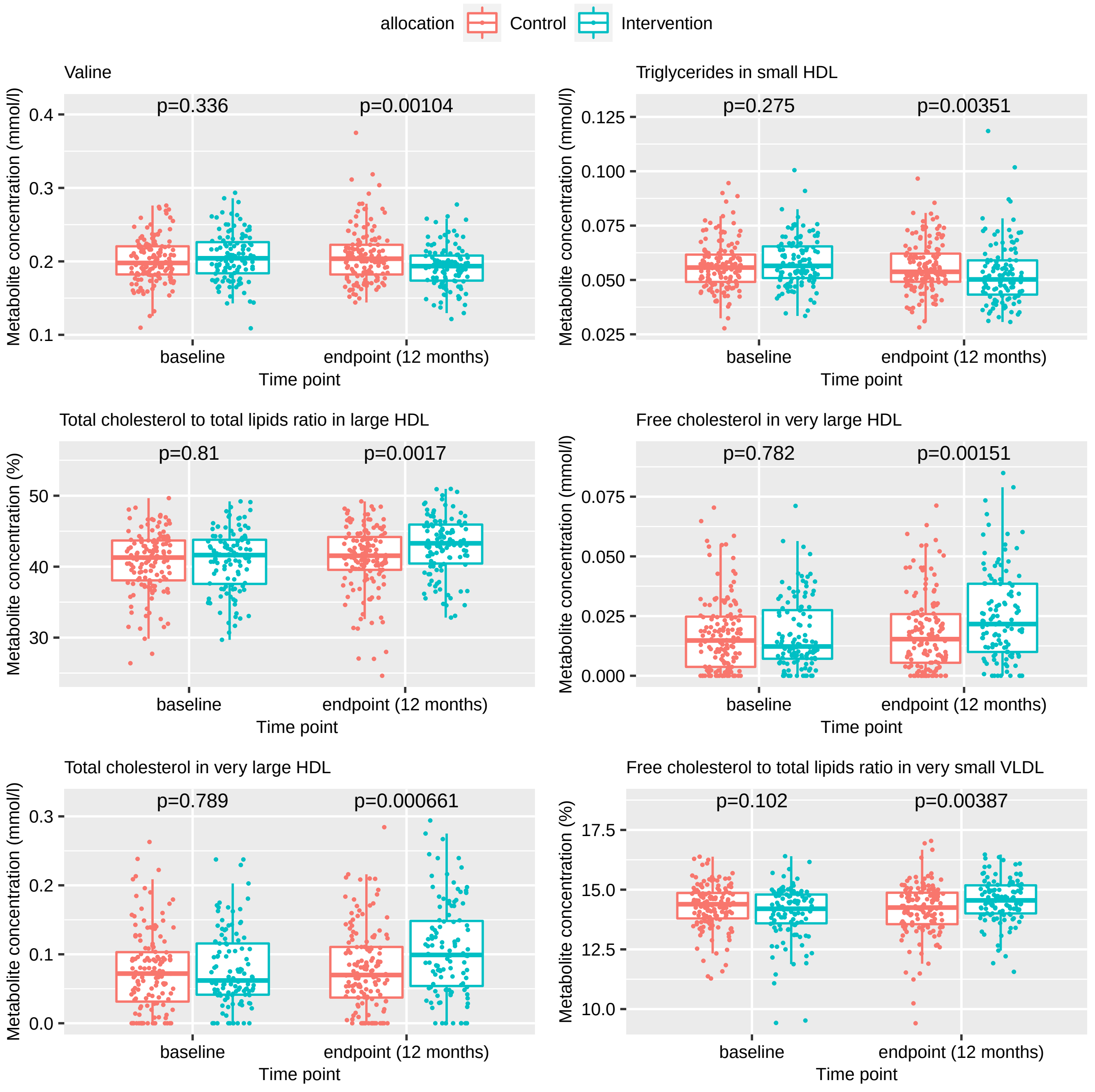

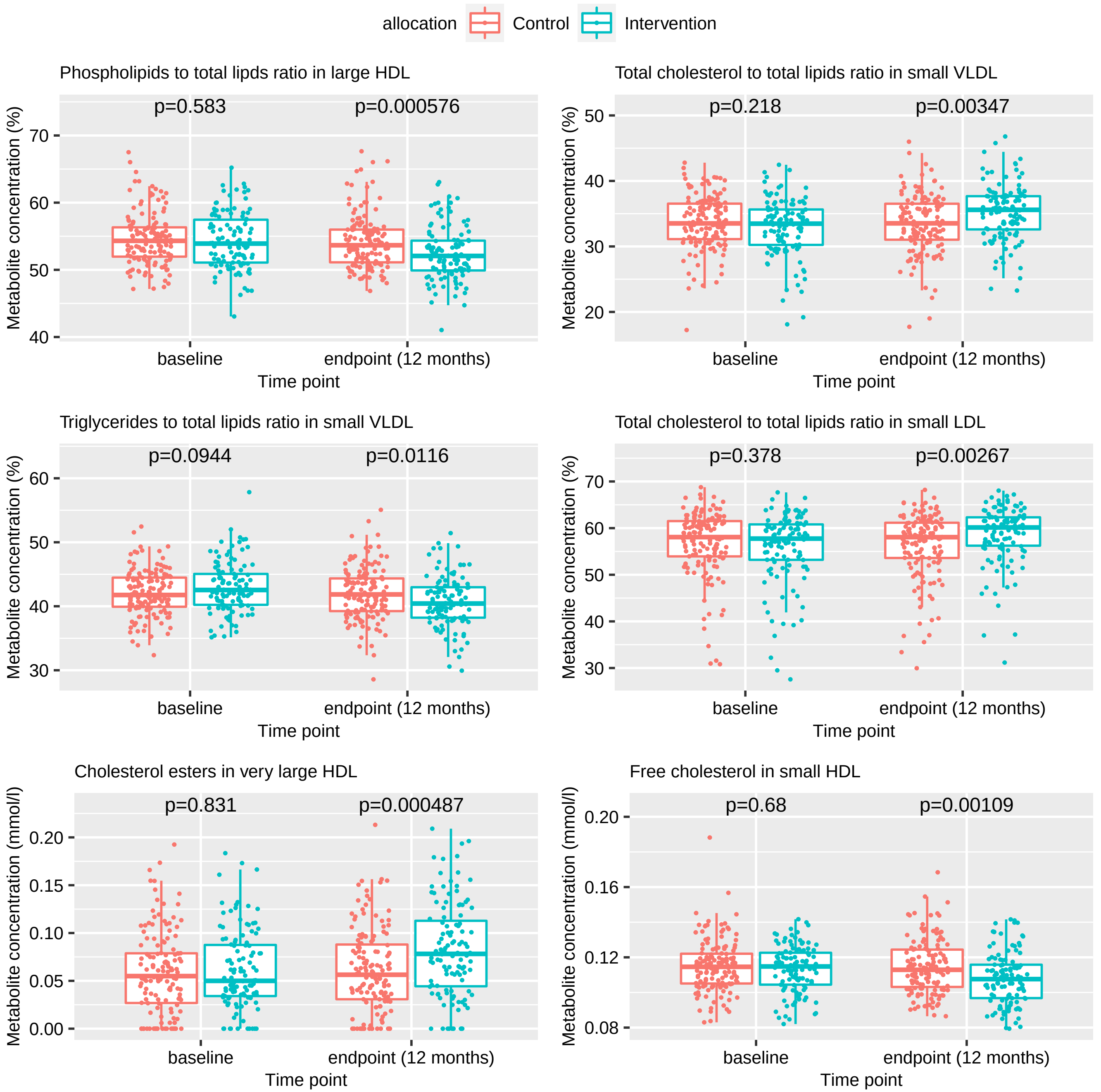

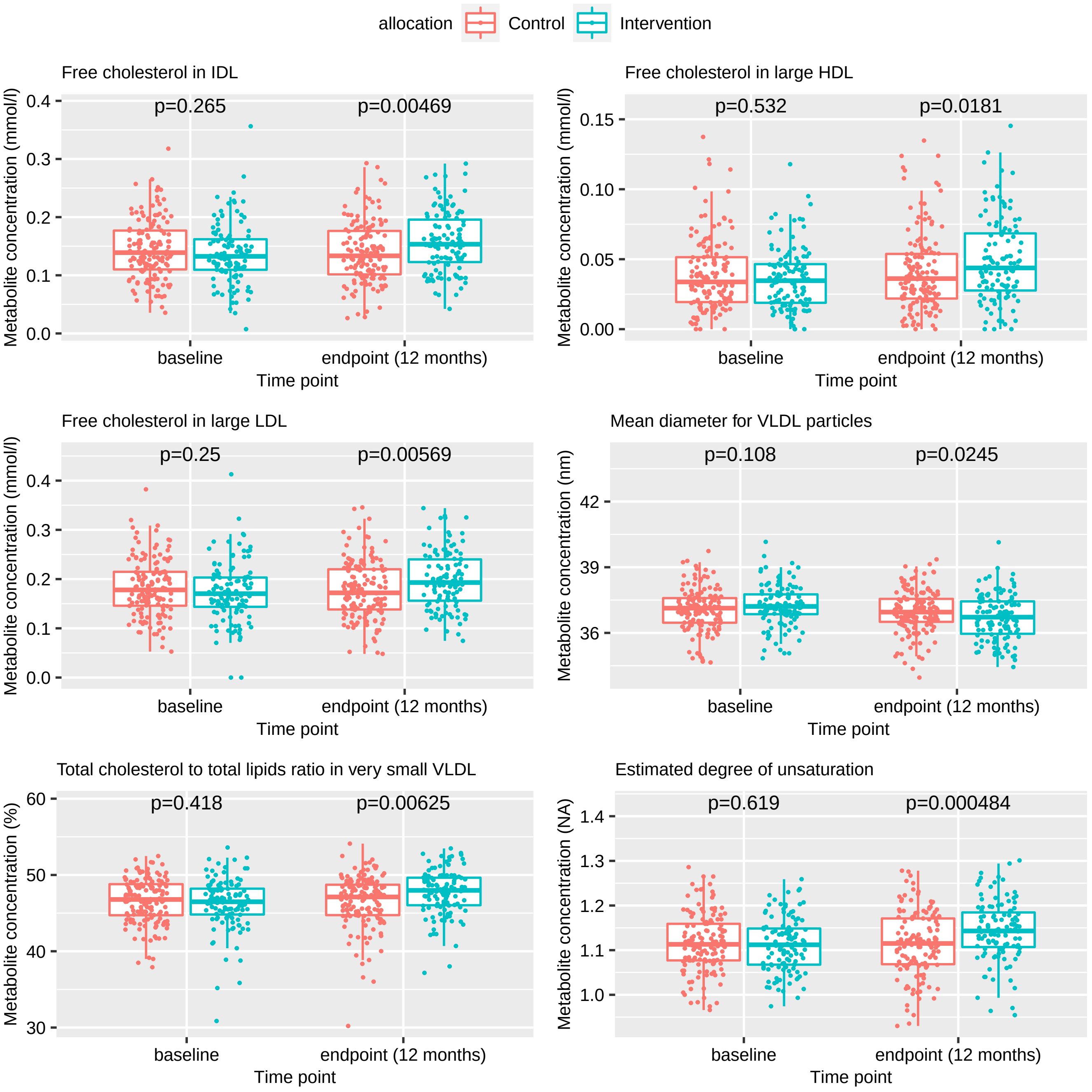

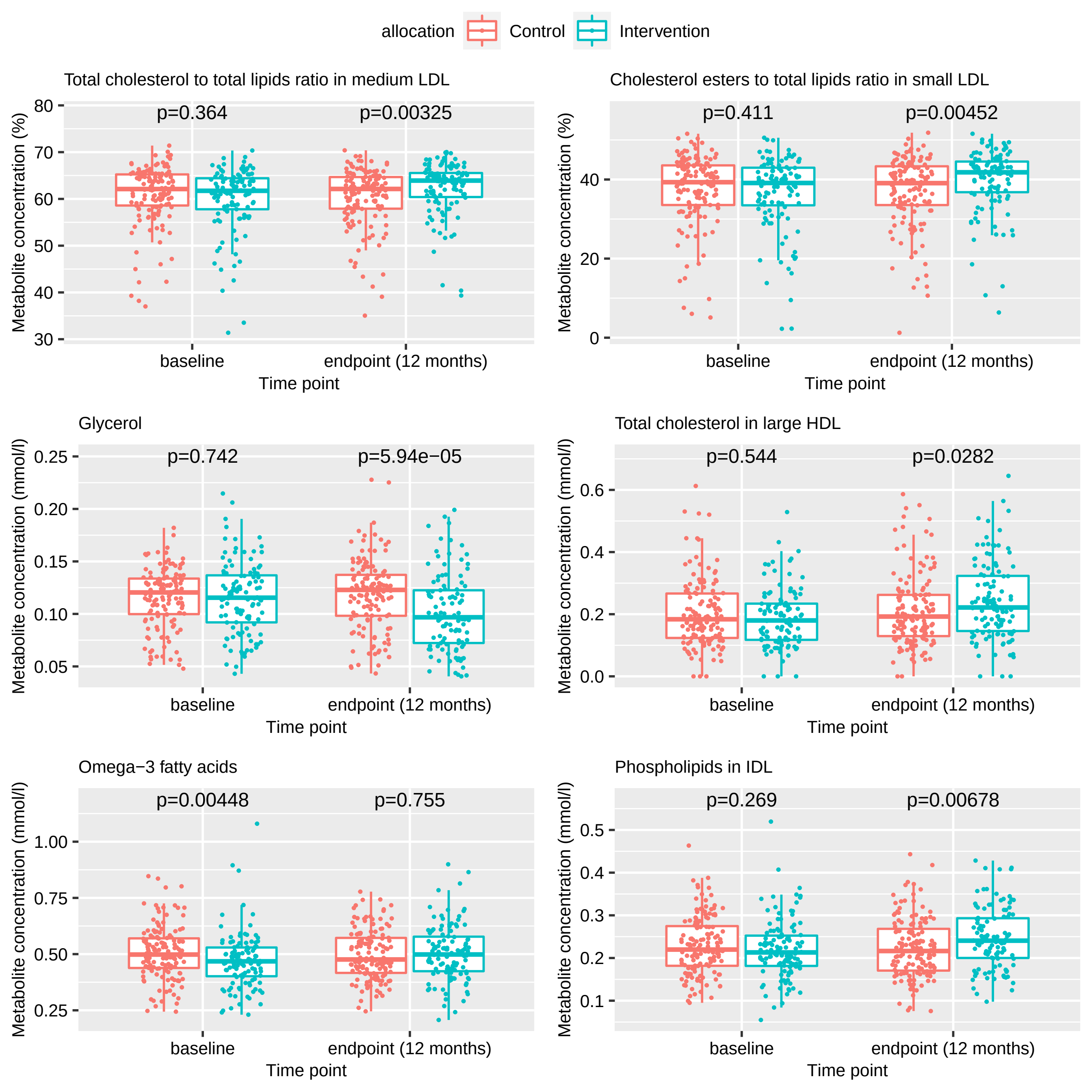

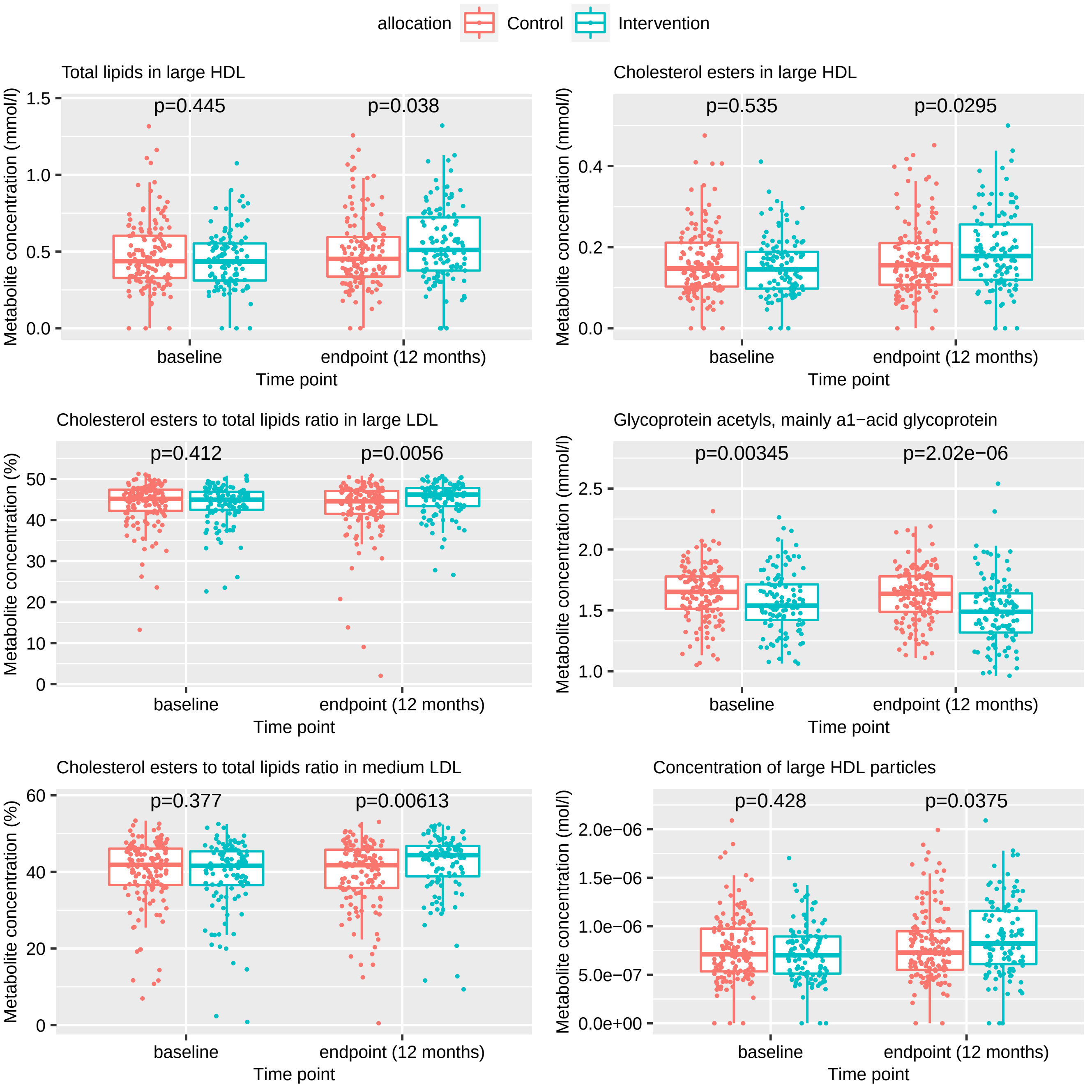

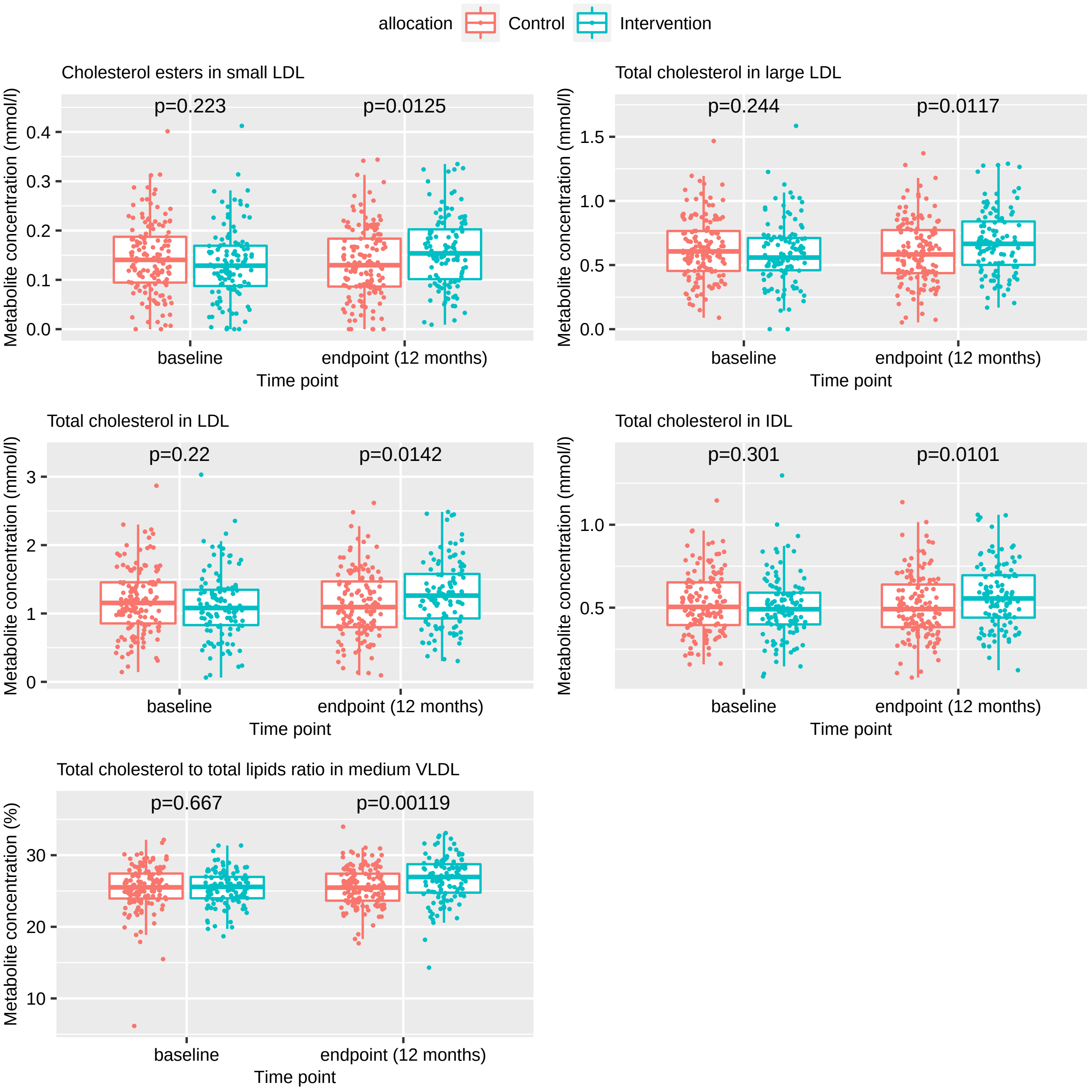

### Supplementary Document 6 - Figure S5

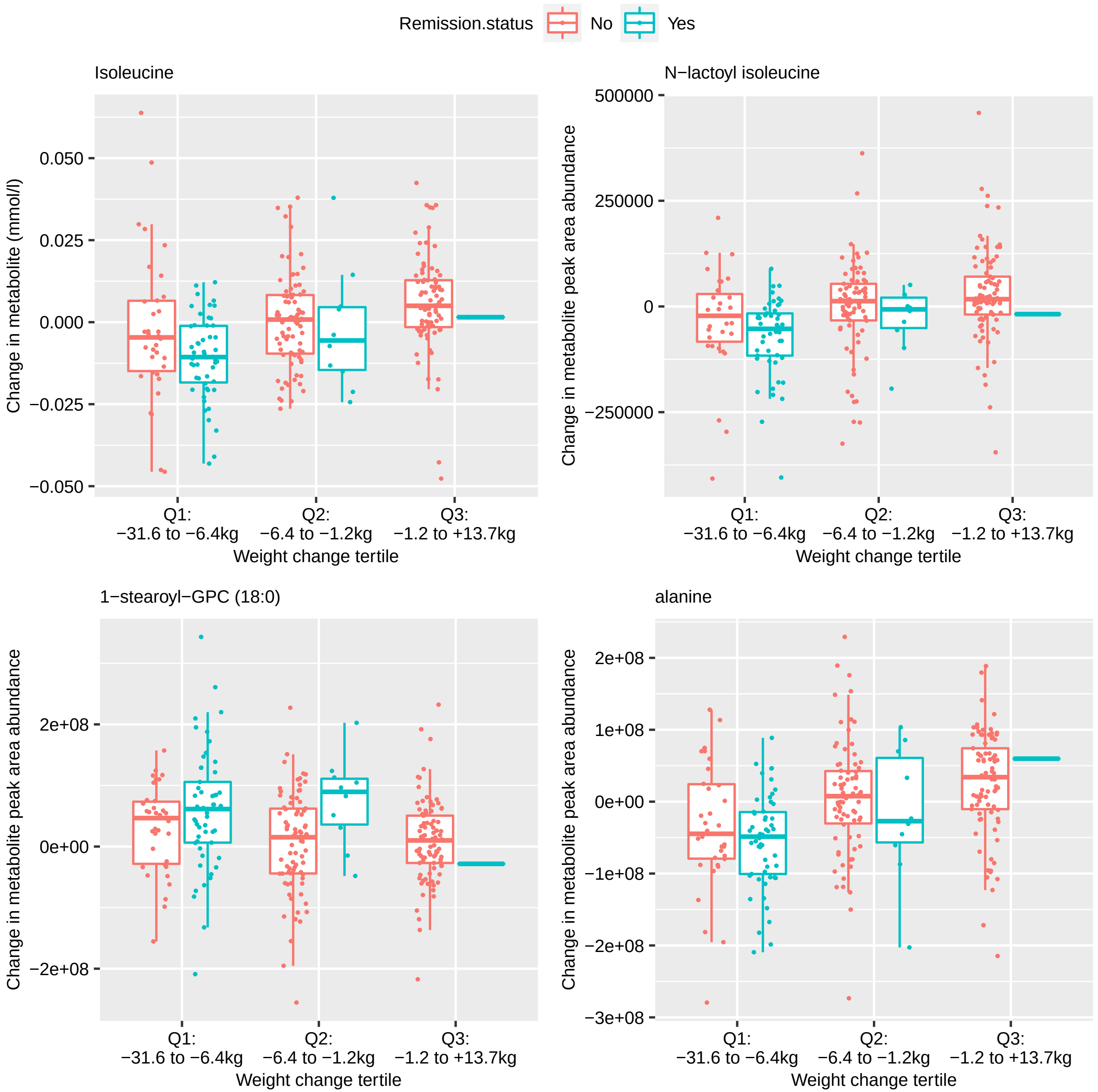

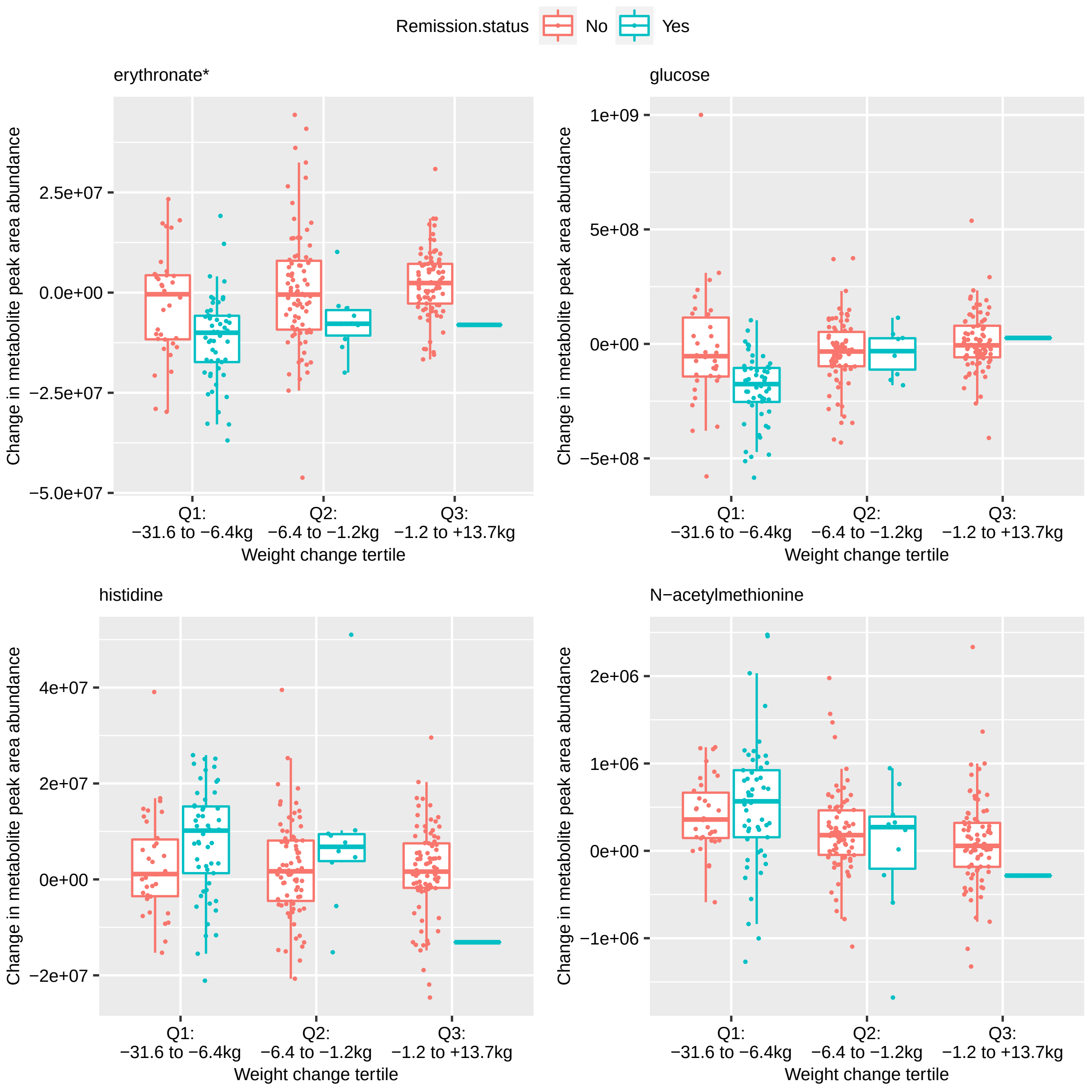

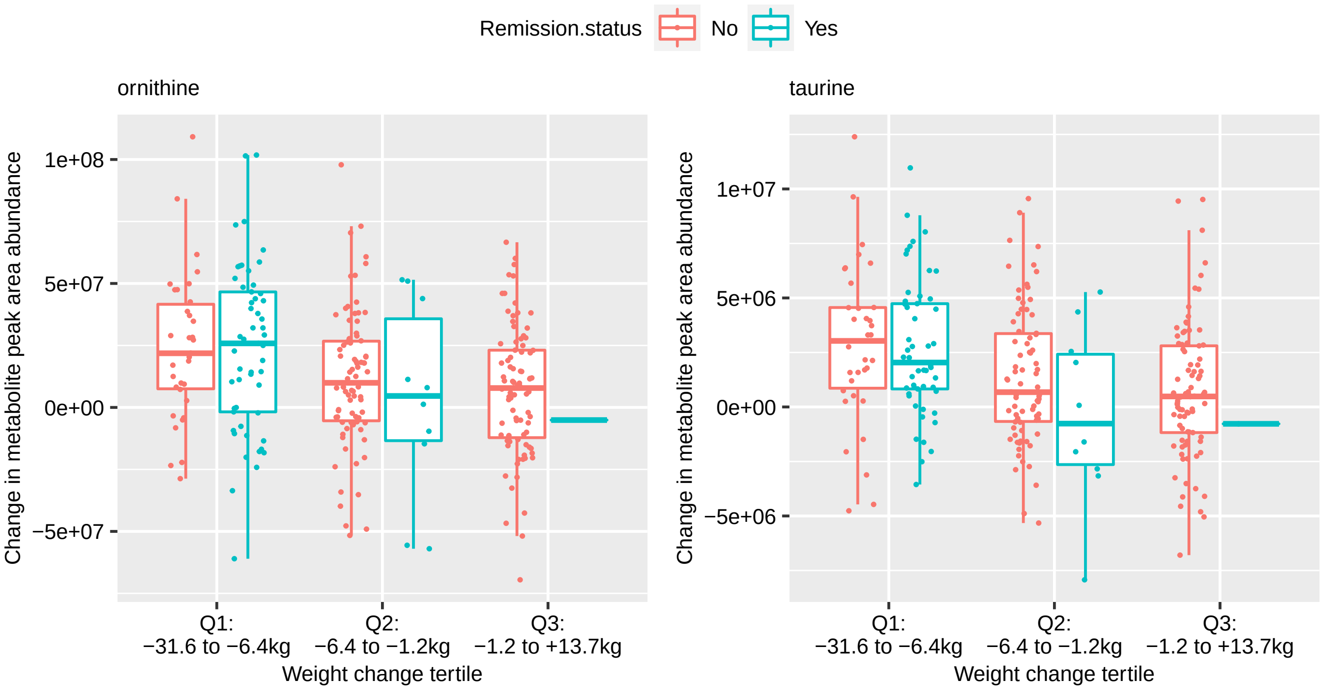
